# Mapping the dynamic genetic regulatory architecture of *HLA* genes at single-cell resolution

**DOI:** 10.1101/2023.03.14.23287257

**Authors:** Joyce B. Kang, Amber Z. Shen, Saori Sakaue, Yang Luo, Saisriram Gurajala, Aparna Nathan, Laurie Rumker, Vitor R. C. Aguiar, Cristian Valencia, Kaitlyn Lagattuta, Fan Zhang, Anna Helena Jonsson, Seyhan Yazar, Jose Alquicira-Hernandez, Hamed Khalili, Ashwin N. Ananthakrishnan, Karthik Jagadeesh, Kushal Dey, Accelerating Medicines Partnership Program: Rheumatoid Arthritis and Systemic Lupus Erythematosus (AMP RA/SLE) Network, Mark J. Daly, Ramnik J. Xavier, Laura T. Donlin, Jennifer H. Anolik, Joseph E. Powell, Deepak A. Rao, Michael B. Brenner, Maria Gutierrez-Arcelus, Soumya Raychaudhuri

**Affiliations:** Center for Data Sciences, Brigham and Women’s Hospital, Boston, MA, USA; Division of Genetics, Department of Medicine, Brigham and Women’s Hospital and Harvard Medical School, Boston, MA, USA; Department of Biomedical Informatics, Harvard Medical School, Boston, MA, USA; Program in Medical and Population Genetics, Broad Institute of MIT and Harvard, Cambridge, MA, USA; Division of Rheumatology, Inflammation, and Immunity, Department of Medicine, Brigham and Women’s Hospital and Harvard Medical School, Boston, MA, USA; Kennedy Institute of Rheumatology, University of Oxford, Oxford, UK; Division of Immunology, Boston Children’s Hospital, Harvard Medical School, Boston, MA, USA; Division of Rheumatology and the Center for Health Artificial Intelligence, University of Colorado School of Medicine, Aurora, CO, USA; Garvan Institute of Medical Research, Sydney, NSW, Australia; Division of Gastroenterology, Massachusetts General Hospital and Harvard Medical School, Boston, Massachusetts, USA; Harvard T. H. Chan School of Public Health, Boston, MA, USA; Psychiatric and Neurodevelopmental Genetics Unit, Massachusetts General Hospital, Boston, MA, USA; The Stanley Center for Psychiatric Research, The Broad Institute of MIT and Harvard, Cambridge, MA, USA; Institute for Molecular Medicine Finland (FIMM), University of Helsinki, Helsinki, Finland; Analytic and Translational Genetics Unit, Department of Medicine, Massachusetts General Hospital, Boston, MA, USA; Klarman Cell Observatory, Broad Institute of Harvard and MIT, Cambridge, MA, USA; Center for Computational and Integrative Biology, Massachusetts General Hospital and Harvard Medical School, Boston, MA, USA; Department of Molecular Biology, Massachusetts General Hospital and Harvard Medical School, Boston, MA, USA; Hospital for Special Surgery, New York, NY, USA; Weill Cornell Medicine, New York, NY, USA; Department of Medicine, University of Rochester Medical Center, Rochester, NY, USA

## Abstract

The human leukocyte antigen (HLA) locus plays a critical role in complex traits spanning autoimmune and infectious diseases, transplantation, and cancer. While coding variation in *HLA* genes has been extensively documented, regulatory genetic variation modulating *HLA* expression levels has not been comprehensively investigated. Here, we mapped expression quantitative trait loci (eQTLs) for classical *HLA* genes across 1,073 individuals and 1,131,414 single cells from three tissues, using personalized reference genomes to mitigate technical confounding. We identified cell-type-specific *cis-*eQTLs for every classical *HLA* gene. Modeling eQTLs at single-cell resolution revealed that many eQTL effects are dynamic across cell states even within a cell type. *HLA-DQ* genes exhibit particularly cell-state-dependent effects within myeloid, B, and T cells. Dynamic *HLA* regulation may underlie important interindividual variability in immune responses.

## Introduction

The human leukocyte antigen (HLA) genes, located within the major histocompatibility (MHC) region on chromosome 6, are central to the immune response. Classical HLA class I and II molecules trigger adaptive immunity by presenting antigens to CD8+ and CD4+ T cells, respectively. Positive and balancing selection has made the coding sequences of these genes among the most polymorphic in the genome (*1*). The HLA locus has the greatest number of associations with immune-mediated diseases and typically has larger effect sizes than all other loci combined (*1–4*). For example, the *HLA-C*06:02* allele is the major genetic risk factor for psoriasis (*5*), and *HLA-DRB1* alleles modulate risk for rheumatoid arthritis (RA) (*6*) and multiple sclerosis (*7*). *HLA* genes also play key roles in cancer by presenting neoantigens and in transplantation, where mismatched *HLA* alleles can result in rejection.

The regulatory mechanisms governing *HLA* genes are not yet well-understood. Previous studies have focused on coding variation altering HLA protein structure, which may affect antigen binding (*6, 8, 9*) or restrict the T cell receptor repertoire (*10–12*). However, mounting evidence indicates that noncoding *HLA* regulatory variation can influence disease (*13–15*). Higher *HLA-C* expression was found to control HIV infection but increase Crohn’s disease risk (*13*). Investigators have argued that risk alleles for systemic lupus erythematosus and vitiligo lie within regulatory regions that increase class II expression in myeloid cells (*14, 15*). Understanding the role of noncoding *HLA* variation in disease requires defining the genetic variation regulating *HLA* gene expression. Previous bulk RNA-sequencing studies have identified expression quantitative trait loci (eQTLs) for *HLA* genes in homogeneous cell lines (*16, 17*). However, *HLA* regulation may vary across cell states within a cell type. For example, we previously demonstrated that allele-specific expression of *HLA* class II changes dynamically in activated memory CD4+ T cells *in vitro* (*18*). Single-cell RNA-sequencing (scRNA-seq) may offer a more comprehensive understanding of *HLA* expression and its regulation by assaying cell states *in vivo* and mapping context-dependent eQTLs (*19–21*).

Because *HLA* genes are highly polymorphic, standard short-read sequencing pipelines that align reads to a single reference genome are biased when quantifying *HLA* expression (*22, 23*). Reads can fail to align if an individual’s allele is dissimilar from the reference allele, resulting in unmapped reads, or reads can “multi-map” to multiple *HLA* genes due to sequence similarity between genes (*24*). This bias confounds eQTL analysis, making it difficult to distinguish genuine genetic associations with *HLA* expression from inaccurate read alignment. In bulk data, personalized reference genomes accounting for individuals’ *HLA* genotypes can overcome this bias (*16, 17, 25, 26*). Here, we developed a personalized pipeline (scHLApers; **Fig. 1C**) extending this approach to single-cell data. We integrated four datasets (**Fig. 1A**) to explore how genetic regulation of classical *HLA* class I (*HLA-A*, *-B*, *-C*) and class II (*HLA-DPA1*, -*DPB1*, *-DQA1*, -*DQB1*, *-DRB1*) gene expression varies dynamically across diverse immune cell states (**Fig. 1D**), offering new insights into complex diseases.

**Fig. 1.**
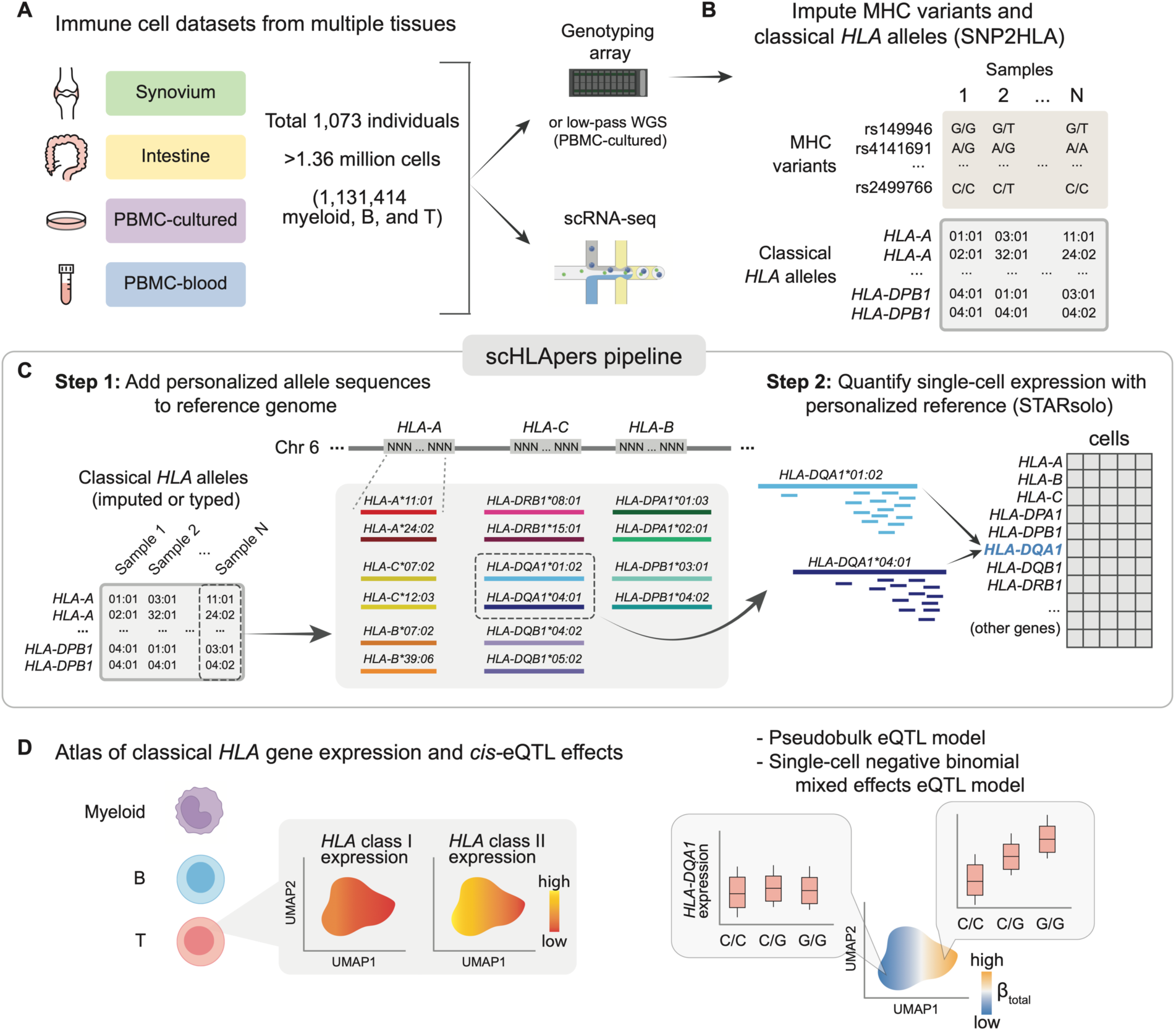
Overview of study and scHLApers pipeline. (**A**) We used four datasets with genotype and scRNA-seq data: Synovium (n=69 individuals, 275,323 cells), Intestine (n=22, 137,321), PBMC-cultured (n=73, 188,507), and PBMC-blood (n=909, 765,079). (**B**) Using the genotype data, we imputed SNPs within the MHC and one-and two-field classical *HLA* alleles. (**C**) Schematic of scHLApers pipeline, where scRNA-seq reads are aligned to a personalized reference for each individual based on classical *HLA* alleles. In the example, an individual is heterozygous for all eight *HLA* genes, so 16 additional contigs are added to the reference. Original reference gene sequences are masked with Ns. scHLApers outputs a whole-transcriptome counts matrix with improved *HLA* gene estimates. Both alleles contribute to count estimation for each gene. (**D**) We generated an atlas of *HLA* expression across all cell types (top) and mapped eQTLs for *HLA* genes in myeloid, B, and T cells (bottom). Schematic of example dynamic eQTL (bottom), where eQTL strength (slope, *β_total_)* changes across T cell states. Abbreviations: PBMCs, peripheral blood mononuclear cells; MHC, major histocompatibility complex; WGS, whole-genome sequencing. Icon of scRNA-seq from BioRender.

## Results

### Quantifying single-cell *HLA* expression with scHLApers

We developed scHLApers, a pipeline that accurately quantifies <u>s</u>ingle-<u>c</u>ell *<u>HLA</u>* expression using a <u>pers</u>onalized reference (**Fig. 1C, Methods, Supplementary Text 1**). First, scHLApers uses an individual’s unique classical *HLA* alleles (**Fig. 1B**) to add the personalized genomic sequences for each two-field allele from the IPD-IMGT/HLA database (*27*) to the standard reference genome in place of the original *HLA* gene sequences. scHLApers then uses STARsolo (*28*) to quantify whole-transcriptome expression in single-cells with multimapping.

### Four cohorts with genotype and single-cell transcriptomic data

To study immune cell states from diverse tissues and biological conditions, including some from disease conditions, we used four scRNA-seq datasets with paired genotype data (**Table S1, Fig. 1A, Fig. S1**). After quality control (**Methods, Table S2**), the combined dataset of 1,073 individuals comprised synovial joint biopsies from an RA cohort (*29*) (Synovium, n=69 individuals), intestinal biopsies from an ulcerative colitis (UC) cohort (*30*) (Intestine, n=22), peripheral blood mononuclear cells (PBMCs) from healthy males cultured *in vitro* with influenza A virus and control conditions (*31*) (PBMC-cultured, n=73), and PBMCs from a large Australian cohort (*32*) (PBMC-blood, n=909).

### Imputing *HLA* alleles and MHC variants

Using SNP2HLA with our group’s multi-ancestry *HLA* reference panel (*24, 33, 34*) (**Methods, Fig. 1B, Fig. S2**), we inferred a common set of 12,050 variants in the MHC with imputation dosage R^2^>0.8 and MAF>1% in each cohort. These included 11,938 SNPs and 112 one-and two-field alleles for classical *HLA* genes (**Fig. 2A, Table S3**). We used the two-field alleles to quantify expression with scHLApers (**Fig. 1C**), and we used both types of variation as input for downstream eQTL analysis (**Fig. 1D**).

**Fig. 2.**
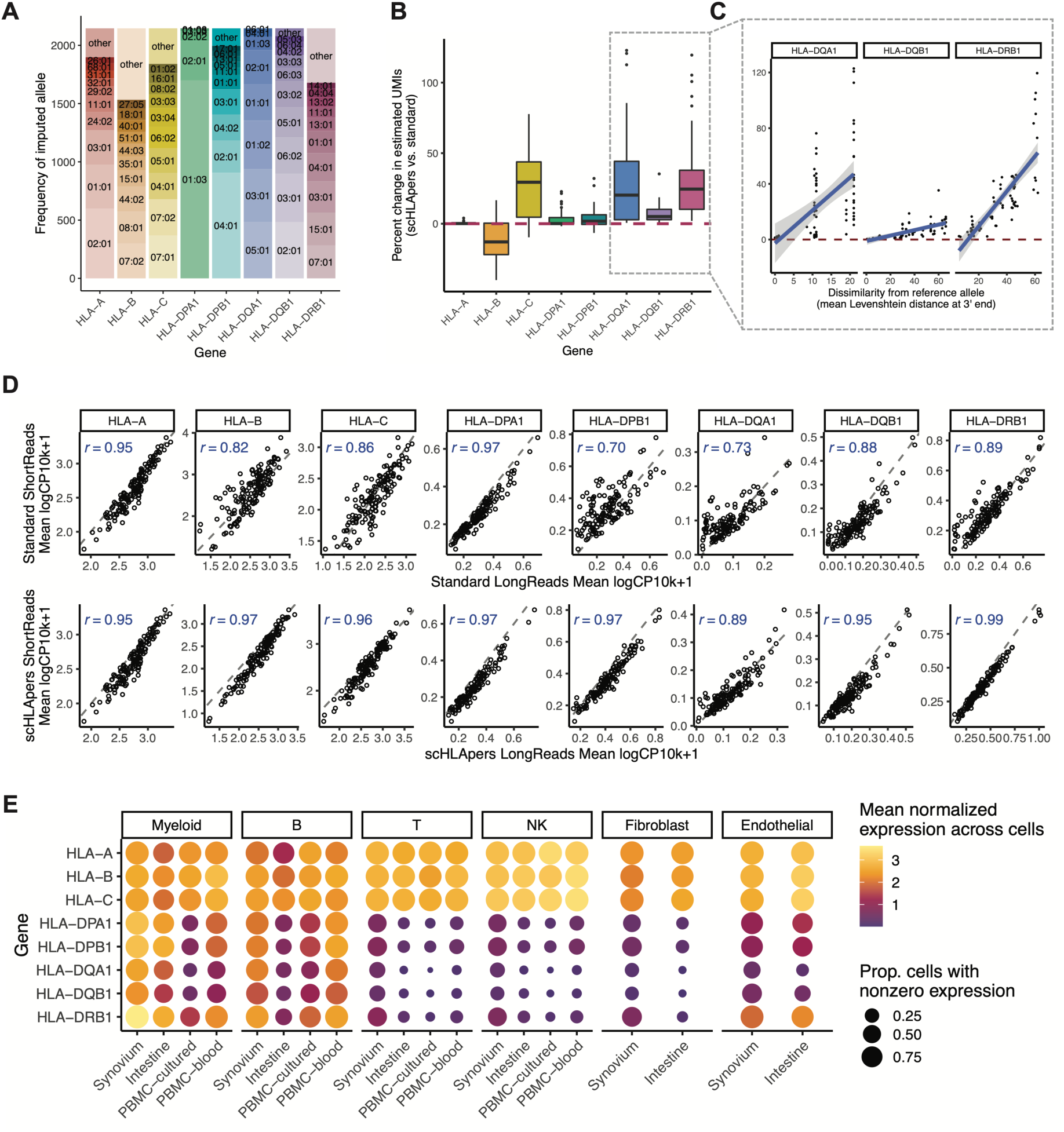
Quantifying single-cell *HLA* expression using a personalized pipeline. (**A**) Frequency of each imputed two-field *HLA* allele across all cohorts. Most common alleles (up to ten) are labeled for each gene, with other alleles grouped into “other.” Alleles with frequency of less than five are not labeled. (**B**) Boxplot (each observation is one sample) showing percentage change (*y*-axis) in the estimated UMIs for each *HLA* gene (*x*-axis) summed across all cells after quantification with scHLApers (compared to a pipeline using the standard reference genome) in the Synovium dataset (n=69 individuals). The center line of the boxplot represents the median, the lower and upper box limits represent the 25% and 75% quantiles, respectively, the whiskers extend to the box limit ±1.5 × IQR, and outlying points are plotted individually. Plot for all cohorts is in **Fig. S4C**. Dotted red line denotes no change. (**C**) Percentage change in estimated expression (*y*-axis) for three example genes (*HLA-DQA1*, *HLA-DQB1*, and *HLA-DRB1*) per sample as a function of the mean (between the individual’s two alleles) Levenshtein distance relative to the reference allele at the 3’ end (*x*-axis). Dotted red line denotes no change. Plot for all genes is in **Fig. S4D**. (**D**) Comparing the estimated *HLA* expression as measured using shorter reads (84 bp, *y*-axis) versus longer reads (289 bp, *x*-axis) in the standard pipeline (top row) compared to scHLApers (bottom row). Each dot shows the mean log(CP10k+1)-normalized expression across cells for one sample in the PBMC-cultured dataset (n=146 samples from 73 individuals). *r* is calculated as Pearson correlation; dashed gray line is the identity line. (**E**) *HLA* expression in different cell types across cohorts: myeloid (n=145,090 cells), B (n=180,935), T (n=805,389), NK (n=125,865), fibroblasts (n=82,651) and endothelial (n=26,300). Dot size indicates the proportion of cells with nonzero expression; color indicates log(CP10k+1)-normalized expression (mean across cells).

### Assessing the performance of scHLApers

We assessed the performance of scHLApers compared to a pipeline without personalization, i.e., using the standard GRCh38 reference genome (**Methods, Figs. S3-5**). We expected estimated *HLA* gene expression to generally increase with scHLApers since it rescues previously unmapped reads. For each individual, we calculated the percentage change in the total UMI count for each *HLA* gene across all cells after personalization. Personalization indeed generally led to higher estimated expression (**Fig. 2B**), with concordant trends across cohorts (**Fig. S4C**). We reasoned that if scHLApers aligns reads more appropriately, then personalization should have larger effects for individuals whose alleles diverge more from reference genome alleles. To assess this, we compared the percentage change in estimated expression per individual to their alleles’ sequence dissimilarity to the reference (based on Levenshtein distance, **Methods**). For all genes except *HLA-B*, individuals with alleles more different from the reference tended to show a greater increase in expression after personalization (**Fig. 2C, Fig. S4D**). The genes whose expression increased the most per individual were *HLA-DRB1* (+29% change, 25th to 75th percentile [+10% to 38% change] in Synovium), *HLA-DQA1* (+29% [+3% to 44%]), *HLA-C* (+26% [+5% to 44%]), and *HLA-DQB1* (+7% [+3% to 10%]), consistent with prior findings in bulk RNA-seq (*17*). Expression of *HLA-DPB1*, *HLA-DPA1*, and *HLA-A* also increased but to a lesser extent (**Table S4**). Unexpectedly, we observed an overall decrease in *HLA-B* counts across all cohorts (**Fig. S4C**). After detailed investigation, we determined this was attributable to improved read alignments from *HLA-B* to *HLA-C* (**Supplementary Text 1**). For individuals with both *HLA-C* alleles similar to the reference allele (*HLA-C*07:02*), *HLA-B* was less affected by personalization (**Fig. S5B,C**). In contrast, for individuals with at least one “non-reference-like” *HLA-C* allele (i.e., other than *HLA-C*07:02*), more reads that aligned to *HLA-B* before personalization aligned better to *HLA-C* after personalization, leading to appropriately decreased *HLA-B* counts.

To assess if scHLApers improved the consistency of expression quantification, we leveraged the fact that each PBMC-cultured library was sequenced using two read lengths (289 bp and 84 bp). We reasoned that a standard pipeline might lead to inconsistent quantification between the longer and shorter read versions of the dataset due to different types of mapping biases for different read lengths. In contrast, personalization should result in consistent quantification of each *HLA* gene between the two versions. Indeed, personalization increased the correlation between the estimated expression in shorter and longer-read data for all genes across samples (**Fig. 2D**; *HLA-B* Spearman *r* = 0.97 scHLApers vs. 0.82 standard, *HLA-C r =* 0.96 vs. 0.86, *HLA-DPB1 r* = 0.97 vs. 0.70). Together, our results demonstrate that aligning reads to a personalized reference improves precision in quantifying single-cell *HLA* expression.

### *HLA* expression across major cell types

After removing low-quality cells (**Table S2, Supplementary Text 2, Fig. S6A-C**), we grouped cells from the four datasets into six major cell types (**Methods, Table S5**) to investigate cell-type-specific *HLA* expression using scHLApers. These include four immune cell types from all cohorts: 145,090 myeloid cells (monocytes, macrophages, and dendritic cells), 180,935 B cells (including plasma), 805,389 T cells, and 125,865 NK cells. It also includes stromal cells from the two solid tissue datasets: 82,651 fibroblasts and 26,300 endothelial cells. As expected, we found that all cell types highly express *HLA* class I genes across tissues, consistent with ubiquitous presentation of self-peptides, whereas class II expression varied (**Fig. 2E**). Specifically, myeloid cells and B cells expressed the highest levels of class II, consistent with their role as professional antigen-presenting cells. Interestingly, all other cell types, such as T cells, also express class II genes, albeit at lower levels. Human T cells have been previously observed to express *HLA* class II upon activation (*18, 35*– *37*), though its function is not well-understood (*38–40*).

### Multi-cohort analysis identifies *HLA* regulatory variants

To identify eQTLs for classical *HLA* genes, we tested the 12,050 MHC-wide variants (**Fig. 3A, Table S6**) for association with the expression of each *HLA* gene in myeloid, B, and T cells. We chose these three cell types because they are well-represented in all datasets and have known roles in antigen presentation (myeloid and B) or prior evidence for state-dependent *HLA* regulation (T) (*18*). For each cell type and individual, we aggregated single-cell expression profiles into a single “pseudobulk” measurement (**Methods, Fig. S6D**). We used linear regression and analyzed all four cohorts together, controlling for covariates and testing 289,200 pairs of variants and *HLA* genes (**Methods, Fig. S7, Table S7**).

**Fig. 3.**
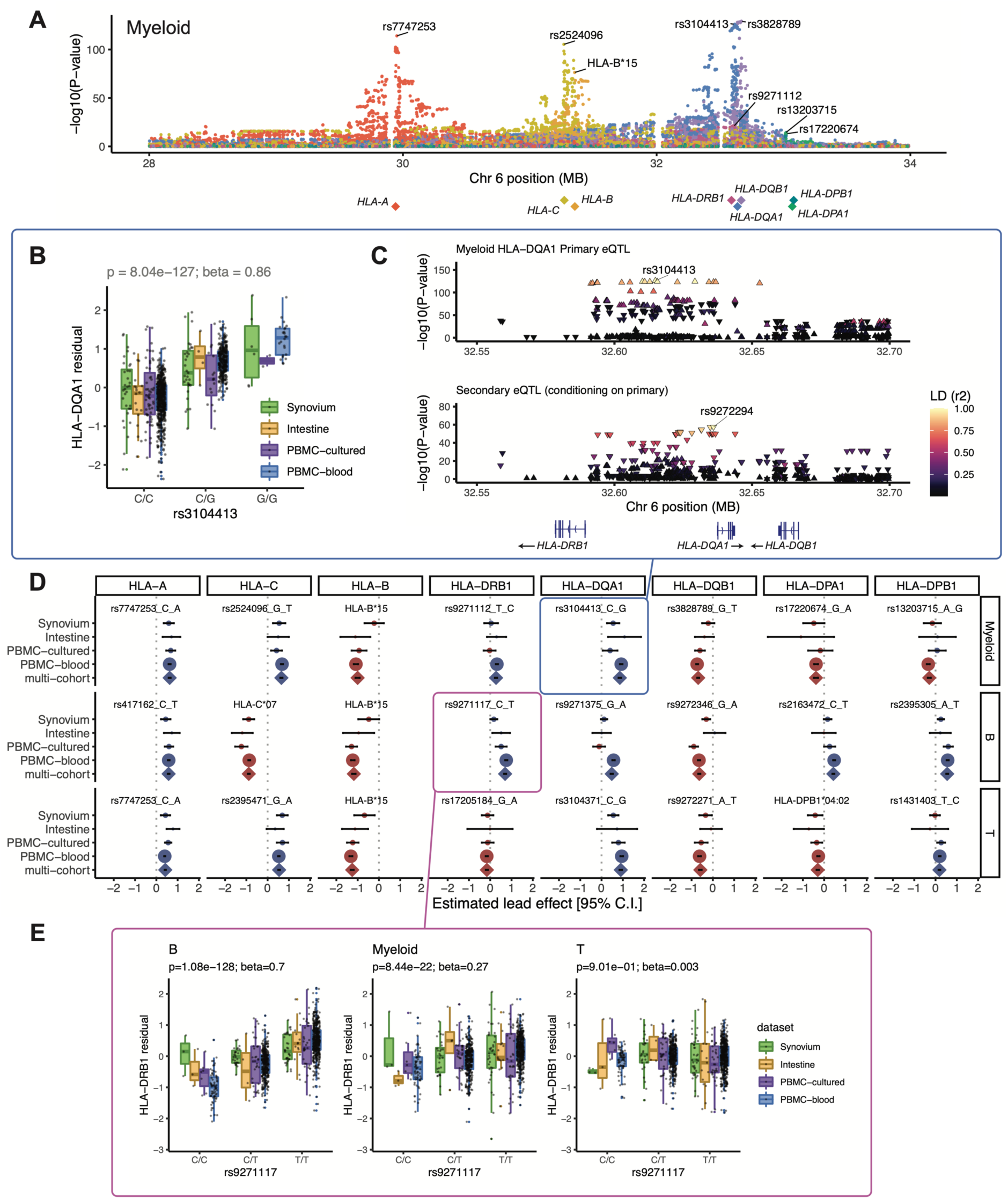
eQTLs for classical *HLA* genes from pseudobulk analysis. (**A**) Manhattan plot showing the significance (*y*-axis) of association between tested MHC variants (*x*-axis) and expression of each *HLA* gene (color) in myeloid cells from the multi-cohort model. Most significant (lead) eQTLs are labeled. Diamond points indicate TSS of each gene. (**B**) Boxplot showing an example lead eQTL (rs3104413). Increased dosage of the G allele (*x*-axis) associates with higher *HLA-DQA1* expression in myeloid cells (*y*-axis: units are the residual of inverse normal transformed mean log(CP10k+1)-normalized expression across cells after regressing out covariates), plotted by dataset (color). All lead eQTLs shown in **Figs. S8-10**. (**C**) Locus zoom plot for the primary (rs3104413) and secondary (rs9272294) eQTLs for *HLA-DQA1* in myeloid cells. Significance of association (*y*-axis) is shown for nearby variants on chromosome 6 (*x*-axis); color denotes LD (*r*^2^ with lead eQTL in multi-ancestry HLA reference). Triangles point upwards for a positive (downwards for negative) effect on expression. Gene bodies and direction of transcription (arrows) for *HLA-DRB1*, *HLA-DQA1*, and *HLA-DQB1* are underneath. (**D**) Grid showing lead eQTLs for each *HLA* gene (columns) in each cell type (rows: myeloid, B, and T). Each element of the grid includes a forest plot with the estimated lead effect size (*x*-axis) and 95% confidence interval (±1.96 s.e.) of the estimate from the multi-cohort analysis (represented by a diamond) and the same variant-gene pair tested for an association within each cohort separately (represented by dots above). The size of the dots/diamond indicates cohort size; color indicates sign of the ALT allele’s effect on expression (blue indicates positive, red indicates negative). The eQTLs boxed in blue and magenta are highlighted in (**B, C**) and (**E**), respectively. (**E**) Example of a cell-type-dependent eQTL (rs9271117) that was the lead eQTL for *HLA-DRB1* and strongest in B cells. Boxplots are formatted analogously to panel (**B**) and show the eQTL variant’s effect for all three cell types separately.

We detected an eQTL for every *HLA* gene in every cell type (*P*-values < 4 × 10^−9^; **Fig. 3B-E, Figs. S8-10, Table S8**). Calculating the effect size of each lead eQTL in each cohort separately, we observed 91.7% (88/96) mean directional concordance across cohorts (**Fig. 3D, Table S9**), suggesting consistent effects across datasets. The B cell results were highly concordant with a previous study on *HLA* eQTLs (*17*), which used bulk RNA-seq data from lymphoblastoid cell lines and found that all eight variants included in both studies showed consistent directions of effect (Pearson *r* = 0.92, **Fig. S11A**).

Most lead variants (19/24) were individual SNPs within the MHC. For example, rs3104413, the lead variant for *HLA-DQA1* in myeloid cells, is located between *HLA-DRB1* and *HLA-DQA1* (*P* = 8.04 × 10^−127^; **Fig. 3B,C**). This SNP commonly cooccurs with the classical *HLA-DQA1*03:01* allele (87.5% of *DQA1*03:01* haplotypes are in phase with the G allele of rs3104413, **Table S10**). The *HLA-DQA1*03:01* allele is part of the DQ8 haplotype, which is associated with type 1 diabetes and celiac disease (*41*).

Some lead eQTLs were individual one-or two-field *HLA* alleles. For example, *HLA-B*15* was the lead eQTL for *HLA-B* in all three cell types (*P* < 3 × 10^−81^) and associated with lower expression of *HLA-B* (**Fig. S11B,C**). A recent study using a new capture RNA-seq method also found that *HLA-B*15* alleles were among the lowest expressed in bulk PBMCs, consistent with our observations (*42*). *HLA-C*07* was the most significant variant for *HLA-C* in B cells (*P* = 2.87 × 10^−210^, **Fig. S9, Fig. S11B**), reflecting reduced expression of *HLA-C*07* alleles relative to other *HLA-C* alleles. This finding could not be explained by read alignment bias (**Fig. S11C**) and is supported by previous work showing that *HLA-C*07* alleles contain a 3’ UTR miRNA binding site that reduces *HLA-C* expression (*43, 44*). Interestingly, the *HLA-C*06:02* and *HLA-C*12:03* alleles, major risk factors for psoriasis (*5*), were associated with higher *HLA-C* expression in all three cell types (*P* < 8 × 10^−40^ and 3 × 10^−8^, respectively, **Table S7**). The increased expression of these *HLA-C* alleles may contribute to psoriasis disease risk (*45*).

### *HLA* eQTLs are cell-type-dependent

We next explored whether *HLA* eQTLs are cell-type-dependent, as reported for other genes (*32, 46*). To test this, we used a mixed-effects model including an interaction term for cell type with genotype (**Methods**). Almost all (22/24) eQTLs exhibited statistically significant cell-type-dependency (interaction *P* < 2.08 × 10^−3^ = 0.05 / 24 tests), and several showed dramatic effects (**Table S11**). The strongest example was the lead eQTL for *HLA-DRB1* in B cells (rs9271117, β = 0.7, *P* = 1.08 × 10^−128^), which was ∼3-fold weaker in myeloid cells (β = 0.27, *P* = 8.44 × 10^−22^) and altogether absent in T cells (*P* = 0.90) (**Fig. 3E**). Similarly, eQTLs for *HLA-DPA1* and *HLA-DPB1* (rs2163472 and rs2395305) exhibited much stronger regulatory effects in B cells compared to myeloid and T cells (**Fig. S12A,B**, β = 0.43 in B vs. 0.04 and 0.08 in myeloid and T; β = 0.55 vs. 0.07 and 0.12, respectively). These results highlight the importance of considering cell type when studying the genetic basis of *HLA* expression.

### Conditional analysis identifies multiple independent eQTL effects in each gene

We used conditional analysis to identify additional regulatory variants beyond the primary eQTL (**Table S12**). For example, after controlling for the effect of rs3104413, a secondary independent variant (rs9272294, *r*^2^ = 0.04 with rs3104413) located ∼1.4 kb upstream of *HLA-DQA1* was also associated with *HLA-DQA1* expression in myeloid cells (*P* = 3.06 × 10^−58^, **Fig. 3C**). We repeated this process to identify up to three additional independent eQTLs (*P* < 5 × 10^−8^) for each gene in each cell type (**Figs. S8-10**). *HLA-B*, *HLA-C*, and *HLA-DQB1* exhibited the most independent signals (3 or more eQTLs per cell type). Most associations (76% = 44/58) were unique to a gene and cell type (*r*^2^ < 0.8 with all other lead variants, **Fig. S13**), but some were shared. For example, the primary eQTLs for *HLA-DPA1* and *HLA-DPB1* in B cells (rs2163472 and rs2395305, respectively) were tightly linked to each other (*r*^2^ = 1.0) and to the secondary signal for *HLA-DPB1* in T cells (rs4435981, *r*^2^ = 0.99). Additionally, the primary eQTLs for *HLA-DQA1* in myeloid and T cells (rs3104413 and rs3104371) were linked (*r*^2^ = 0.86), and the secondary signals shared the same lead variant (rs9272294).

### *HLA* genes exhibit cell-state-dependent expression

We next investigated whether *HLA* expression varies across cell states within a cell type. For each cell type (myeloid, B, or T), we integrated the single cells from all four datasets into a joint low-dimensional embedding (**Fig. 4**). To this end, we applied principal components analysis to the two tissue datasets and removed batch and dataset-specific effects using Harmony (*47*), then projected the cells from the two PBMC datasets onto the same harmonized principal component axes (hPCs) using Symphony (*48*) (**Methods, Fig. S14**).

**Fig. 4.**
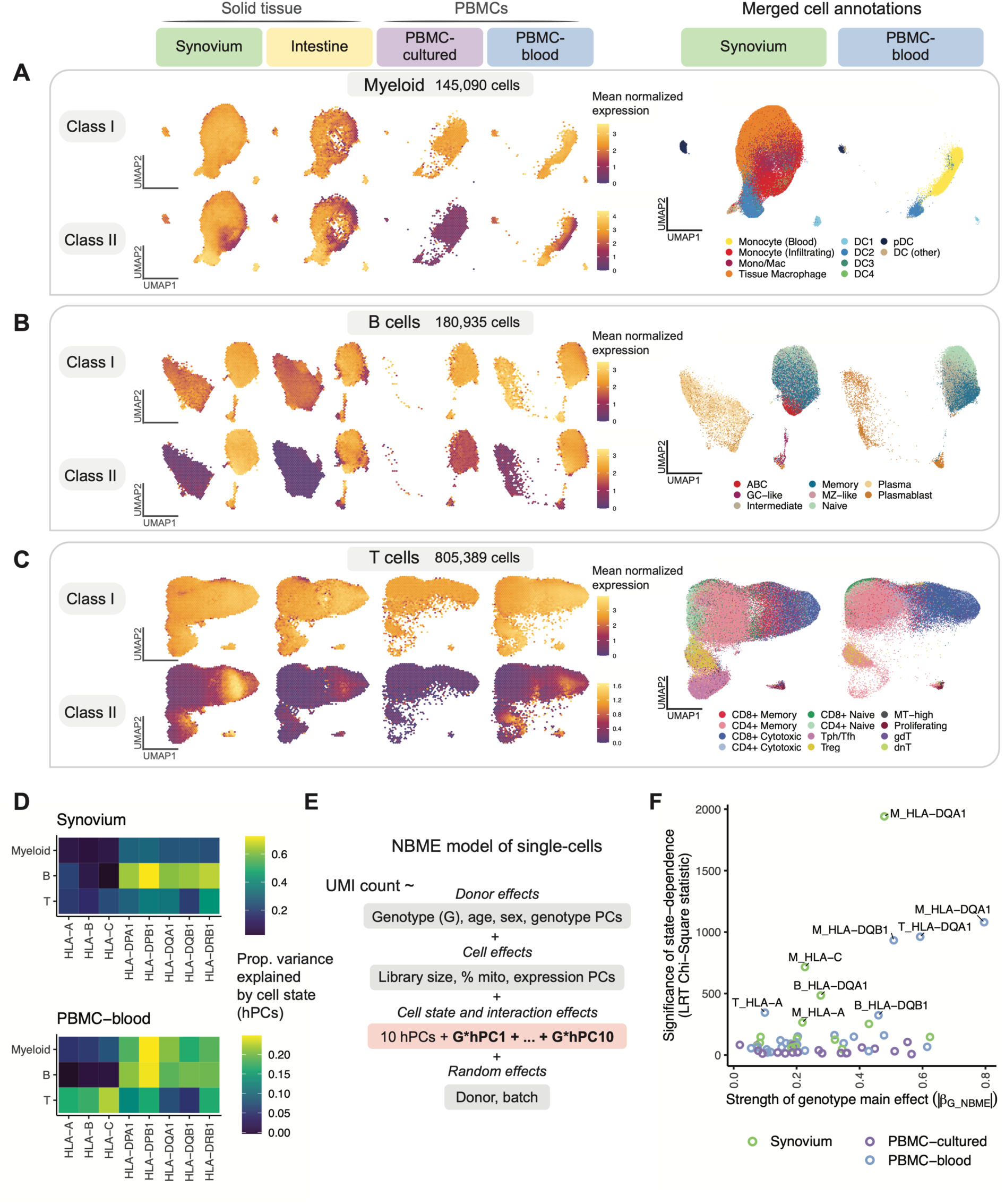
Identifying dynamic eQTLs by modeling single-cells in shared cell state embedding. (**A-C**) UMAP of cells generated using tissue-defined embedding (top 10 hPCs from Synovium and Intestine), with PBMC datasets projected into the same space. The plot is divided into three sections: (**A**) myeloid cells, (**B**) B, and (**C**) T cells. Left: class I and II *HLA* expression across cells across datasets. Cells are binned into hexagons to avoid overplotting (50 bins per horizontal and vertical UMAP directions) and colored by mean log(CP10k+1)-normalized expression of class I/II genes per bin (e.g., for class I, mean of *HLA-A*, *-B*, and *-C*). Right: cell state annotations (color) for a representative PBMC (PBMC-blood) and solid tissue (Synovium) dataset from merging annotations from each dataset to a shared set of labels. (**D**) Estimated proportion of variance in UMI counts explained by cell state hPCs (color) across *HLA* genes and cell types. (**E**) NBME model of single cells used to identify cell-state-dependent regulatory effects. Pink box highlights terms for cell state (10 hPCs per cell type) and their interaction with genotype. (**F**) Testing lead eQTLs identified in multi-cohort pseudobulk analysis for cell-state dependence using the NBME model in each dataset (color) in myeloid (“M”), B, and T cells. Magnitude of genotype main effect (*x*-axis) versus the significance of cell-state interaction (*y*-axis), measured using Chi-Square (χ²) statistic from LRT comparing full model (**E**) to null model without GxhPC interaction terms. Abbreviations: UMAP, uniform manifold approximation and projection; NBME, negative binomial mixed effects; LRT, likelihood ratio test; hPC, Harmonized PC.

The shared single-cell embedding allowed us to compare *HLA* expression patterns across fine-grained transcriptional states. Both class I and II expression varied widely across cell states within a given cell type (**Fig. 4A-C, Figs. S15-17**). By quantifying the variance explained by cell state for each gene (**Methods**), we found that cell state generally explained a greater proportion of variance in class II expression (mean 30%, 25^th^ to 75^th^ percentile [17-37%] across all cohorts) compared to class I (mean 19%, [8-34%]) (**Fig. 4D, Table S13**). The abundance of certain cell states differed considerably between blood and tissues. For example, tissue macrophages and infiltrating monocytes were absent or at low abundance in PBMCs. However, *HLA* expression patterns were generally similar in cell states shared across tissues, suggesting that cell state rather than tissue context was driving expression. For example, conventional DC1 and DC2 cells expressed the highest levels of class II among myeloid cells in both blood and tissue (**Fig. 4A**). Among B cells (**Fig. 4B**), class II expression was lower in plasma cells than in B cells, reflecting the downregulation of class II in the transition to plasma cells (*49, 50*). Among T cells, proliferating and CD8+ cytotoxic cells expressed the highest levels of class II (**Fig. 4C**).

### Modeling dynamic eQTLs at single-cell resolution

Single-cell-resolution eQTL models (*19, 20, 51*), which model expression in individual cells, can identify dynamic eQTLs—regulatory effects that change as cells transition across continuous cell states. Dynamic effects can be masked in pseudobulk analysis and may reflect cell-state-specific transcription factors binding to specific regulatory elements.

To investigate whether *HLA* eQTLs are dynamic, we used a single-cell negative binomial mixed-effects (NBME) model (**Methods**). Briefly, we modeled the UMI count of each gene as a function of genotype and its interaction with cell state, accounting for sample-level covariates (age, sex, ancestry), cell-level fixed effects (library size, percentage mitochondrial UMIs, expression PCs), and random effects for donor and batch (**Fig. 4E**). To represent a cell’s state, we used the top ten hPCs for each major cell type (**Methods**). We use this continuous multivariate representation of cell state when modeling eQTLs to capture cell state variation in a flexible and unbiased manner. We tested for cell-state interactions (GxhPC) within each dataset using the same cell-state definitions across datasets. We tested the lead eQTLs identified by our pseudobulk analysis, comprising 58 variant-gene pairs with robust genotype main effects and excluding the Intestine dataset due to its small sample size (**Methods**). We confirmed that the model has well-calibrated type I error when testing for cell-state interactions (**Fig. S18C,D**).

We observed that most eQTLs (78% = 45/58) showed statistically significant cell-state dependence (interaction *P* < 8.6 × 10^−4^ = 0.05/58 tests; **Table S14**). Indeed, every *HLA* gene tested was dynamic in at least one cell type, and *HLA-DQA1*, *HLA-DQB1*, *HLA-C*, and *HLA-A* were the most state-dependent (**Table S15**). Most interaction effects were modest relative to the main genotype effect (**Table S14**). Interestingly, the PBMC-cultured dataset exhibited much less significant cell-state interactions overall (**Fig. 4F**), despite being similar in size to the Synovium dataset. This is possibly due to cell state differences in cultured cells compared to cells collected *in vivo*.

### Comparing dynamic effects across cell states

We next assessed the strength of dynamic regulatory effects in relation to annotated cell states. For each eQTL, we calculated each cell’s estimated total eQTL effect size (*β_total_)* from the genotype main effect and interaction effects weighted by the cell’s position along each hPC (**Methods**) (*19*). This allowed us to compare the eQTL’s strength across cell states. For example, in PBMC-blood T cells, the effect of the *HLA-A* eQTL (rs7747253, interaction *P* = 4.9 × 10^−68^) was strongest in proliferating cells (mean *β_total_)*= 0.23 for proliferating vs. 0.10 for other T cells, **Fig. 5A**), suggesting the variant plays a more significant role in regulating *HLA-A* expression during T cell proliferation than at rest. The same variant was the lead *HLA-A* eQTL in myeloid cells, where it exhibited the strongest effects in specific dendritic cell subtypes (**Fig. S19A-D**).

**Fig. 5.**
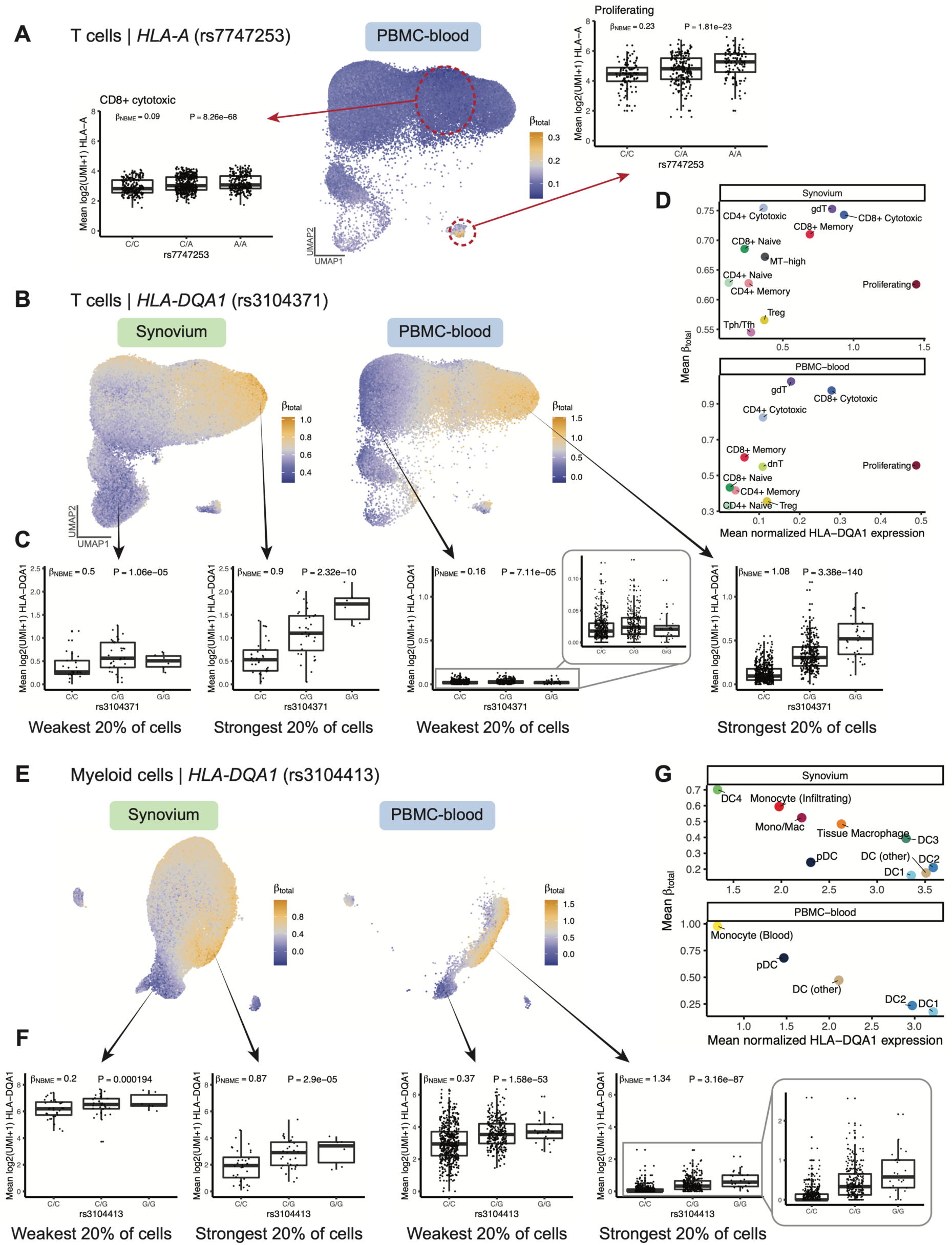
Dynamic *HLA* regulatory effects. (**A**) Dynamic *HLA-A* eQTL (rs7747253) in T cells (n=82,423 cells in Synovium, n=538,579 in PBMC-blood). UMAP shows PBMC-blood T cells colored by estimated eQTL strength (*β_total_)*). Boxplots for the eQTL effect are shown for two annotated cell states (CD8+ cytotoxic and proliferating, outlined in red circles), showing mean log2(UMI+1) of *HLA-A* across all cells in the cell state per individual by genotype. *β_NBME_)* and *P-* value values are derived from fitting the NBME model without cell state interaction terms on the discrete cell populations and comparing to a null model without genotype using an LRT (n=908 individuals, m=96,516 cells for CD8+ cytotoxic; n=409, m=739 for proliferating). (**B-D**) Dynamic *HLA-DQA1* eQTL (rs3104371) in T cells. (**B**) UMAP colored by eQTL strength (*β_total_)*), from blue (weakest) to orange (strongest). (**C**) Boxplots showing the eQTL effects in cells from the top and bottom quintile of *β_total_)*, showing mean log2(UMI+1) per individual (*y*-axis) by genotype. Labeled *β_NBME_)* and *P-*value are derived from fitting the NBME model without cell state interaction terms on the cells from the discrete quintile and comparing to a null model without genotype using an LRT. (**D**) Scatter plot showing the mean *β_total_)* (*y*-axis) compared to the mean log(CP10k+1)-normalized expression of *HLA-DQA1* (*x*-axis) across annotated cell states (color). (**E-G**) is analogous to (**B-D**), showing dynamic *HLA-DQA1* eQTL (rs3104413) in myeloid cells (n=66,789 cells in Synovium, 40,568 in PBMC-blood). For all boxplots, center line represents median; lower and upper box limits represent the 25% and 75% quantiles, respectively; whiskers extend to box limit ±1.5 × IQR; points are plotted individually.

We observed the most significant cell-state interaction effects for *HLA-DQ* genes (**Fig. 4F**), specifically *HLA-DQA1* in T cells (interaction *P* = 2.9 × 10^−200^ in PBMC-blood) and *HLA-DQA1* and *HLA-DQB1* in myeloid cells (interaction *P* < 1 × 10^−195^ in both Synovium and PBMC-blood). In T cells (**Fig. 5B-D**), the *HLA-DQA1* eQTL (rs3104371) had the strongest effects in gamma-delta (γδ), cytotoxic CD8+, and cytotoxic CD4+ T cells, a finding that replicated in Synovium (**Fig. 5D**). All three of these cell states exhibit cytotoxic activity. Our results indicate that *HLA-DQA1* expression is under dynamic genetic regulation in T cells, and further studies to clarify its functional role are warranted.

In myeloid cells, PBMC-blood and Synovium showed similar patterns of regulation for the *HLA-DQA1* eQTL (rs3104413, **Fig. 5E-G**). The strongest effects were observed in a subpopulation of monocytes in PBMC-blood and infiltrating monocytes and DC4 cells (which are similar to CD16+ monocytes (*52*)) in Synovium (**Fig. 5G**), suggesting that the underlying regulatory mechanisms governing the dynamic eQTL are active in both blood and synovial tissue. The estimated *β_total_)* values were robust to whether the embedding was defined using the tissue datasets or PBMC-blood dataset alone (Pearson *r* across cells=0.896, **Fig. S19E-G**). In contrast to the T cell *HLA-DQA1* example, the eQTL strength was negatively correlated with the expression of the gene. That is, the expression of *HLA-DQA1* is highest in conventional DC1 and DC2 cells, but the eQTL is weakest in those states (**Fig. 5G**). *HLA-DQB1* also showed similar patterns of eQTL strength as *HLA-DQA1* in PBMC-blood (*r* across cells = 0.953), suggesting that *HLA-DQ* genes are coordinately regulated.

In B cells, the *HLA-DQA1* and *HLA-DQB1* eQTLs (rs9271375 and rs927346) were also state-dependent (interaction *P* < 2 × 10^−9^ in Synovium and PBMC-blood), with plasma cells and plasmablasts exhibiting the strongest effects (**Fig. S20**). Interestingly, the overall trend in B cells was similar to myeloid cells (and opposite of T cells) in that cell states with higher *HLA-DQ* expression (pre-activated B cells and conventional DCs, respectively) had weaker eQTL effects. In contrast, states with lower expression (plasma cells and monocytes) had stronger effects. A potential explanation is that cells critical for antigen presentation, such as DCs and pre-activated B cells (*53, 54*), have mechanisms to maintain high *HLA-DQ* expression to ensure proper function, such that genetic effects contribute less to expression differences. Meanwhile, cell states with lower expression may have evolved greater genetic diversity in their antigen presentation capabilities, leading to diversity in immune responses across individuals.

## Discussion

This study demonstrates highly variable cell-type and cell-state-specific expression and genetic regulation of *HLA* genes. By integrating four diverse datasets from multiple tissues capturing a broad set of cell states and contexts, we found that classical *HLA* gene expression is under *cis*-regulation. Class II genes show particularly variable strengths of genetic regulation depending on cellular context. At the cell-type level, B cells display much stronger regulatory effects for *HLA-DRB1*, *HLA-DPA1*, and *HLA-DPB1* than myeloid and T cells (**Fig. S12**). Single-cell resolution eQTL modeling revealed that many eQTLs are cell-state-dependent, especially for *HLA-DQ* genes (**Figs. 4-5**). We previously showed that *HLA-DQ* exhibits state-dependent regulation in CD4+ T cells *ex vivo* (*18*). Here, we demonstrated that *HLA-DQ* is dynamically regulated in multiple cell types across tissues *in vivo*.

Variation in the HLA is hypothesized to have evolved to confer selective advantages in immune response to pathogens (*55*), maternal-fetal tolerance (*56*), and susceptibility to autoimmune diseases (*57*), depending on environmental contexts. Coding variation in *HLA* genes affects the quality of presented antigens by determining which peptide sequences are presented, and population diversity enables collective responsiveness to diverse pathogens. Concurrently, *HLA* regulatory variation may affect the quantity of antigen presentation, leading to different thresholds of immune responsiveness. It has been shown that the expression levels of *HLA-C* alleles can affect immunogenicity in unrelated donor hematopoietic cell transplantation (*58*), and *HLA* downregulation in tumors may affect response to immune checkpoint inhibitors (*59, 60*). The presence of multiple independent regulatory effects at each *HLA* gene and cell-type and cell-state-specific effects suggests that regulatory variation may have been selected to ensure diverse immune responses within a population.

There are several limitations of this study. First, our reference-based *HLA* imputation may have missed ultra-rare alleles. Long-read sequencing or sequence-based typing with PCR could eventually improve the detection of all possible noncoding *HLA* variants (*61, 62*). Second, we were not able to fine-map the eQTLs to precise causal variants because of the high degree of linkage disequilibrium in the MHC region. Functional work evaluating candidate causal variation may ultimately define causal variation. Additionally, we did not perform colocalization with GWAS associations because standard tools (e.g., coloc (*63*)) might not be appropriate for the MHC region because they assume a single effect, whereas coding and regulatory effects in LD may jointly contribute to disease associations.

Future data generation efforts that increase the size and ancestral diversity of genotyped single-cell cohorts will continue to improve our understanding of state-dependent and population-specific regulatory effects and aid in fine-mapping efforts (*64*).

## Data availability

For the Synovium dataset, the genotype and raw scRNA-seq data will become available upon publication of the original manuscript (*29*). For Intestine, the raw scRNA-seq data (bam files) was obtained from the Broad Data Use Oversight System (DUOS) (dataset name: Ulcerative_Colitis_in_Colon_Regev_Xavier); the genotype data will become available on dbGaP. For PBMC-cultured, the raw scRNA-seq data (FASTQ files) was obtained from GEO (PRJNA682434), and the imputed low-pass WGS data is publicly available at SRA (PRJNA736483) and Zenodo (https://doi.org/10.5281/zenodo.4273999). For PBMC-blood (OneK1K cohort), both the raw scRNA-seq data (bam files) and genotyping data are publicly available on GEO (GSE196830). The reprocessed version of all scRNA-seq count matrices from this study after realignment with scHLApers will be publicly available on Zenodo upon acceptance.

## Code availability

Code and tutorials to run the scHLApers pipeline are available on GitHub (https://github.com/immunogenomics/scHLApers). Scripts for reproducing analyses in the manuscript will become available on GitHub upon acceptance.

## Materials and Methods

### 1. Quantifying single-cell *HLA* expression with scHLApers

We developed the scHLApers (<u>s</u>ingle-<u>c</u>ell *<u>HLA</u>* expression using a <u>pers</u>onalized reference) pipeline to accurately quantify classical *HLA* expression in scRNA-seq data. As input, the pipeline takes in scRNA-seq read-level data (FASTQ or BAM) and *HLA* allele calls. If sequence-based typing is unavailable, *HLA* alleles can be imputed using genotyping data (see “HLA imputation”). A personalized reference is created for each individual by adding personalized *HLA* allele sequences as extra contigs to the reference and masking the original reference *HLA* gene sequences. The output is a whole-transcriptome counts matrix with improved *HLA* expression estimates. The code and tutorials to run scHLApers are available at https://github.com/immunogenomics/scHLApers.

#### 1.1 Preparing the *HLA* allelic sequence database

scHLApers requires a database of genomic *HLA* allele sequences. To prepare this, we downloaded the IPD-IMGT/HLA database (https://github.com/ANHIG/IMGTHLA) (v3.47.0). The database contains sequence alignment files for full-length genomic sequences (i.e., four-field resolution, ending in “gen.txt”) and nucleotide coding sequences (i.e., two-and three-field resolution, ending in “nuc.txt”). We filled in any incomplete genomic sequences with bases from the most similar complete allele using the *hla_compile_index* function from the ‘hlaseqlib’ R package (v0.0.3) (https://github.com/genevol-usp/hlaseqlib). Coding allele sequences with no corresponding genomic sequence were substituted with the genomic sequence of the most similar allele with a genomic sequence based on the Hamming distance of coding sequences. For *HLA-A, HLA-DQA1, HLA-DQB1*, *HLA-DPA1*, and *HLA-DPB1,* we padded the 5’ and 3’ ends of the allelic sequences from IPD-IMGT/HLA with extra bases from the GRCh38 reference to ensure that they did not have any missing sequence content compared to the reference sequences. The reference gene boundaries were defined by the Gencode v38 annotation file.

#### 1.2 Creating personalized reference genome and annotation files

scHLApers creates a personalized reference genome (FASTA) and annotation file (GTF) for each individual. Based on the *HLA* allele calls, scHLApers creates a FASTA file for each individual with their genomic allelic sequences from the allelic sequence database. Each allele is included as a separate contig, with the allele name as the identifier. If multiple four-field versions exist for a given two-field allele, the corresponding XX:XX:01:01 allele sequence is chosen. The original reference classical *HLA* gene sequences are masked with “NNN…” to prevent reads from aligning to them. The personalized allelic sequences are then concatenated with the masked GRCh38 reference genome to produce the personalized reference.

In the personalized annotation file (GTF), all entries corresponding to the classical *HLA* genes are removed from the original Gencode v38 annotation file. New entries are added for each personalized allele with the “seqname” column labeled as the allele name (matching the identifier in the personalized reference FASTA file), the “feature name” as “exon” to enable read alignments to the entire sequence, the “start” and “end” positions as “1” and the length of the sequence, respectively, and the strand as “+” since all sequences in the database are defined as the forward strand. The “attribute” column is labeled with “transcript_id” as the allele name (e.g., IMGT_A*01:01:01:01) and “gene_id” and “gene_name” as the gene name (e.g., IMGT_A), allowing alignments to either allele of the gene to contribute to its total UMI count.

#### 1.3 Quantifying single-cell expression

Using the personalized genome and annotations, scHLApers performs single-cell read alignment and expression quantification using STARsolo (*28*) (v2.7.10a). STARsolo performs barcode correction, UMI collapsing, and optimal distribution of multimapping reads (i.e., reads mapping to either overlapping genes or multiple paralogous genes at separate loci), which are typically discarded in standard pipelines. We chose STARsolo over pseudoalignment-to-transcriptome methods because it can identify splice junctions *de novo*, which is useful because the transcript isoform usage for each *HLA* allele is not readily available. The personalized genome index is generated using STARsolo -- runMode genomeGenerate, and read alignment is performed with --runMode alignReads. The user specifies the appropriate UMI length (--soloUMIlen), cell barcode whitelist file (--soloCBwhitelist), and assay type (--soloType CB_UMI_Simple for droplet-based data). Additionally, scHLApers counts all reads overlapping gene’s introns and exons (--soloFeatures GeneFull_Ex50pAS) and optimally distributes multimapping reads using an expectation-maximization algorithm (--soloMultiMappers EM). The parameters --soloCBmatchWLtype 1MM_Nbase_pseudocounts, --soloUMIfiltering MultiGeneUMI_CR, and --soloUMIdedup 1MM_CR are used to match CellRanger results. Users can output a coordinate-sorted BAM file to view individual read alignments (--outSAMtype BAM SortedByCoordinate and --outSAMunmapped Within).

### 2. Cohorts with paired single-cell transcriptomics and genotype data

We obtained data from four existing studies with scRNA-seq and genotype data from the same individuals (**Table S1**). These include (i) synovial biopsies from RA patients and osteoarthritis controls (Synovium) (*29*), (ii) intestinal biopsies from UC patients and healthy controls (Intestine) (*30*), (iii) PBMCs from healthy males that were treated *in vitro* with both influenza A virus and mock conditions (PBMC-cultured) (*31*), and (iv) PBMCs collected from a large population cohort (PBMC-blood) (*32*).

#### 2.1 Synovium

The original study (*29*) collected synovial biopsies from patients with RA (n=70) and control patients with osteoarthritis (n=9) as part of the Accelerating Medicines Partnership (AMP) RA/SLE Phase 2 Consortium. In this study, one RA sample was excluded due to lack of genotyping, and nine more individuals were excluded because we could not impute phased alleles for all classical *HLA* genes (see “HLA imputation”), resulting in a final cohort of 69 individuals. For three individuals with repeat biopsies, we included only initial biopsies. The original study used CITE-seq to simultaneously measure single-cell RNA and 58 protein markers, but this study uses only the RNA data. RNA libraries were generated using the 10x Genomics 3’ v3 protocol, and sample-level FASTQ files were used for input to scHLApers.

#### 2.2 Intestine

The original study included intestinal biopsies from 30 individuals (*30*). We accessed the read-level scRNA-seq data via the Broad Data Use Oversight System (https://duos.broadinstitute.org; dataset: Ulcerative_Colitis_in_Colon_Regev_Xavier). After removing five individuals without genotyping data and three individuals for whom we could not impute phased alleles for all classical *HLA* genes, the final cohort consisted of 22 individuals for this study. RNA libraries were generated using the 10x 3’ v1 protocol (n=12) or 3’ v2 protocol (n=10). Sample BAM files were used for input to scHLApers.

#### 2.3 PBMC-cultured

The original study collected whole blood from healthy male donors (n=90) of African and European ancestry (*31*). Samples from each individual were treated with influenza A virus or mock conditions (90*2 = 180 samples). Libraries were prepared using the 10x 3’ v2 protocol and multiplexed in 30 experimental batches for sequencing. We downloaded the batch-level scRNA-seq FASTQ files from GEO (GSE162632). We removed one individual with missing genotype data, one based on our IBD threshold (IBD PI_HAT > 0.9), and 15 individuals for whom we could not impute phased alleles for all classical *HLA* genes, leading to a final cohort of 73 individuals. To demultiplex the batches, we obtained a barcode-to-sample mapping file from the original authors. We used the *filterbarcodes* function from sinto (v0.8.4) and the mapping file to demultiplex the batch-level BAM file into sample-level BAM files for input to scHLApers.

#### 2.4 PBMC-blood

The OneK1K cohort (*32*) was recruited from patients and relatives at clinical sites and retirement villages in Australia. PBMCs were collected and sequenced using the 10x 3’ v2 protocol. We obtained multiplexed read-level data in BAM file format across 77 batches from the original authors. We used the *filterbarcodes* function from sinto (v0.8.4) to demultiplex each batch-level BAM file into constituent sample-level BAM files using a batch-to-sample cell barcode mapping provided by the original authors. Starting with an initial cohort of 973 individuals with paired genotyping and scRNA-seq data, we excluded three samples due to elevated missing data rates during genotype quality control (QC) and one sample missing in the barcode mapping file. We also removed 60 samples where we could not impute phased alleles for all classical *HLA* genes, leading to a final cohort of 909 samples for this study.

### 3. Quality control of genotyping data

All cohorts were genotyped using genotyping arrays, except for PBMC-cultured, which used low-pass whole-genome sequencing (WGS) (**Table S1**). We processed the genotyping data and performed QC using PLINK v1.90, as described below following the tutorial at https://github.com/immunogenomics/HLA_analyses_tutorial/blob/main/tutorial_HLAQCImputation.ipyn <u>b</u> (*24*). Genome-wide variants were used to calculate PCs to control for genetic ancestry in eQTL analysis, and variants in the extended MHC (defined here as chr 6: 28000000-34000000) were used for HLA imputation.

#### 3.1 Synovium

We genotyped donors in the AMP RA/SLE Network (including the 69 donors in this study) using the Illumina Multi-Ethnic Genotyping Array split across three batches. We lifted over the data from hg38 to hg19 coordinates, converted reverse strand variants to forward alleles with snpflip (v.0.0.6), and removed duplicated variants with a custom script. We performed initial QC to remove variants with high missing call rates or violating Hardy-Weinberg equilibrium (--geno 0.1 --hwe 1e-8) and removed 1 sample with high missingness (--mind 0.01). We then applied variant filters (--hwe 1e-6, --geno 0.01) and removed indels. We merged all three batches, removed multi-allelic variants, and removed variants with MAF <1% (--maf 0.01) and >1% missingness (--geno 0.01) across all samples. A total of 820,019 genome-wide variants (10,159 in MHC) and 788 individuals passed genotype QC.

#### 3.2 Intestine

The genotype data was obtained directly from collaborators and comprised three batches, denoted batch1 (774 individuals), batch2 (874 individuals), and batch3 (1,335 individuals). The batch1 data was generated using an Illumina Infinium Global Screening Array, and the others used a custom GWAS SNP array. Each batch was quality controlled separately. We first removed SNP duplicates, performed an initial variant filtering (--geno 0.1 --hwe 1e-10), then removed samples with high missingness (--mind 0.01) or high sample-relatedness (IBD PI_HAT > 0.9). We removed variants with >2% missingness (--geno 0.02) or >0.35 allele frequency difference from 1000 Genomes project phase 3 v.5 data. We then applied final filters (--maf 0.01 --hwe 1e-10 --mind 0.01 --geno 0.02). For batch1, 488,343 genome-wide variants (7,544 in MHC) and 765 individuals passed QC. We merged the batch2 and batch3 datasets by their 222,030 shared SNPs genome-wide (772 in MHC), and 2,121 individuals passing QC.

#### 3.3 PBMC-cultured

The imputed WGS data (VCF, n=91) was obtained directly from the original authors (data available at SRA accession PRJNA736483). To match the HLA imputation reference panel, we lifted over the variants from GRCh38 to hg19 positions using CrossMap (v0.6.1) with chain file http://hgdownload.soe.ucsc.edu/goldenPath/hg38/liftOver/hg38ToHg19.over.chain.gz. We marked genotypes with missing calls using bcftools (v1.9) if posterior genotype probabilities were <95%. We removed duplicated and multiallelic variants using PLINK v1.90 and a custom script. After initial variant QC (--geno 0.1 and –hwe 1e-10), we performed sample-level QC based on genotype missingness (--mind 0.01) and genetic relatedness using IBD comparisons on variants pruned for high LD. One sample (HMN52561) was excluded based on high genetic relatedness to another sample (HMN17122; IBD PI_HAT > 0.9). We removed variants with >2% missingness or >0.25 allele frequency difference from 1000 Genomes phase 3 v.5, resulting in 5,133,858 variants passing QC. As a final step, we subset variants to those in our HLA imputation reference panel and removed variants with MAF<5% (10,814 MHC variants and 90 individuals passing QC).

#### 3.4 PBMC-blood

The raw genotyping data in hg19 coordinates was obtained from the original authors. We used snpflip (v0.0.6) to flip 247,054 variants to the forward strand and removed 682 duplicates. We performed an initial SNP QC (--geno 0.1, --hwe 1e-10) and removed three individuals with elevated missingness rates (--mind 0.01). We removed variants with high missingness (--geno 0.02), 2,508 ambiguous (A/T or C/G) SNPs, and variants with >0.3 allele frequency difference from 1000 Genomes phase 3 v.5. After applying final filters (--maf 0.01 --hwe 1e-10 --mind 0.01 --geno 0.02), 972 individuals and 487,471 genome-wide variants (7,046 in MHC) passed QC.

### 4. HLA imputation

#### 4.1 HLA imputation with SNP2HLA

We used SNP2HLA (https://github.com/immunogenomics/HLA_analyses_tutorial/blob/main/scripts/SNP2HLA.py) to perform HLA imputation using version 2 of our group’s multi-ethnic reference panel described in Sakaue et al. (*24, 33, 34*). We performed imputation on the full genotyping datasets (i.e., not limited to samples with paired scRNA-seq), then subset the imputed VCF file to the samples with scRNA-seq. Two types of genetic variation were imputed: SNPs within the MHC (n=14,691) and classical *HLA* alleles at one-and two-field resolution for *HLA-A, -B, -C, -DPA1*, *-DPB1, -DQA1*, *-DQB1,* and *-DRB1* (n=570). In SNP2HLA output, REF and ALT values for classical alleles are set to “A” and “T” denoting absence and presence of the allele, respectively. The two-field *HLA* alleles were used in scHLApers to make personalized references. SNP2HLA outputs an individual’s imputed dosage (0–2) and inferred genotype (GT: 0|0, 0|1, 1|0, or 1|1) for every *HLA* allele in the reference panel. Note that for a subset of individuals, we could not confidently call two-field alleles for one or more *HLA* genes, and the dosage was split across multiple alleles (<0.5 for any given allele). We excluded these individuals (9 Synovium, 3 Intestine, 15 PBMC-cultured, and 60 PBMC-blood individuals, representing <8% of total samples) to avoid introducing a technical batch effect. All downstream analyses included 1,073 individuals for whom we could confidently impute phased alleles for every *HLA* gene (GT: 0|1 and 1|0 for two alleles or GT: 1|1 for one allele).

#### 4.2 Quality control of imputed MHC variants

We performed QC on the imputed MHC-wide variants using custom R scripts and the ‘vcfR’ (v1.12.0) package. Because the HLA reference uses hg19 coordinates, we first lifted over the imputed variants to GRCh38 using CrossMap (v0.6.1) and chain file http://hgdownload.soe.ucsc.edu/goldenPath/hg19/liftOver/hg19ToHg38.over.chain.gz. Then, we subset to the relevant samples and calculated the MAF within the subset. We retained variants with imputation DR2>0.8 and MAF>0.01 in each cohort. For the Intestine cohort, which was genotyped on two different arrays, we first filtered by DR2 within each array then merged them by the intersecting variants before filtering by MAF >0.01 across the merged cohort. We took the intersection of variants across all four cohorts passing our QC thresholds to arrive at a final set of 12,050 variants for eQTL testing (**Fig. S2**): 112 one-and two-field *HLA* alleles and 11,938 intergenic variants.

### 5. Assessing the performance of scHLApers

#### 5.1 Applying scHLApers to all four cohorts

We applied scHLApers to quantify single-cell expression for all four datasets. As a comparison, we also ran a standard pipeline that used STARsolo with the same parameters as scHLApers but with the original GRCh38 reference (with no personalization) and discarding multimapping reads. For both versions, we generated BAM files containing unmapped reads (samtools view -b -f 4) and reads aligning to the MHC and personalized contigs using samtools (v1.4.1). We removed empty droplets and low-quality cells by filtering the count matrices by cell barcodes (see “Processing single-cell expression data”).

For the read length concordance analysis, the PBMC-cultured dataset contained reads of two different lengths (84 and 289 bp). We generated long and short-read versions of the dataset by creating separate BAM files by sequence length and running scHLApers on longer and shorter reads separately. To visually inspect read alignments and coverage across the personalized allelic contigs in scHLApers, we used Integrative Genomics Viewer (IGV v2.11.2).

#### 5.2 Comparing percent change to dissimilarity from the reference alleles

We assessed how expression estimates (summed UMI counts across all cells for a sample) changed from a standard pipeline (sp_exp) to the scHLApers pipeline (pers_exp) (**Eq. 1**) with respect to the dissimilarity between the reference allele and personalized alleles.

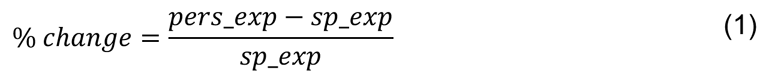

Dissimilarity was defined as the Levenshtein distance between the genomic GRCh38 allele and personalized allele sequences, calculated using the *stringdist* function in the ‘stringdist’ (v0.9.8) R package. Since all datasets used 10x 3’ assays, the read coverage was predominantly at the 3’ end of the gene (**Fig. S3**). Hence, distances were calculated at the 3’ end using sequence segments of 500 bp (*HLA-A*, *HLA-B*, *HLA-C*, *HLA-DRB1*), 1,000 bp (*HLA-DQA1*, *HLA-DPA1*), 1,500 bp (*HLA-DQB1*), or 2,500 bp (*HLA-DPB1*), encompassing the region where reads accumulated. For individuals heterozygous for a gene, we took the mean of the two distances. The GRCh38 reference allele sequences are listed in Darby et al. (*65*) (*A*03:01*, *B*07:02*, *C*07:02*, *DQA1*01:02*, *DQB1*06:02*, *DRB1*15:01*, *DPA1*01:03*, *DPB1*04:01*). We confirmed these by performing a multiple sequence alignment between the IPD-IMGT/HLA allelic sequences and the reference sequence using the *msaClustalW* function from the ‘msa’ (v1.22.0) R package.

#### 5.3 Investigating read mapping between *HLA-B* and *HLA-C*

To quantify the rescuing of unmapped reads and identify reads “jumping” between different genes, we tracked where reads aligned in scHLApers versus the standard pipeline. We analyzed the BAM files output from both pipelines using a custom R script and the *scanbam* function in ‘Rsamtools’ (v2.6.0). A given read can align to the classical *HLA* genes (i.e., personalized contigs for scHLApers or gene regions defined by Gencode v38 for the standard pipeline), another location in the MHC outside of classical *HLA* genes, another location outside of the MHC, or be unmapped. We used the multiple sequence alignment for *HLA-C* to generate a phylogenetic tree of *HLA-C* allele sequences using the “Neighbor Joining” option in Jalview (v2.11.0). By grouping the *HLA-C* alleles by similarity to the reference allele (*C*07:02*) based on the tree, we could observe the relationship between the dosage of “reference-like” *HLA-C* alleles and the change in *HLA-B* counts.

### 6. Processing single-cell expression data

#### 6.1 Quality control of single-cell data

For Synovium, Intestine, and PBMC-cultured datasets, we subset the count matrix output from scHLApers to the cells passing QC in the original studies (i.e., barcodes present in published cell metadata). For the PBMC-blood dataset, we started from the original cells but performed additional filtering steps to remove suspected doublets (see **Supplementary Text 2**). Then, we performed uniform cell-level quality control procedures on all cohorts, removing cells with <500 genes and >20% mitochondrial counts.

#### 6.2 Defining major cell types and merged cell annotations

We defined a common set of six major cell types across the four datasets: myeloid (monocytes, macrophages, and DCs), B (including plasma), T, NK, fibroblast, and endothelial by aggregating fine-grained cell annotations. For Synovium, Intestine, and PBMC-cultured, these fine-grained annotations came from the originally published cell annotations. For PBMC-blood, we used the Seurat Azimuth PBMC CITE-seq reference (https://app.azimuth.hubmapconsortium.org/app/human-pbmc) to transfer labels to the cells following the more stringent doublet removal (**Supplementary Text 2**). We removed cells from the following annotations that did not fall under our major cell type categories of interest: “Mu-0: Mural” and “T-21: Innate-like” cells in Synovium; “Glia,” “CD69-Mast,” “CD69+ Mast,” and “Pericytes” for Intestine; “NKT” and “neutrophils” for Intestine; and “HSPC,” “Platelet,” “Doublet,” “Eryth,” and “MAIT” for PBMC-blood. The final cell numbers can be found in **Table S2**. We generated cell-type-specific count matrices for downstream analyses, removing cells from individuals with fewer than five cells of the cell type. To obtain a version of finer-grained cell annotations to aid in the interpretation of cell embeddings, we manually merged the fine-grained cell annotations for myeloid, B, and T cells in Synovium and PBMC-blood datasets to a shared set of common cell state labels (e.g., PBMC-blood “CD4 CTL” and “CD4 TEM” and Synovium “T-12: CD4+ GNLY+” were merged into “CD4+ Cytotoxic”, see **Table S5**).

### 7. Pseudobulk eQTL analysis

#### 7.1 Generation of pseudobulk profiles

For each cell type (myeloid, B, and T), we generated “pseudobulk” versions for each dataset. First, we performed library size normalization using log(CP10k+1) within each cell, then aggregated all cells per sample by taking the mean normalized expression of each gene to obtain a samples-by-genes matrix (*66*). We performed rank-based inverse normal transformation for each gene, including genes with nonzero expression in greater than half of the samples.

#### 7.2 Multi-cohort eQTL model

To control for genetic ancestry, we used PLINK (v1.90) to calculate genotype PCs (gPCs) using 66,827 shared genome-wide variants across all four datasets. For PCA, we included all individuals from the full array cohorts passing QC (including those without paired scRNA-seq data, **Fig. S1F**). To infer hidden determinants of gene expression variation, we ran probabilistic estimation of expression residuals (PEER) (*67*) on each pseudobulk expression matrix for each dataset and cell type separately, using the ‘peer’ R package (v1.0). We used different numbers of PEER factors (K) for each dataset to account for the varying number of individuals in each cohort (K=7 for Synovium, 2 for Intestine, 7 for PBMC-cultured, and 20 for PBMC-blood, **Fig. S7A**). We generated covariate-corrected expression residuals, accounting for sex, age, ancestry (5 gPCs), 10x chemistry (for Intestine), and PEER factors.

To identify eQTLs for each classical *HLA* gene, we incorporated all four datasets into a single model (“multi-cohort model”) to boost power. We combined the expression residuals from all datasets together for each cell type (**Fig. S7B**). For PBMC-cultured, which included both influenza-stimulated and noninfected cells for each sample, we included only the noninfected cells in the analysis. We tested each of the 12,050 MHC-wide variants for association with residualized expression (*E_resid_*) using linear regression (**Eq. 2**), controlling for the dataset to account for systematic differences across cohorts. This provided a pooled estimate for each eQTL effect across datasets. For lead eQTLs in the multi-cohort model, we also ran the model in each dataset separately (without the dataset term) to compare the concordance across datasets.

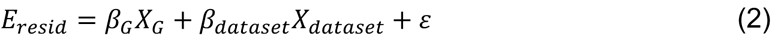

#### 7.3 Comparison to Aguiar et al. bulk eQTL study

We compared the lead eQTL effects identified in this study to a bulk RNA-seq study by Aguiar et al. (*17*) on *HLA* eQTLs in LCLs. We obtained eQTL summary statistics from the original authors and limited the comparison to B cells in this study as they are most biologically similar to LCLs.

Because some variants tested in this study were not tested in Aguiar et al., we restricted the comparison to the lead variants among those tested in both.

#### 7.4 Grouping classical *HLA* alleles by lead eQTL variants

To determine how classical one-and two-field *HLA* alleles track with lead eQTL variants, we compared the co-occurrence between eQTL variants and *HLA* alleles for the associated gene. To calculate co-occurrence (*Occ_allele,eQTL_*, ranging from 0 to 1), we used the multi-ethnic HLA reference panel dataset from HLA imputation (*24*). Because the reference dataset is phased, we could calculate the proportion of reference haplotypes (n = 20,349 samples * 2 chromosomes = 40,698 haplotypes) containing the ALT allele of each lead eQTL using a custom R script (**Eq. 3**).

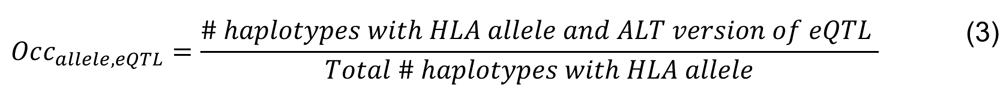

#### 7.5 Cell-type interaction analysis

To determine whether lead eQTLs are cell-type-dependent, we modeled the residualized expression from all three cell types together using a linear mixed-effects model, adding a fixed effect for cell type (myeloid, B, or T), an interaction term between variant and cell type (Gxcell_type), and a random effect for donor to account for the non-independent sampling of cell types from the same donor (**Eq. 4**). To ascertain the significance of the cell-type-dependency, we compared the full model to a null model without the interaction term using a likelihood ratio test (LRT) (*lrtest* function from ‘lmtest’ v0.9-39 R package).

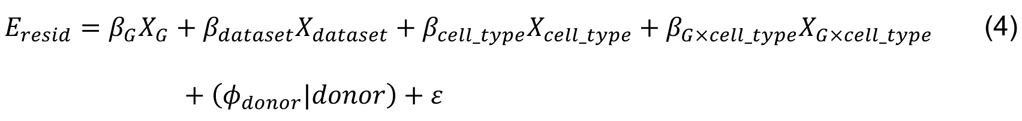

#### 7.6 Conditional analysis

To identify additional eQTLs independent from the lead eQTL, we performed up to three additional rounds of conditional analysis for each gene and cell type using the multi-cohort model, conditioning on the lead eQTL(s) from the previous round(s). We terminated early if the lead eQTL did not reach a significance of *P*<5e-8. We used PLINK (v1.90) (--ld) to calculate LD *r^2^* values between every pair of lead eQTLs across cell types and rounds of conditional analysis using the multi-ethnic HLA reference panel.

#### 7.7 Visualizations

To generate boxplots of pseudobulk eQTL effects, we used the expression residuals and regressed out the effect of dataset (not already corrected during PEER). For the Manhattan plots, because each gene has multiple potential transcription start sites (TSS) depending on the transcript, we selected the transcript with the midpoint chromosomal start position across transcripts. LD *r^2^* values for the locus zoom plot were calculated using PLINK (v.1.90) and the multi-ethnic HLA reference panel.

### 8. Creating a single-cell atlas of *HLA* expression

#### 8.1 Mapping cells into a shared embedding

To create low-dimensional cell state embeddings of single-cells across datasets, we first integrated the two tissue datasets (Synovium and Intestine). For each cell type (myeloid, B, and T), we concatenated the counts matrices from both datasets and filtered to the union of the top 1,500 variable genes per dataset calculated using the variance stabilizing transform (vst) method, excluding cell cycle genes (Seurat v4.1.0 s.genes and g2m.genes), mitochondrial (MT-), and ribosomal (RPL-, RPS-) genes. We scaled the variable genes across all cells, calculated the top 10 PCs (using the ‘irlba’ v2.3.5 R package), then removed sample and dataset-specific effects using Harmony (*47*) (v0.1.0) (parameters: V_sample_ = 0.5, V_dataset_ = 1, nclust = 50, and sigma = 0.2), resulting in a 10-dimensional “Harmonized PC” (hPC) embedding. We visualized the embedding in 2D using UMAP, calculated with the *umap* function in the ‘uwot’ (v0.1.11) R package, with n_neighbors = 30 and min_dist = 0.2. We then projected the two PBMC datasets into the same tissue-defined embedding using Symphony (*48*) (v0.1.0) to align analogous cell states across tissues. For PBMC-cultured, we included cells from both influenza-stimulated and noninfected samples. Symphony mapping was performed one query dataset at a time, correcting for ‘sample’ effects in the query.

As an alternative approach, we also explored *de novo* integration of all four datasets together. We used the top 1,500 variable genes per dataset (top 1,000 for T cells) and Harmony integration with V_dataset_ = 0.5, V_batch_ = 0.5, and V_sample_ = 0.5 (batch defined as the sample for Synovium, 10x chemistry for Intestine, and experimental batch for PBMC datasets). However, the tissue-defined embeddings produced a cleaner visual separation of cell states, particularly for myeloid cells (**Fig. S14**) and were therefore used for downstream analysis.

#### 8.2 Quantifying proportion of expression variance explained by cell state

To estimate the percent of variance in *HLA* expression explained by cell state, we fit a negative binomial mixed-effects (NBME) model of the UMI count of each *HLA* gene across cells in each cell type. We included donor-level fixed effects for age, sex, and ancestry (5 gPCs), cell-level fixed effects for scaled log(total UMI count), scaled percent mitochondrial UMIs, and cell state (10 hPCs), and random effects for donor (and experimental batch for PBMC datasets). The NBME models (including all other versions described in subsequent sections) were fit using the *glmer.nb* function from the ‘lme4’ (v1.1-28) R package with options nAGQ = 0 and “nloptwrap” optimizer. We used the *r.squaredGLMM* function from the ‘MuMIn’ (v.1.43.17) R package (*68*) to estimate the marginal R^2^ using the ‘delta’ method for the full model (**Eq. 5**) as well as a model without cell state terms. The difference between the R^2^ values between the two models was used to estimate the proportion of variance explained by cell state.

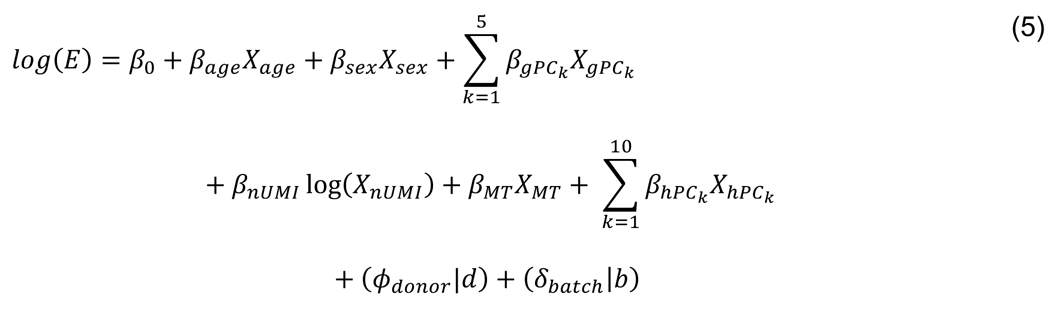

#### 8.3 Defining a cell embedding using PBMC-blood alone

We also defined an alternative cell state embedding for each cell type using cells from PBMC-blood alone. To do this, we used the same dimensionality reduction pipeline described above for the tissue-defined embedding, except we used the top 2,000 variable genes across PBMC-blood for each cell type and corrected for experimental batch with Harmony (V_batch_ = 2).

### 9. Single-cell eQTL analysis

We used a single-cell NBME eQTL model to test *HLA* eQTLs for cell-state dependency. The model is adapted from the Poisson mixed-effects (PME) model recently described by our group (*19*). We used NBME in this study because we found that the LRT *P-*values from the PME model exhibited inflation when testing for cell-state interactions (**Fig. S18C**; see “Evaluating model calibration for testing cell-state interaction”), likely because *HLA* genes exhibit greater overdispersion than other genes, whereas NBME was well-calibrated. We first used an NBME model without cell state to define the set of variant-gene pairs with robust genotype main effects within each dataset. We then used an NBME model with cell state to test for dynamic effects. We excluded the Intestine dataset due to small sample size (n=22).

#### 9.1 Testing for genotype effect using NBME model without cell state

Using the lead eQTL variants identified in the pseudobulk multi-cohort model above (8 genes * 3 cell types = 24 variants), we tested each eQTL using a single-cell NBME model (**Eq. 6**) to assess the genotype effect. We modeled the per-cell UMI count of each *HLA* gene in each major cell type and dataset separately (24 variants * 3 datasets = 72 variant-gene pairs to test). We included the same donor and cell-level fixed and random effects as in **Eq. 5**, except without cell state terms (hPCs) and adding additional terms for donor genotype (G) and five expression PCs (ePCs), which are calculated on each dataset separately to account for technical effects (akin to PEER factors in pseudobulk). We determined the significance of the genotype effect by comparing to a null model without genotype using an LRT with 1 degree of freedom.

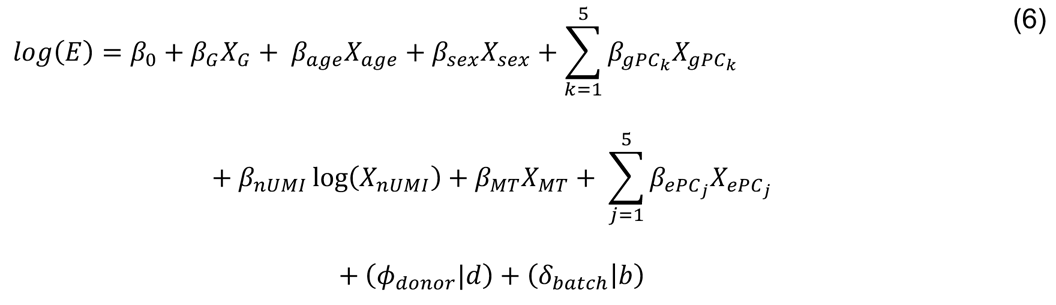

We compared the genotype main effect size and significance from the NBME model (**Eq. 6**) to the pseudobulk eQTL model using the PBMC-blood dataset. Significance was represented by LRT *P*-values in the NBME model and Wald *P*-values in the pseudobulk linear model (run on PBMC-blood separately).

To define variants with robust main effects to test for cell-state interaction, we included only variant-gene pairs within a cell type and dataset with a significant genotype main effect (LRT *P-*value < 0.05), resulting in a total of 58 variant-gene pairs.

#### 9.2 Testing for cell state interaction using NBME model

To test the 58 variant-gene pairs for dynamic regulatory effects, we modeled the eQTLs at single-cell resolution using an NBME model (**Eq. 7**). While the model can use any cell state variable (e.g., clusters, pseudotime trajectory), we reasoned that hPCs would provide a principled and unbiased way to define continuous cell states. We include the same donor and cell-level fixed and random effects as in **Eq. 6**, with the addition of cell state (hPC1-10 from the tissue-defined Symphony embeddings) and genotype interaction with cell state (GxhPC1 + … + GxhPC10). To assess whether the eQTL is cell-state-dependent, we compared the full model (**Eq. 7**) to a null model without interaction terms using an LRT with 10 degrees of freedom.

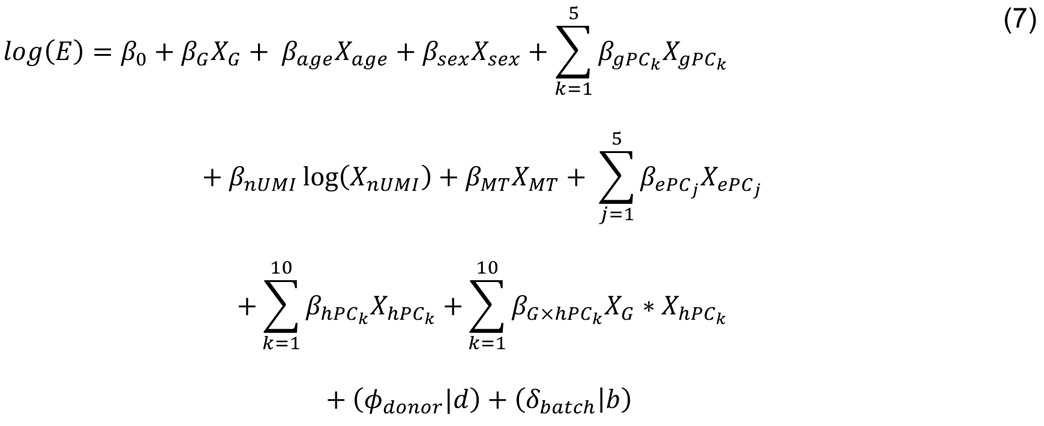

#### 9.3 Evaluating model calibration for testing cell-state interaction

We analyzed the calibration of the NBME model when testing for interaction between genotype and cell state. Using the PBMC-blood cells and embedding defined in PBMC-blood alone, we permuted cell state (10 hPCs as a block) across all cells, then ran the NBME model for each variant-gene pair (**Eq. 7**) and assessed its significance using LRT, which should yield uniform *P*-values if the model is well-calibrated. We repeated this process for 1,000 permutations and compared the results to the equivalent analysis performed with a PME model (*glmer* function from ‘lme4’ R package with family = ‘poisson’).

#### 9.4 Comparing eQTL strength across cell states

For a given eQTL, we combined the genotype main effect (*β_G_*) with the interaction effects of each hPC (estimated in **Eq. 7**), weighted by each cell’s position along each hPC (e.g., *β_G×hPC1_ × hPC1*) to score each cell based on its estimated total eQTL effect size (**Eq. 8**). This allowed us to compare the strength of the eQTL across cell states by plotting the estimated *β_total_)* of each cell in UMAP coordinates and comparing the mean *β_total_)* across cell state annotations.

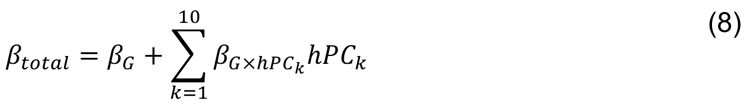

By binning cells by 5 quantiles of estimated *β_total_)*, we calculated the main genotype effect in each quantile separately (*β_NBME_)*) using **Eq. 6**, determining significance by LRT comparing to a null model without the genotype term.

To compare the *β_total_)* estimates derived from the tissue-defined embedding to those from the embedding defined using PBMC-blood alone for the myeloid *HLA-DQA1* eQTL (rs3104413), we ran the same NBME cell-state interaction model (**Eq. 7**) except using 10 hPCs defined in PBMC-blood (see “Defining a cell embedding using PBMC-blood alone”). We calculated the Pearson correlation between the *β_total_)* estimates produced by the two embeddings.

## Data Availability

For the Synovium dataset, the genotype and raw scRNA-seq data will become available upon publication of the original manuscript (Zhang et al., bioRxiv, 2022). For Intestine, the raw scRNA-seq data (bam files) was obtained from the Broad Data Use Oversight System (DUOS) (dataset name: Ulcerative_Colitis_in_Colon_Regev_Xavier); the genotype data will become available on dbGaP. For PBMC-cultured, the raw scRNA-seq data (FASTQ files) was obtained from GEO (PRJNA682434), and the imputed low-pass WGS data is publicly available at SRA (PRJNA736483) and Zenodo (https://doi.org/10.5281/zenodo.4273999). For PBMC-blood (OneK1K cohort), both the raw scRNA-seq data (bam files) and genotyping data are publicly available on GEO (GSE196830).

## Acknowledgments

We would like to thank Alex Dobin, Haley Randolph, Helena Lau, Christine Stevens, and members of the Raychaudhuri Lab, in particular Anika Gupta and Yuriy Baglaenko, for their helpful input and discussions.

## Author contributions

Conceptualization: J.B.K., S.R.

Methodology: J.B.K., A.Z.S., S.S., Y.L., S.G., S.R.

Formal analysis: J.B.K., A.Z.S., S.S., Y.L., S.G., L.R.

Investigation: A.N., V.R.C.A., C.V., K.L., M.G.-A.

Resources: F.Z., A.H.J., S.Y., J.A.-H., H.K., A.N.A., K.J., K.D., AMP RA/SLE, M.J.D., R.J.X., L.D., J.H.A., J.E.P., D.A.R., M.B.B.

Supervision: S.R.

Writing – original draft: J.B.K., S.R. Writing – review & editing: all authors

