## Supplementary Information for "Mapping the dynamic genetic regulatory architecture of *HLA* genes at single-cell resolution"

**Kang et al.**

**The PDF file includes:**

Supplementary Texts 1 and 2
Figs. S1 to S20
Tables S1 to S15

### Supplementary Text 1

This section contains additional results from assessing the performance of the scHLApers pipeline.

#### Examining reads switching alignments from *HLA-B* to *HLA-C*

Given the shared evolutionary history of class I genes (*69*), we hypothesized that the observed decrease in *HLA-B* expression after personalization was due to reads aligned to *HLA-B* in the standard pipeline aligning to a different gene in scHLApers. By tracking where individual reads aligned before and after personalization for Synovium and PBMC-cultured using the BAM files generated by STARsolo, we found that in both datasets, 99% of reads that previously aligned to *HLA-B* (but aligned to a different location after personalization) aligned instead to *HLA-C*. We then tracked where the read alignments to *HLA-C* in scHLApers "came from" in the standard pipeline (**Fig. S5A**). For Synovium, 14.8% came from *HLA-B* in the standard pipeline, 75.1% were originally also aligned to *HLA-C* in the standard pipeline, 8.3% came from unmapped reads, and the remaining 1.8% came from other genomic regions. For PBMC-cultured, the breakdown was 2.5% *HLA-B*, 51.7% *HLA-C*, 44.4% unmapped, and 1.4% other.

Interestingly, an individual's change in *HLA-B* counts depended on their *HLA-C* genotype, supporting the observed decrease in *HLA-B* after personalization. Performing a multiple sequence alignment on the *HLA-C* alleles present in our cohorts showed that the reference allele (*HLA-C*07:02*) grouped with a set of similar "reference-like" alleles (**Fig. S5B**): *HLA-C*04:04*, *C*04:01*, *C*18:01*, *C*14:02*, *C*14:03*, *C*01:02*, *C*03:04*, *C*03:03*, *C*03:05*, *C*03:02*, *C*17:01*, *C*07:01*, and *C*07:04*. For individuals with both *HLA-C* alleles similar to the reference allele (*HLA-C*07:02*), *HLA-B* was less affected by personalization (**Fig. S5B,C**). However, for individuals with at least one "non-reference-like" *HLA-C* allele (i.e., other than *HLA-C*07:02*), some reads aligned to *HLA-B* before personalization aligned better to *HLA-C* after personalization, leading to decreased *HLA-B* counts.

#### Application of scHLApers pipeline to 10x 5'-based dataset

All four datasets included in the study used 10x 3'-based single-cell libraries. As a proof-of-concept analysis demonstrating the feasibility of scHLApers on 5'-based data, we also applied scHLApers to a separate 10x 5'-based dataset from a subset of Synovium individuals (n=9 individuals, 26,638 cells, see **Fig. S4B**). We found that in 5'-based data, estimates for all eight classical *HLA* genes increased after personalization.

#### Technical note on *HLA* allele calling and allele-specific expression

scHLApers requires *HLA* allele calls per individual, which can be obtained directly by sequence-based typing or by HLA imputation using genotyped variants. There have been efforts to use bulk RNA-seq to infer *HLA* alleles without orthogonal genotype data (*17, 70*); however, inferring alleles from single-cell reads with high accuracy may prove challenging beyond one-field resolution (*71*). Allele-specific expression (ASE) analysis is an alternative way to detect regulatory effects. However, it is challenging to map reads unambiguously between alleles using short-read 3'-based sequencing data because it largely excludes the highly variable 5' region of the gene (**Fig. S3**). In contrast, 5'-based data may be more effective for ASE.

### Supplementary Text 2

See the separate document (Kang_etal_SuppText_2.pdf, appended to the end of this PDF), which contains additional methods regarding the removal of suspected doublets for the PBMC-blood dataset (OneK1K cohort). We present these methods as a separate supplementary text to allow it to serve as a standalone entity and be referenced by subsequent work.

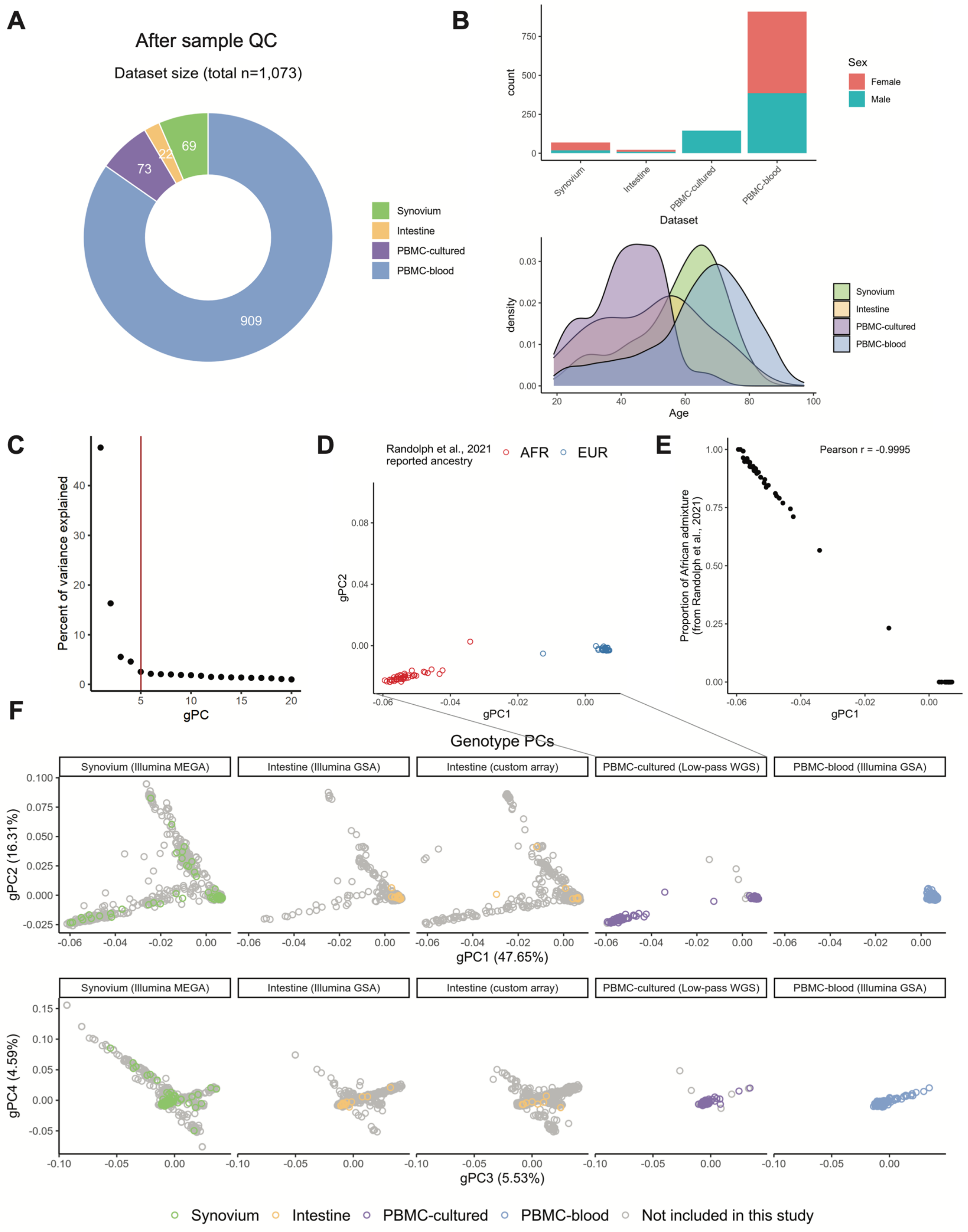

**Fig. S1. Cohort demographics.** (**A**) Cohort sizes after QC. (**B**) Sex and age distributions. Note: PBMC-cultured is male-only. (**C-F**) Genotype PCs (gPC) capturing genetic ancestry, calculated on the intersecting genome-wide variants across all cohorts. (**C**) Percentage of variance explained by each gPC. Red line denotes five gPCs used in the eQTL analysis. (**D**) Reported ancestry (African = red, European = blue) of PBMC-cultured individuals. (**E**) PBMC-cultured individuals along gPC1 (*x*-axis) versus estimated proportion of African admixture (as reported by original study, *y*-axis). (**F**) Top four gPCs across individuals. Colors denote individuals included in the eQTL analysis, whereas gray individuals were genotyped on the same array (used in PCA) but did not have available scRNA-seq data. Note: all PBMC-blood individuals are European ancestry; Intestine cohort was genotyped across two arrays.

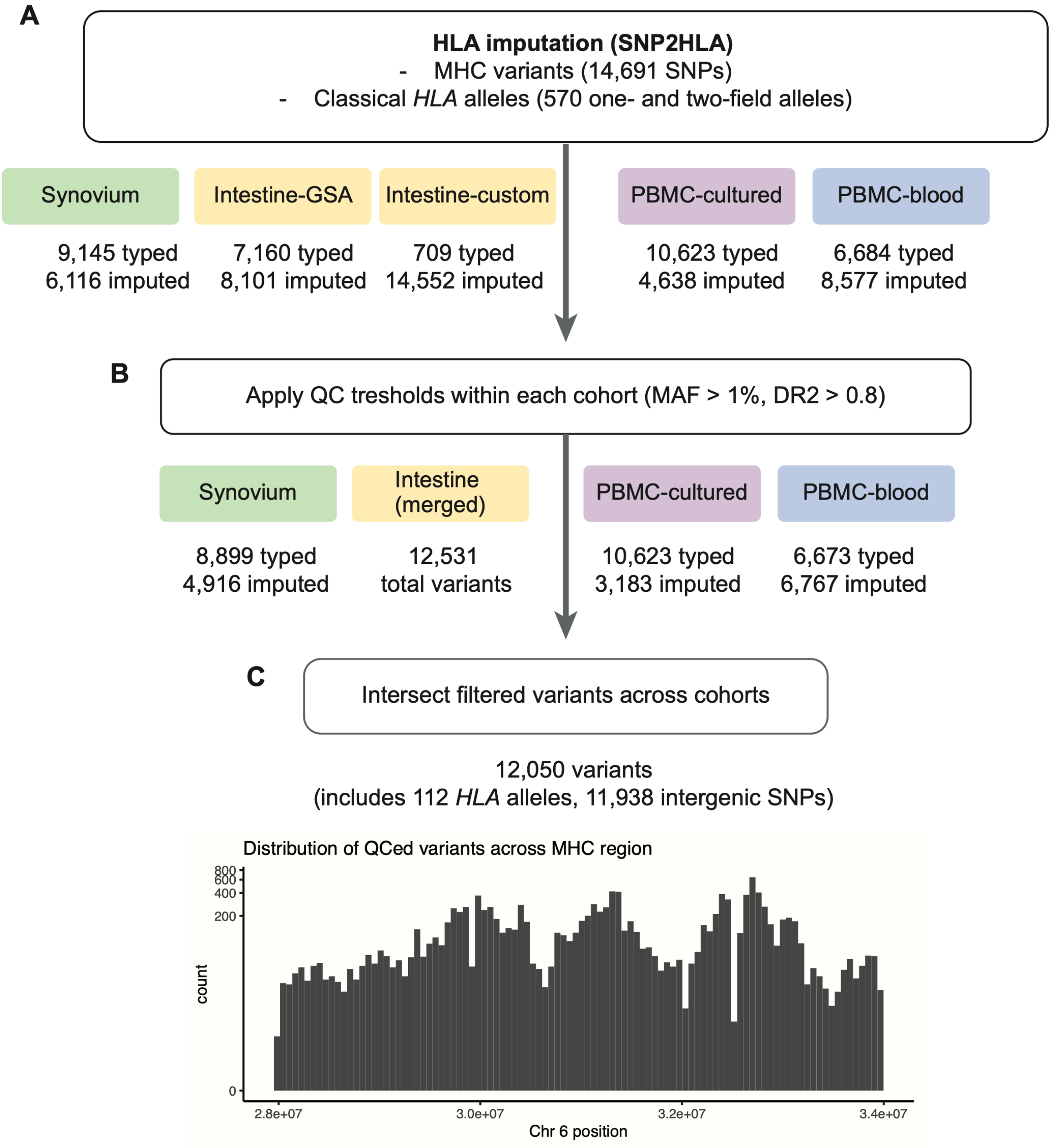

**Fig. S2. Imputation and quality control of MHC variants for eQTL analysis.** (**A**) The number of starting typed and imputed MHC variants in each cohort; the Intestine dataset was genotyped on two arrays. (**B**) The number of variants remaining after filtering for MAF > 1% and DR2 > 0.8 within each cohort separately. (**C**) The final number of variants used in eQTL analysis after taking the intersection of variants passing QC across cohorts. The histogram shows the distribution of variants across the MHC region (*x*-axis).

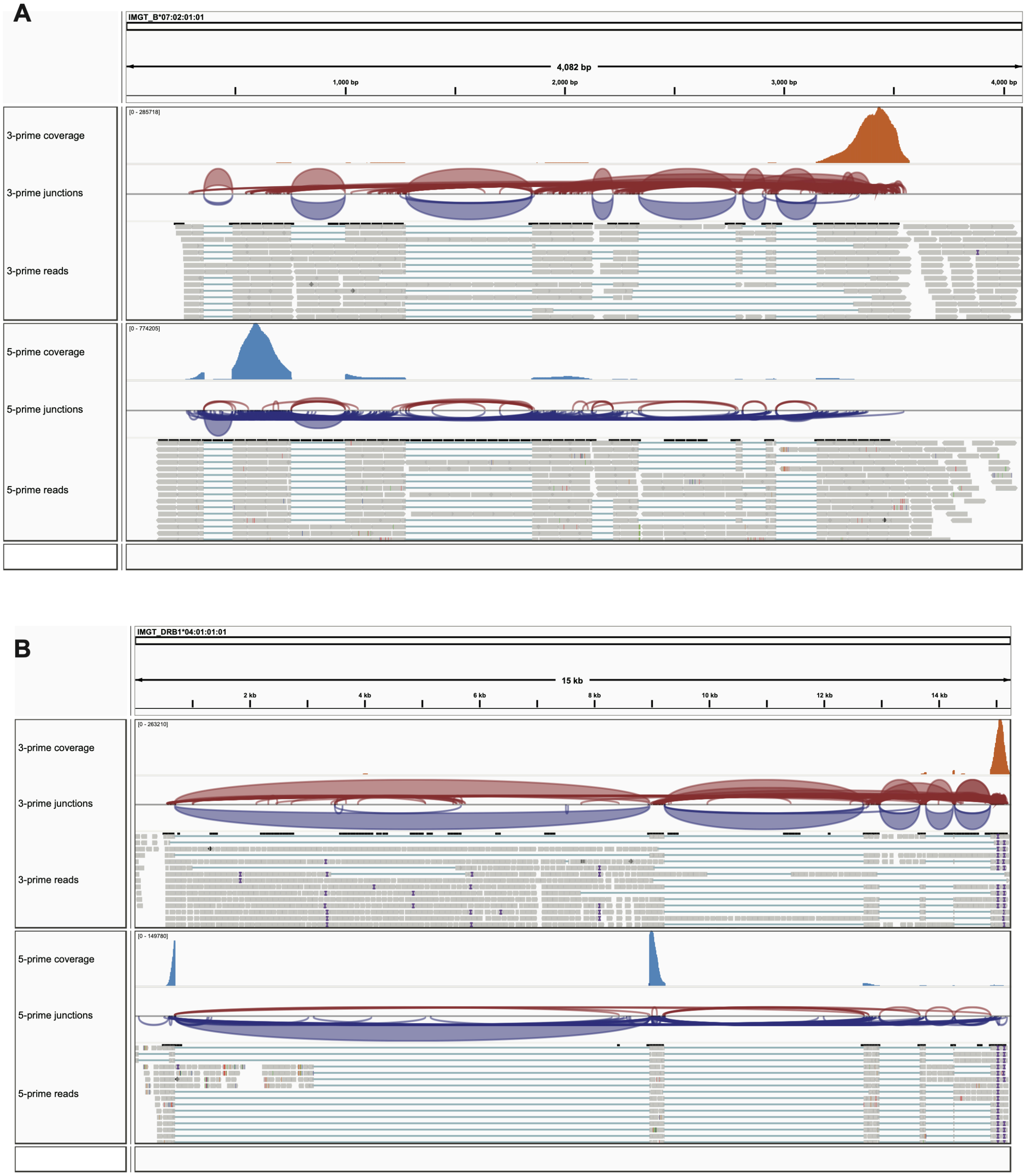

**Fig. S3. Read coverage for 10x scRNA-seq 3' and 5' assays.** (**A**) Integrative Genomics Viewer (IGV) screenshot showing read alignments from scHLApers pipeline to a representative class I allele for two samples (from the same Synovium individual), sequenced with 10x 3' assay (top, orange track) and 5' assay (bottom, blue). Additional tracks show inferred splice junctions and example individual read alignments. (**B**) Same as in (**A**) except shows alignments to a representative class II allele.

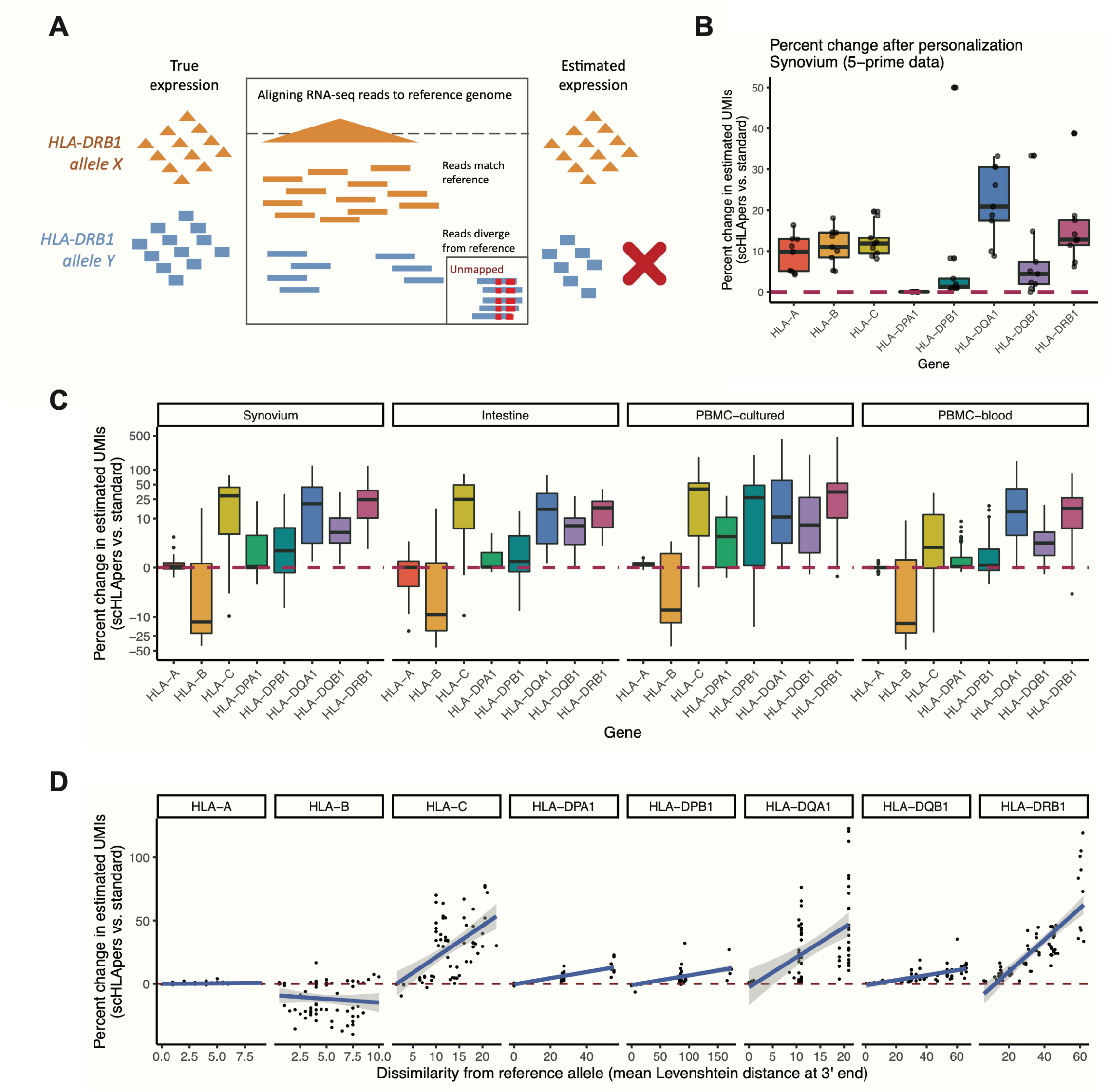

**Fig. S4. Correcting *HLA* expression estimation bias with scHLApers.** (**A**) Schematic showing how high *HLA* gene polymorphism leads to bias in read alignment to a single reference genome. Consider two hypothetical individuals who are either homozygous for *HLA-DRB1* allele X (orange) or allele Y (green), where the reference allele is X. Reads from X will align perfectly to the reference, leading to accurate *HLA-DRB1* quantification. However, for Y, reads will fail to align to the reference due to discordant sequence content, leading to unmapped reads and underestimation of expression. (**B**) Percentage change per individual (*y*-axis) in total UMIs for each *HLA* gene (*x*-axis) across all cells after scHLApers (compared to standard pipeline) in 10x 5'-based data (n=9 individuals, 26,638 cells, subset of main Synovium cohort). (**C**) Percentage change in expression (total UMIs for *HLA* gene per individual, *y*-axis) across cohorts (Synovium n=69 individuals, Intestine n=22, PBMC-cultured n=73, PBMC-blood n = 909). (**D**) Percentage change in estimated expression (total UMIs for *HLA* gene per individual, *y*-axis) in Synovium (n=69) as a function of the mean (between the individual's two alleles) Levenshtein distance relative to the GRCh38 reference allele at the 3' end of each gene (*x*-axis). For (**B-D**), dotted red lines denote no change. For all boxplots, center line represents median; lower and upper box limits represent the 25% and 75% quantiles, respectively; whiskers extend to box limit ±1.5 × IQR; outlying points are plotted individually.

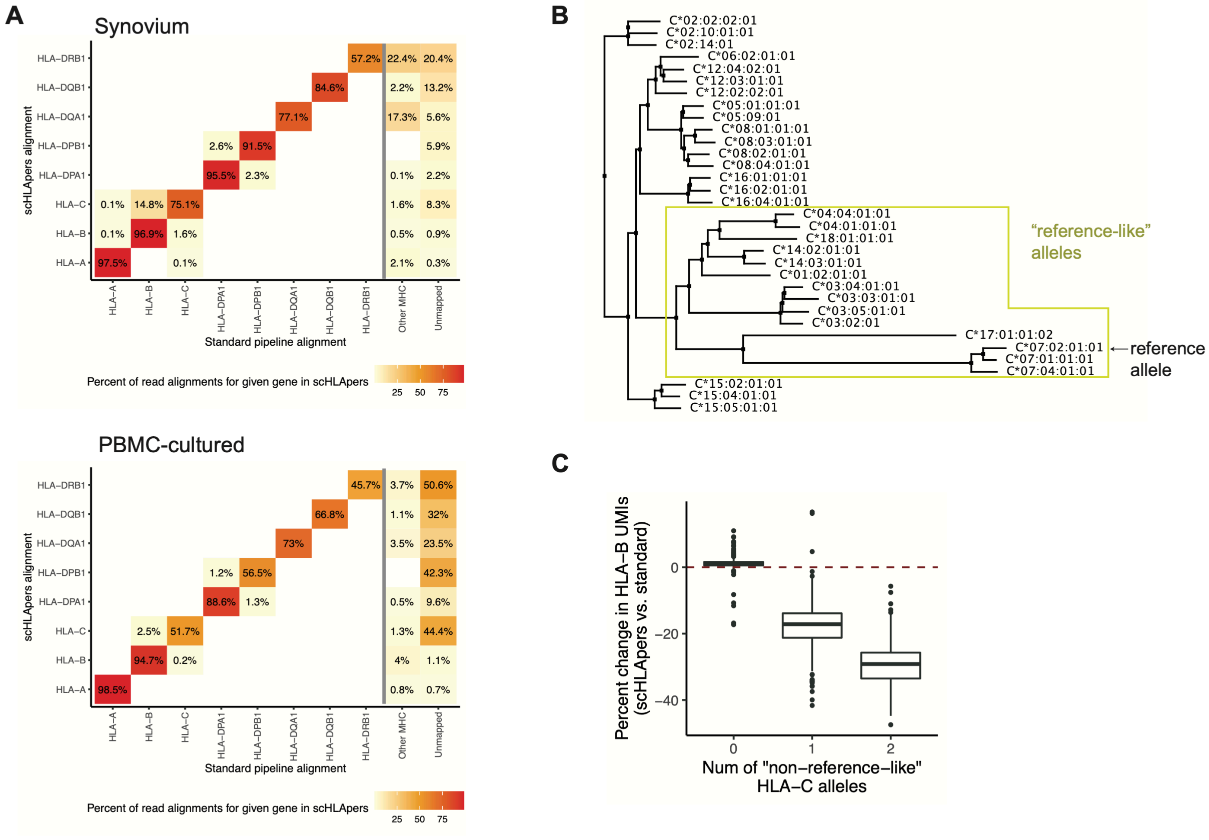

**Fig. S5. Reads switching alignments between *HLA-B* and *HLA-C*.** (**A**) Heatmap showing the alignment of reads to each gene in scHLApers (rows) versus where the same read aligned ("came from") in the standard pipeline (columns) for Synovium (top) and PBMC-cultured (bottom). Columns include *HLA* genes, other regions in the extended MHC, or unmapped reads. Rows sum to 100%, and a darker color indicates that more of the reads aligning to a given gene in scHLApers came from the corresponding location in the standard pipeline. (**B**) Phylogenetic tree derived from a multiple sequence alignment of *HLA-C* allelic genomic sequences. The reference allele is *C*07:02*. Yellow box shows alleles similar to the reference ("reference-like"). (**C**) Boxplot showing the change in *HLA-B* estimated UMI counts summed across cells from each sample (*y*-axis) compared to the genotype for *HLA-C* in terms of dosage of "non-reference-like" alleles (*x*-axis). The center line of the boxplot represents the median, the lower and upper box limits represent the 25% and 75% quantiles, respectively, the whiskers extend to the box limit ±1.5 × IQR, and outlying points are plotted individually. Dotted red line denotes no change.

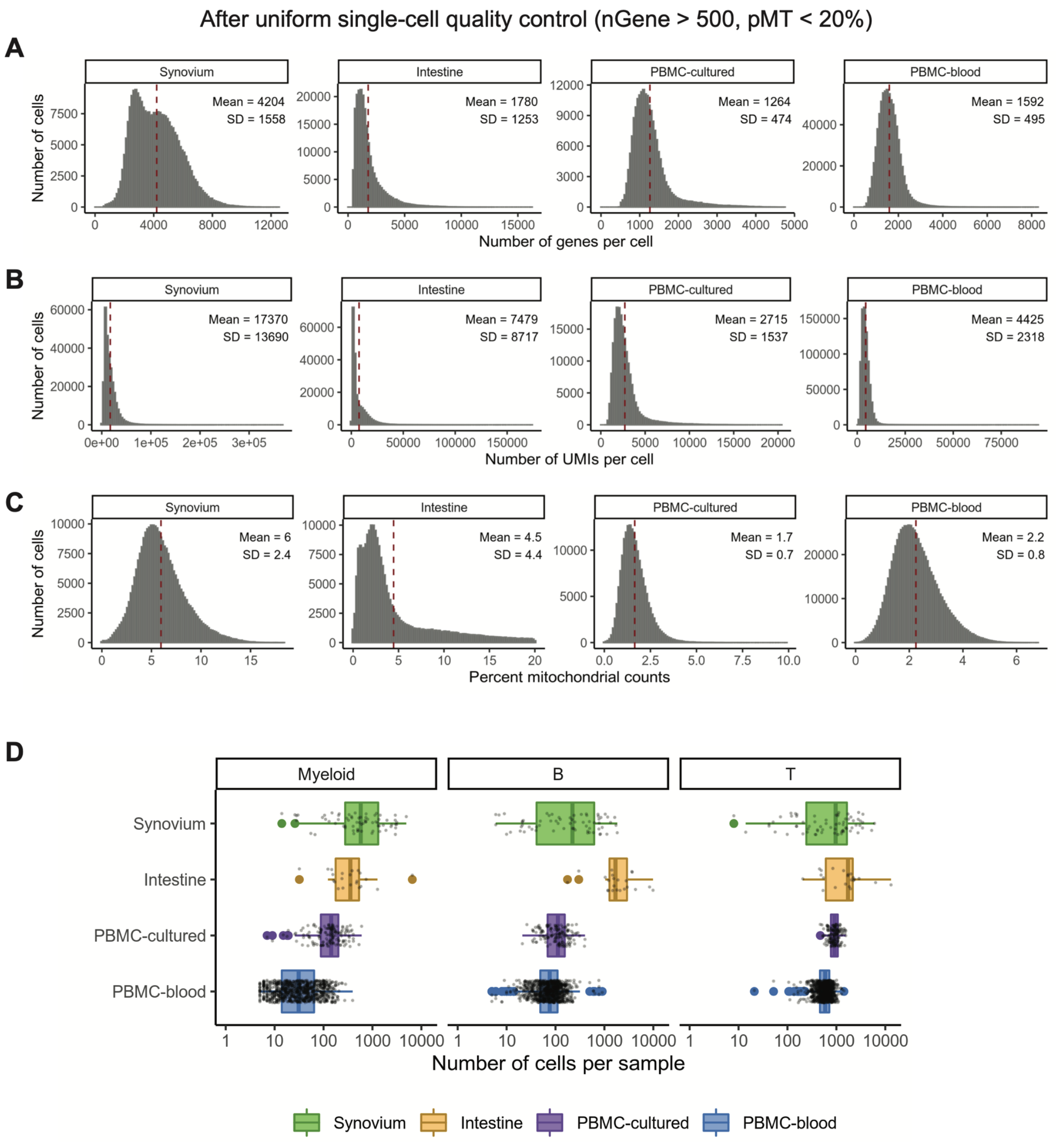

**Fig. S6. Single-cell dataset metrics after QC.** Metrics for scRNA-seq data for each cohort after uniform QC (removing cells with fewer than 500 genes or greater than 20% mitochondrial UMIs). (**A**) The number of genes per cell. (**B**) The number of UMIs per cell. (**C**) The percentage of mitochondrial UMIs per cell. The red dotted line indicates the mean value across cells; mean and standard deviation (SD) are listed. (**D**) Number of cells per sample in eQTL analysis by cell type and cohort (colors). The center line of the boxplot represents the median, the lower and upper box limits represent the 25% and 75% quantiles, respectively, the whiskers extend to the box limit ±1.5 × IQR, and all points are plotted individually.

A

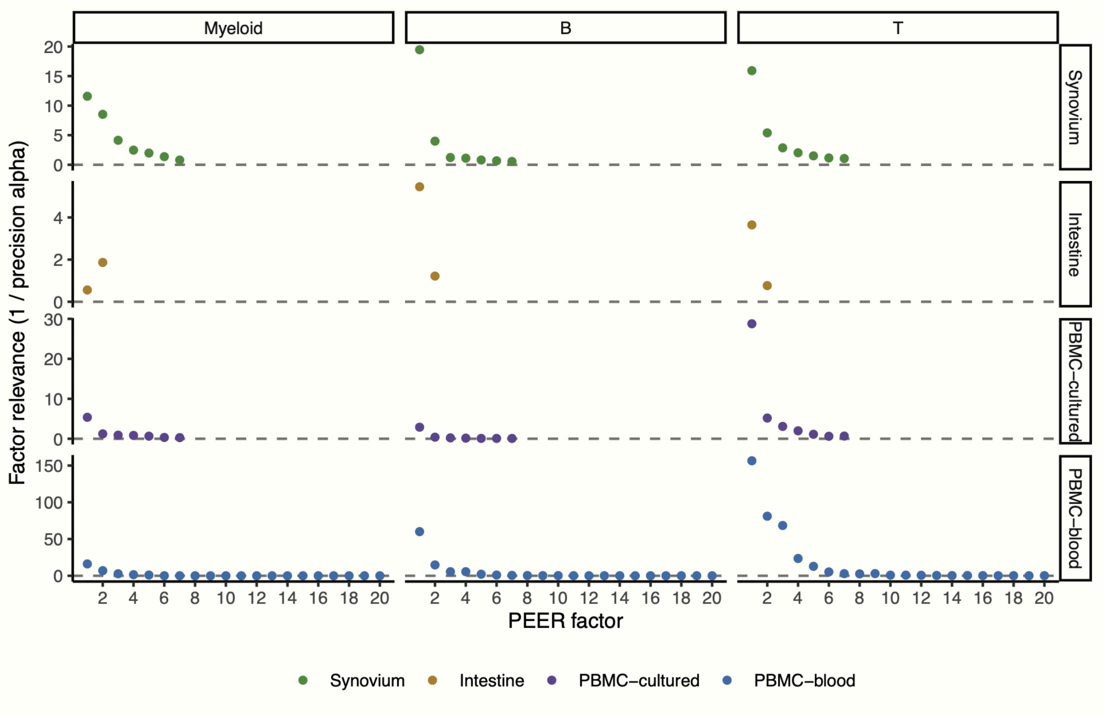

B

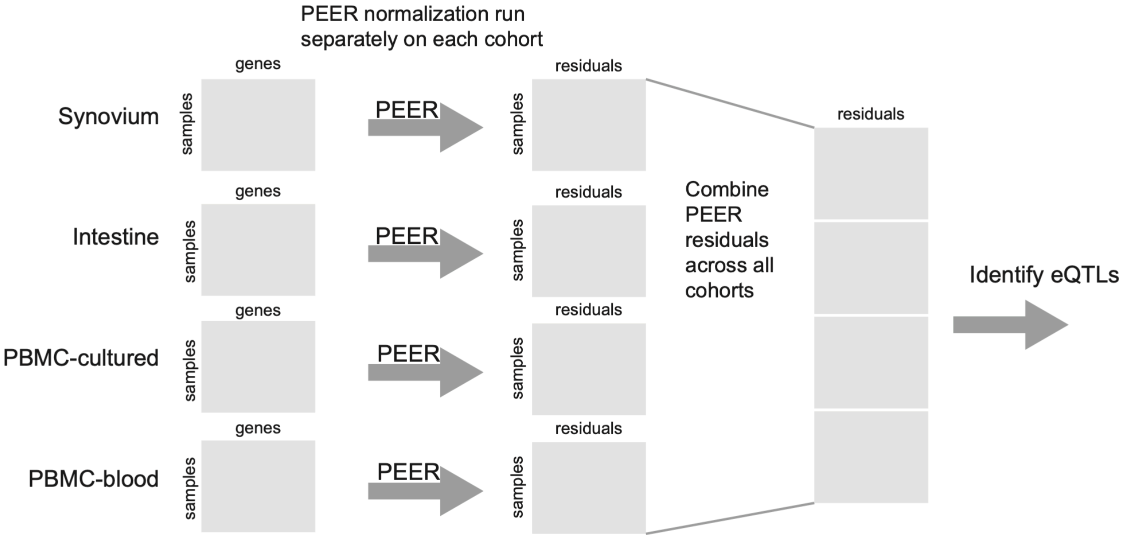

**Fig. S7. PEER factor relevance and schematic of the multi-cohort model.** (**A**) Relevance of each PEER factor (*y*-axis) for each dataset and cell type. Different numbers of PEER factors were used for each cohort (K=7 for Synovium, 2 for Intestine, 7 for PBMC-cultured, and 20 for PBMC-blood). (**B**) Schematic of pseudobulk eQTL multi-cohort analysis strategy. Cells were mean-aggregated within each sample to obtain a samples-by-genes matrix for each cohort and cell type. Then, we ran inverse normal transformation and PEER factor normalization separately within each cohort to obtain a samples-by-residuals matrix for each cohort. We concatenated these matrices into a single matrix across all cohorts. We identified eQTLs for *HLA* genes using a single linear model, modeling the residual as a function of genotype and cohort across all individuals.

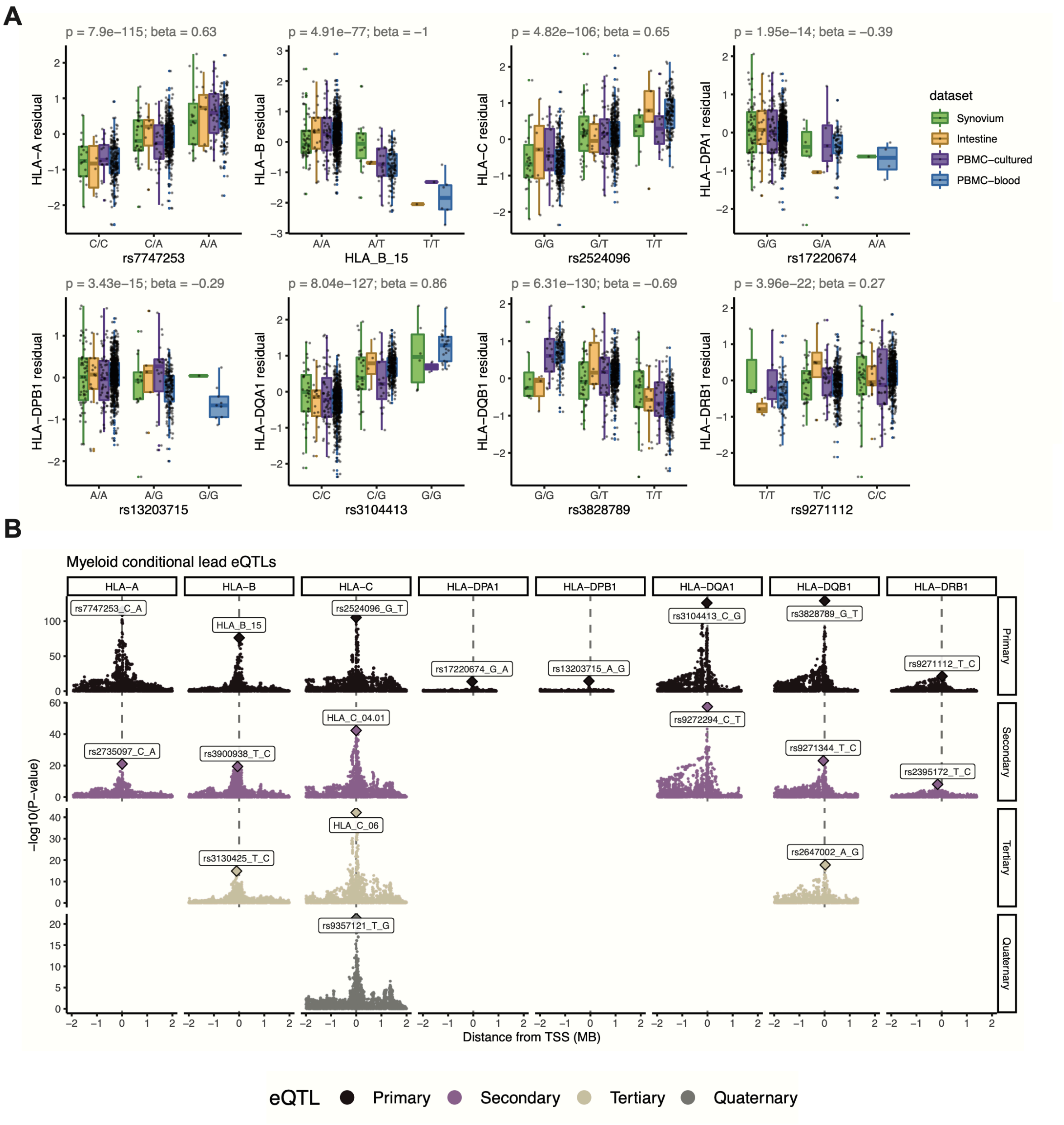

**Fig. S8. Pseudobulk eQTL results in myeloid cells.** (**A**) Boxplot showing the effect of the lead eQTL for each gene from the multi-cohort model. The genotype of each individual (*x*-axis) is plotted against the inverse-normal transformed residual of the gene's expression (after adjusting for covariates, *y*-axis), colored by cohort. Variants starting with "HLA" denote *HLA* alleles. Boxplot center line represents the median; lower and upper box limits represent the 25% and 75% quantiles, respectively; whiskers extend to box limit ±1.5 × IQR; outlying points are plotted individually. (**B**) We performed up to three additional rounds of conditional analysis to identify independent eQTLs. Manhattan plots showing the distance from TSS (*x*-axis, TSS ± 2MB of each gene) versus the significance of association with gene expression (*y*-axis). Each row represents one round of conditional analysis, and each subsequent round controls for the lead effects from the previous rounds. Blank elements in the grid indicate that no variants reach *P*-value < 5e-8.

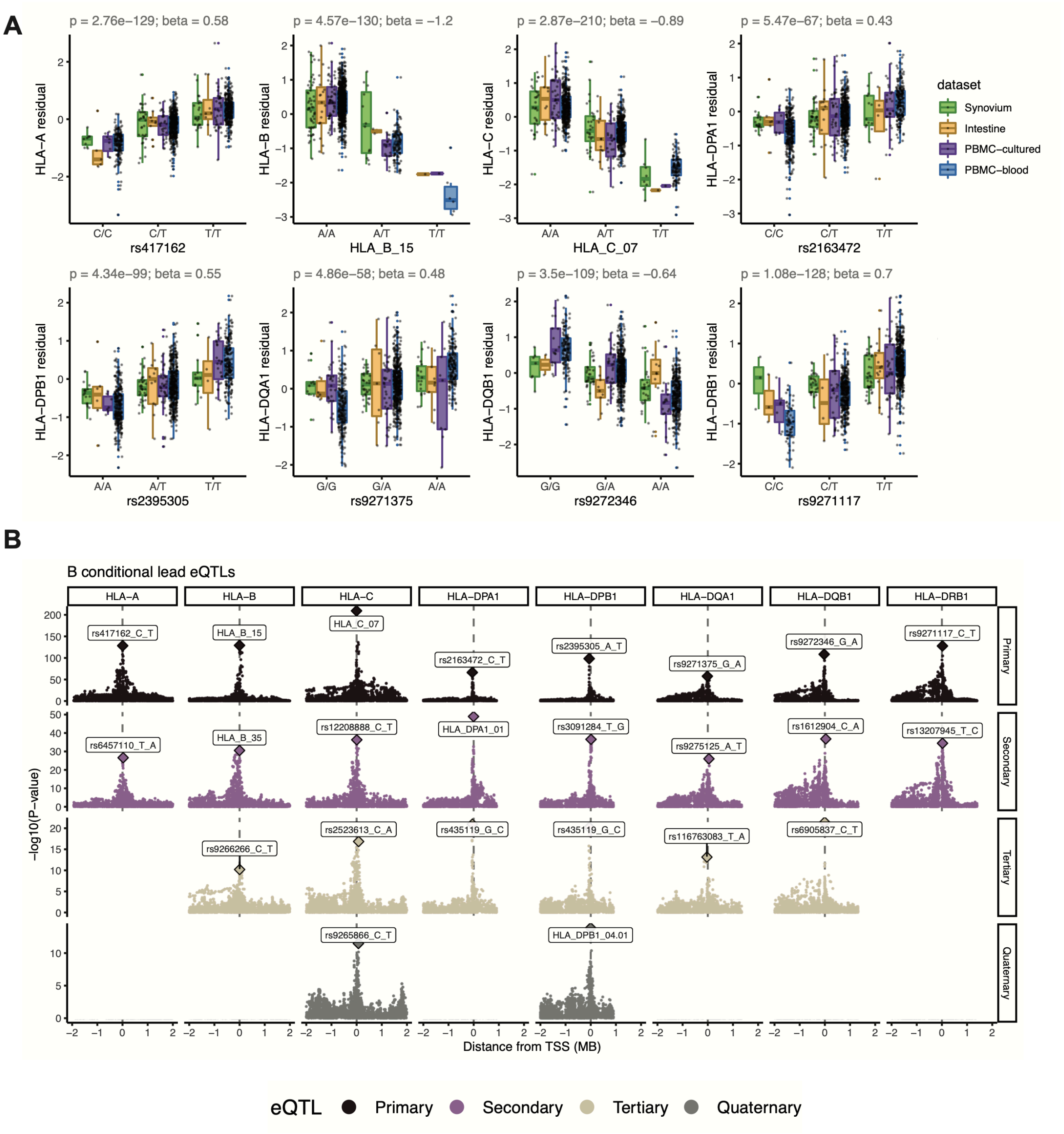

**Fig. S9. Pseudobulk eQTL results in B cells.** Same as **Fig. S8** but for B cells.

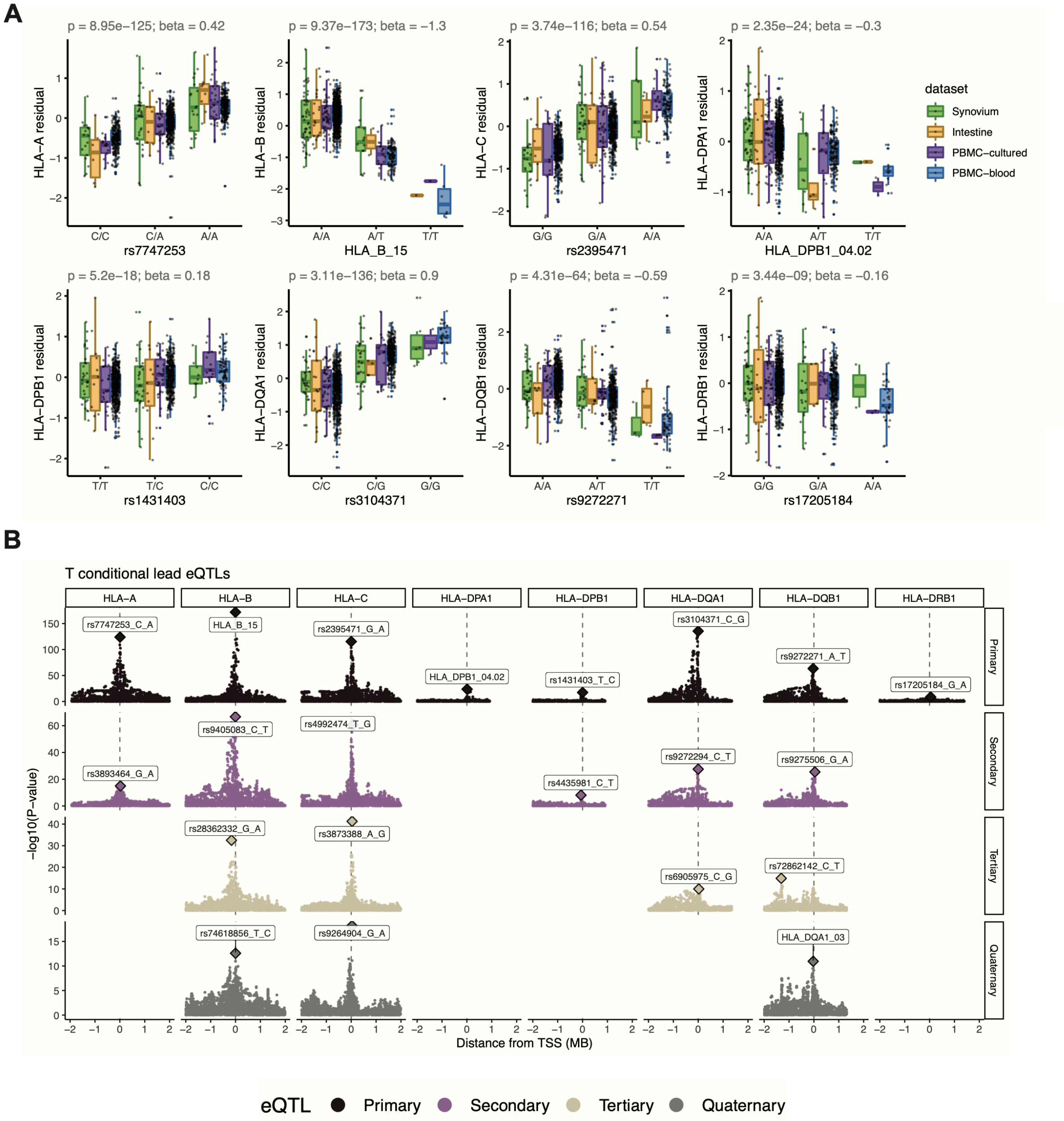

**Fig. S10. Pseudobulk eQTL results in T cells.** Same as **Fig. S8** but for T cells.

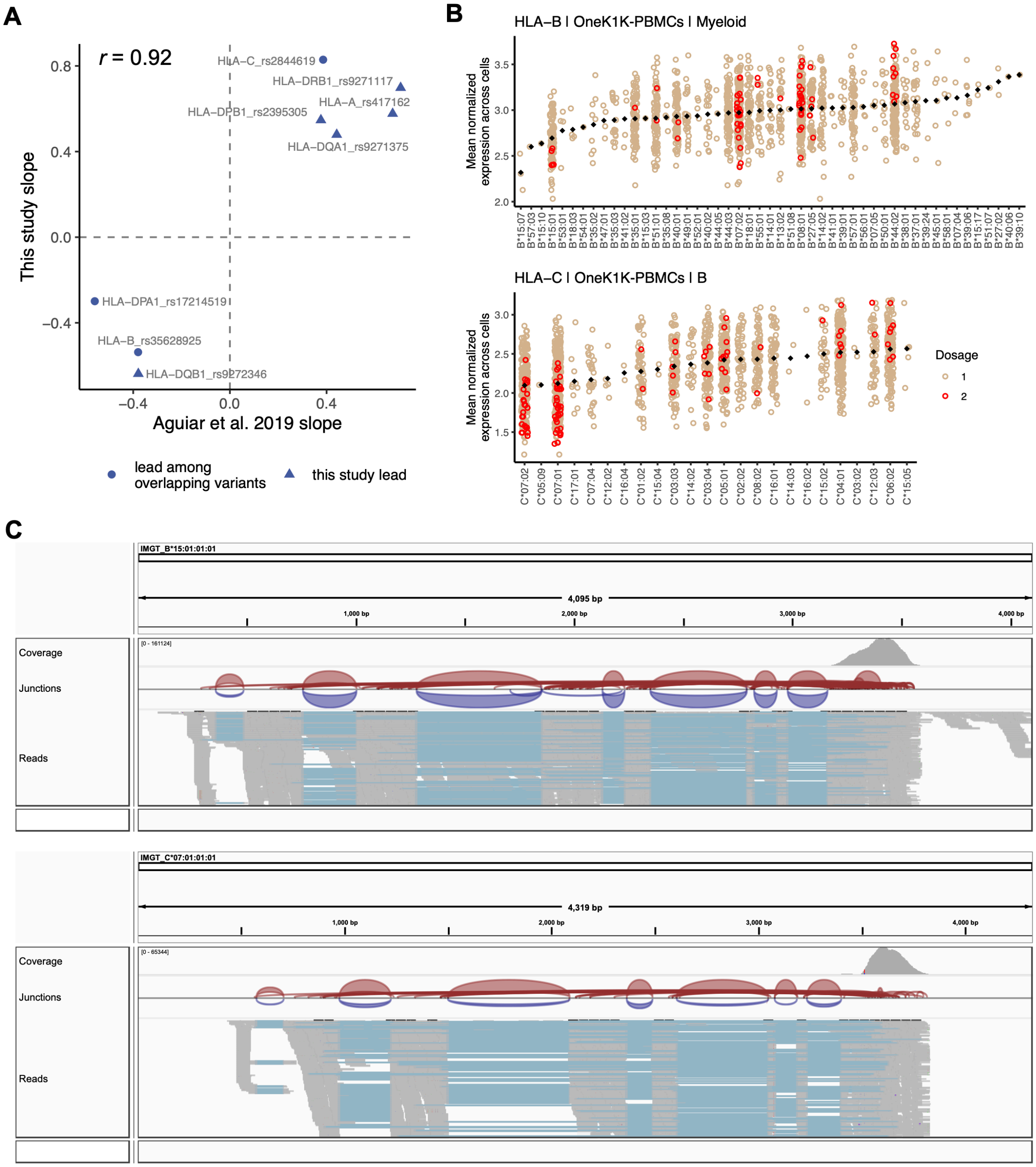

**Fig. S11. Concordance with Aguiar et al., differential allelic expression, and IGV read alignment visualization.** (**A**) Concordance between the effect sizes of lead *HLA* eQTLs identified in the multi-cohort pseudobulk model for B cells (this study, *y*-axis) and the same variant's effect in LCLs identified through bulk RNA-seq eQTL analysis (Aguiar et al., *x*-axis). Because not all lead variants in this study were directly comparable due to different sets of tested variants, we tested the concordance of the most significant variant present in both datasets (triangles indicate that the exact lead variant in this study was also tested in Aguiar et al., whereas circles indicate "substitute" lead variants was used for comparison). (**B**) *HLA-B* expression in myeloid cells (top) and *HLA-C* expression in B cells (bottom), showing mean log(CP10k+1)-normalized expression (*y*-axis) across cells for each individual in PBMC-blood by allele (*x*-axis). Each individual's expression value is plotted once if they are homozygous (red) and twice if heterozygous (tan) for each allele (imputed dosage is rounded to the nearest integer). The black diamonds show the mean value for each allele (used to order the *x*-axis). (**C**) IGV screenshots showing read alignments for alleles *HLA-B*15:01* and *HLA-C*07:01*, associated with lower expression of the respective genes, for a representative individual in Synovium.

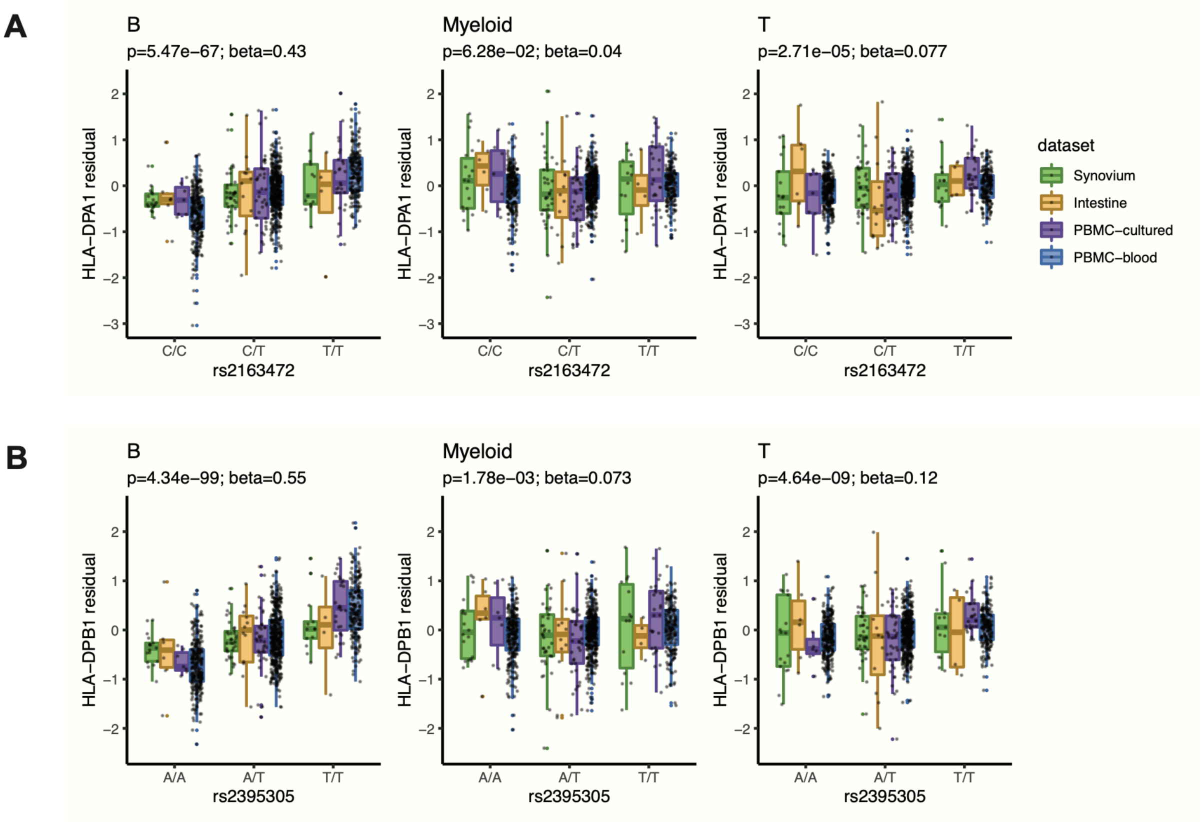

**Fig. S12. Examples of cell-type-dependent eQTLs for *HLA-DP* genes.** Boxplots across cell types (columns) comparing the effects of B cell lead eQTLs (rows) for (**A**) *HLA-DPA1* and (**B**) *HLA-DPB1*. In both examples, the lead eQTL was identified in B cells and was weaker in myeloid and T cells. The genotype of each individual (*x*-axis) is plotted against the inverse-normal transformed residual of the gene's expression (after adjusting for covariates, *y*-axis). Boxplots are colored by cohort, with individual points overlaid; center line represents the median; lower and upper box limits represent the 25% and 75% quantiles, respectively; whiskers extend to box limit ±1.5 × IQR; outlying points are plotted individually.

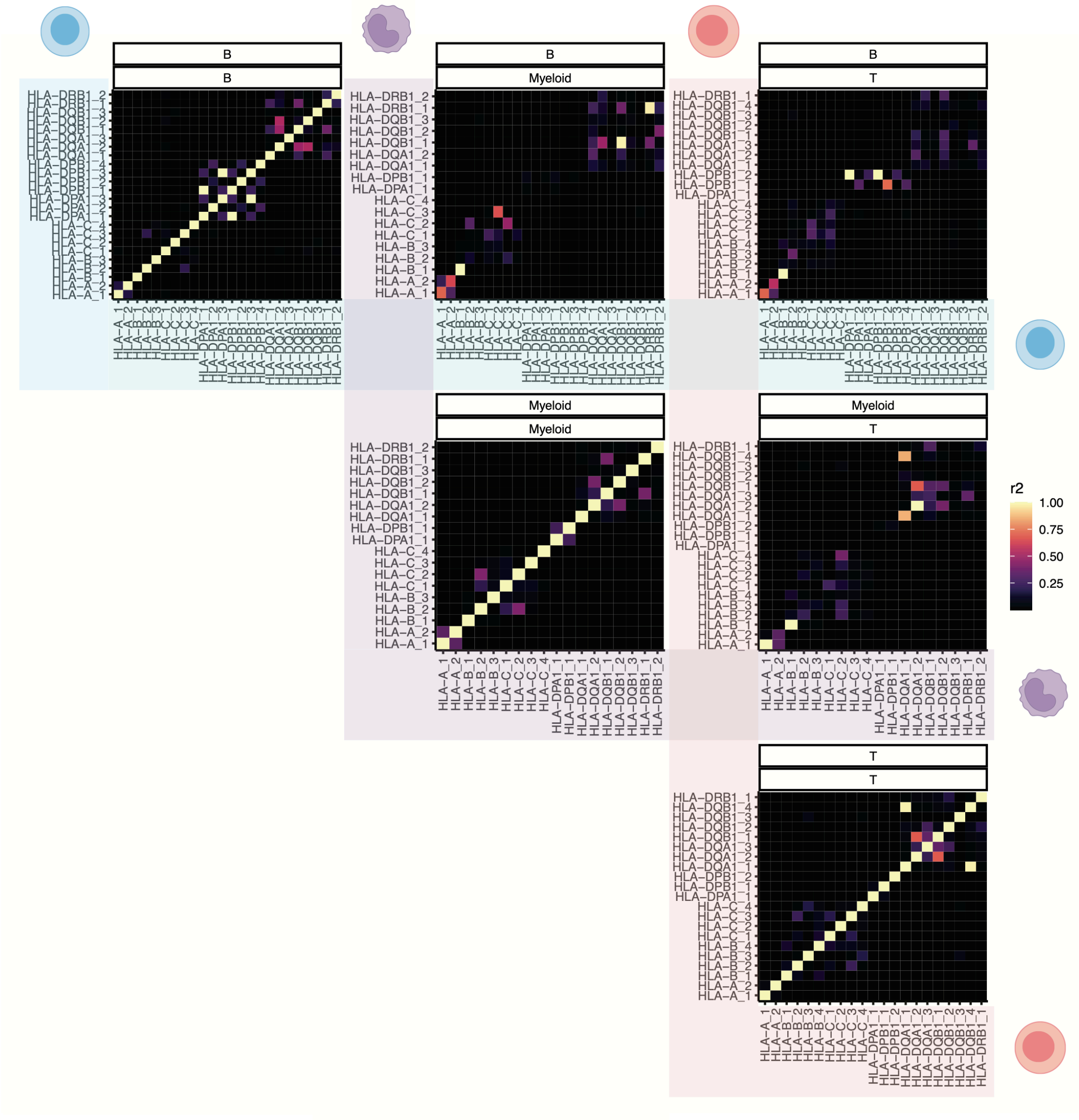

**Fig. S13. Linkage disequilibrium among conditionally independent eQTLs across genes and cell types.** Heatmaps showing the LD relationship among lead eQTL variants identified in the multiple rounds of conditional analysis. For each pair of cell types among B (blue), myeloid (purple), and T cells (red) (including self-pairs), the plot shows the LD ($r^2$, color) between each pair of eQTLs. Each eQTL is labeled as HLA-X_Y, where X is the gene and Y is the round of conditional analysis (e.g., HLA-B_3 represents the tertiary eQTL for *HLA-B*). LD is calculated using the multi-ancestry MHC reference used for HLA imputation.

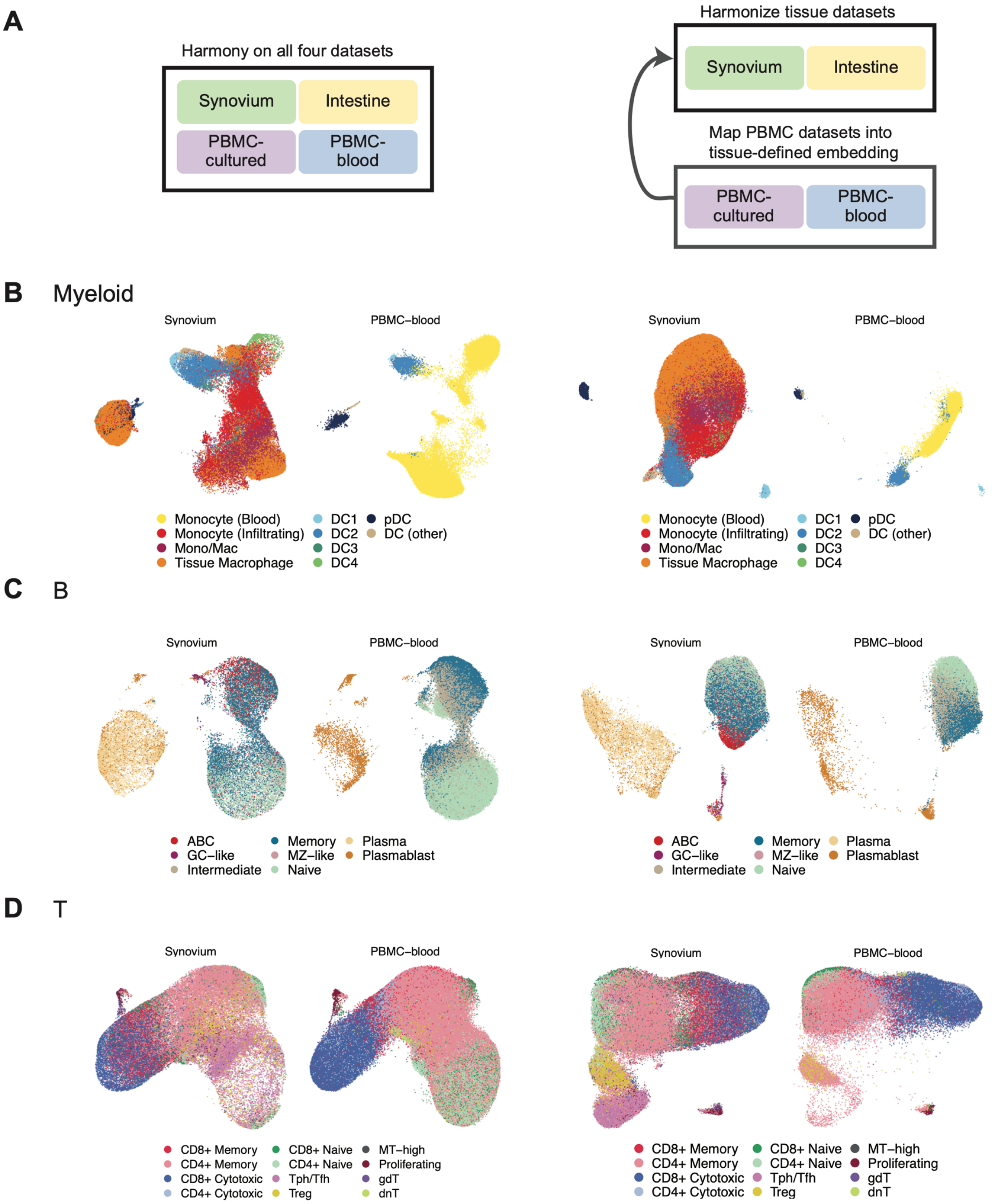

**Fig. S14. Two strategies for embedding cells from multiple datasets.** (**A**) Schematic of *de novo* integration of all datasets using Harmony (left) versus a reference-mapping-based approach where the two solid tissue datasets were used to construct the embedding, and PBMC datasets were mapped into the same coordinate space using Symphony (right). (**B-D**) The resulting UMAP embeddings for Synovium and PBMC-blood datasets using each approach for (**B**) myeloid, (**C**) B, and (**D**) T cells, colored by merged cell state annotations.

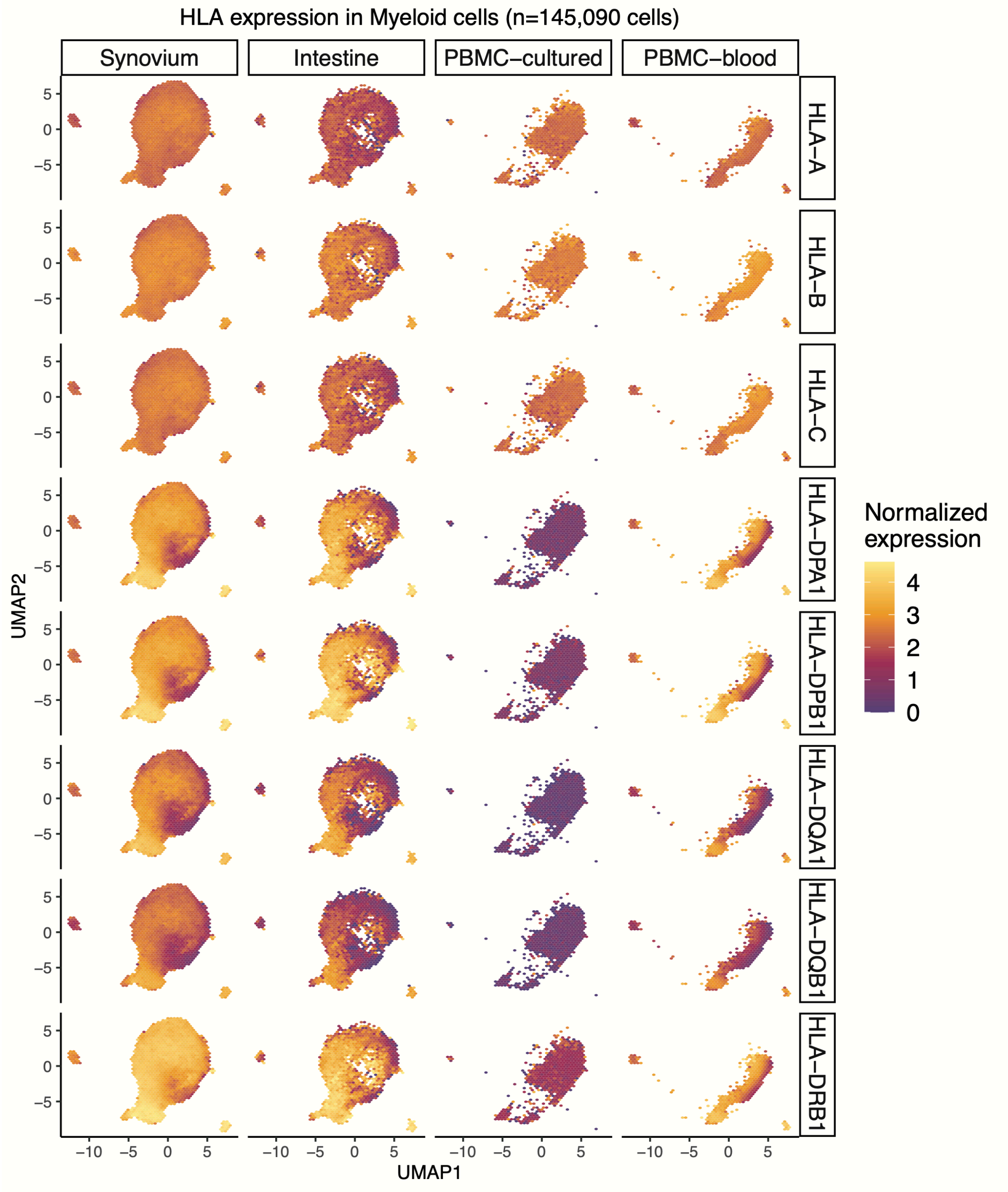

**Fig. S15. Atlas of *HLA* gene expression in myeloid cells across four datasets.** Expression of eight classical *HLA* genes (rows) in myeloid cells in Synovium (n=66,789 cells), Intestine (n=14,492 cells), PBMC-cultured (n=23,241 cells), and PBMC-blood (n=40,568 cells) plotted on a hexagon-binned UMAP to address overplotting (50 bins per both horizontal and vertical directions), with each bin colored by mean log(CP10k+1)-normalized expression of the gene.

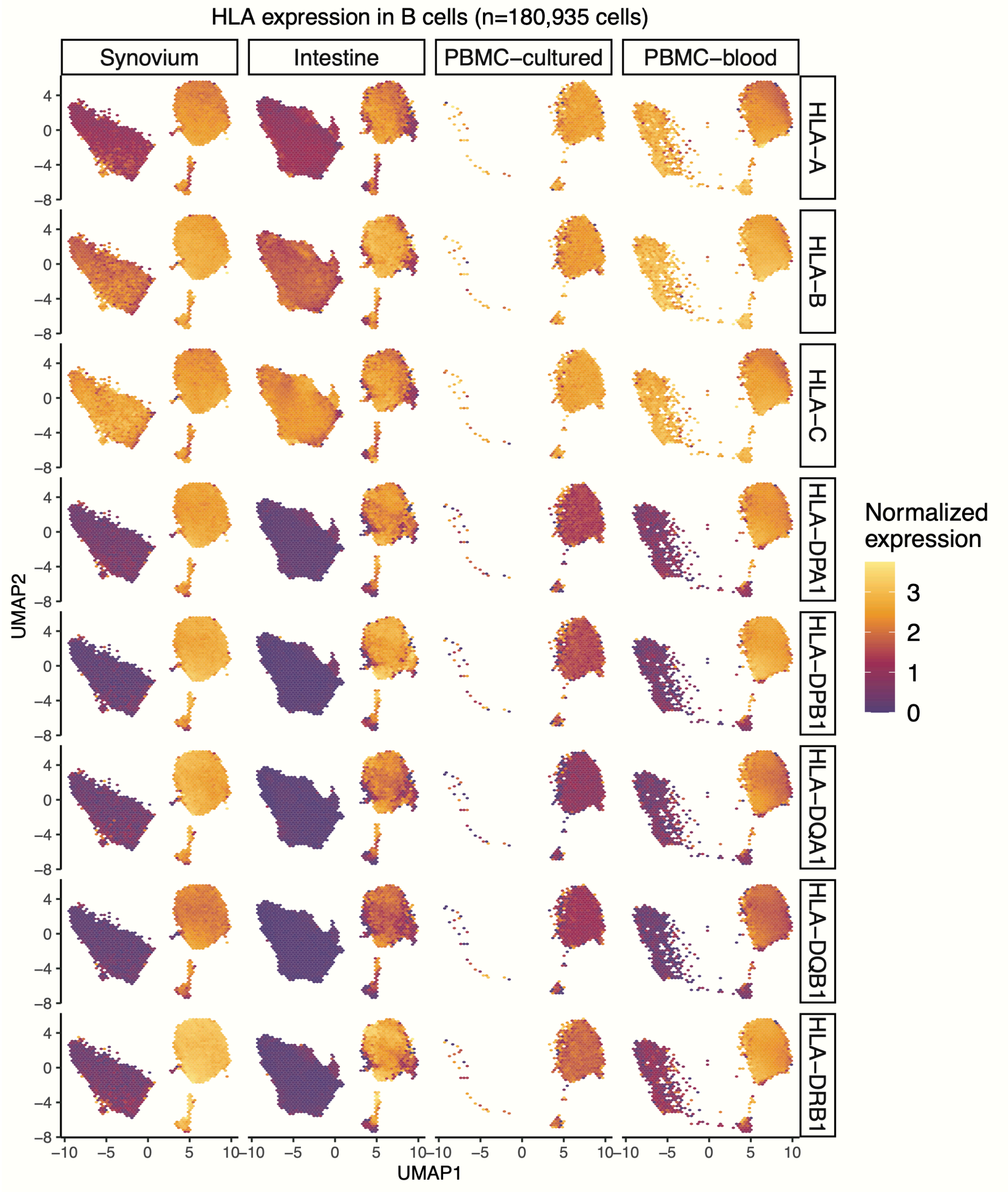

**Fig. S16. Atlas of *HLA* gene expression in B cells across four datasets.** Same as **Fig. S15** but for B cells in Synovium (n=25,917 cells), Intestine (n=56,572 cells), PBMC-cultured (n=17,662 cells), and PBMC-blood (n=80,784 cells).

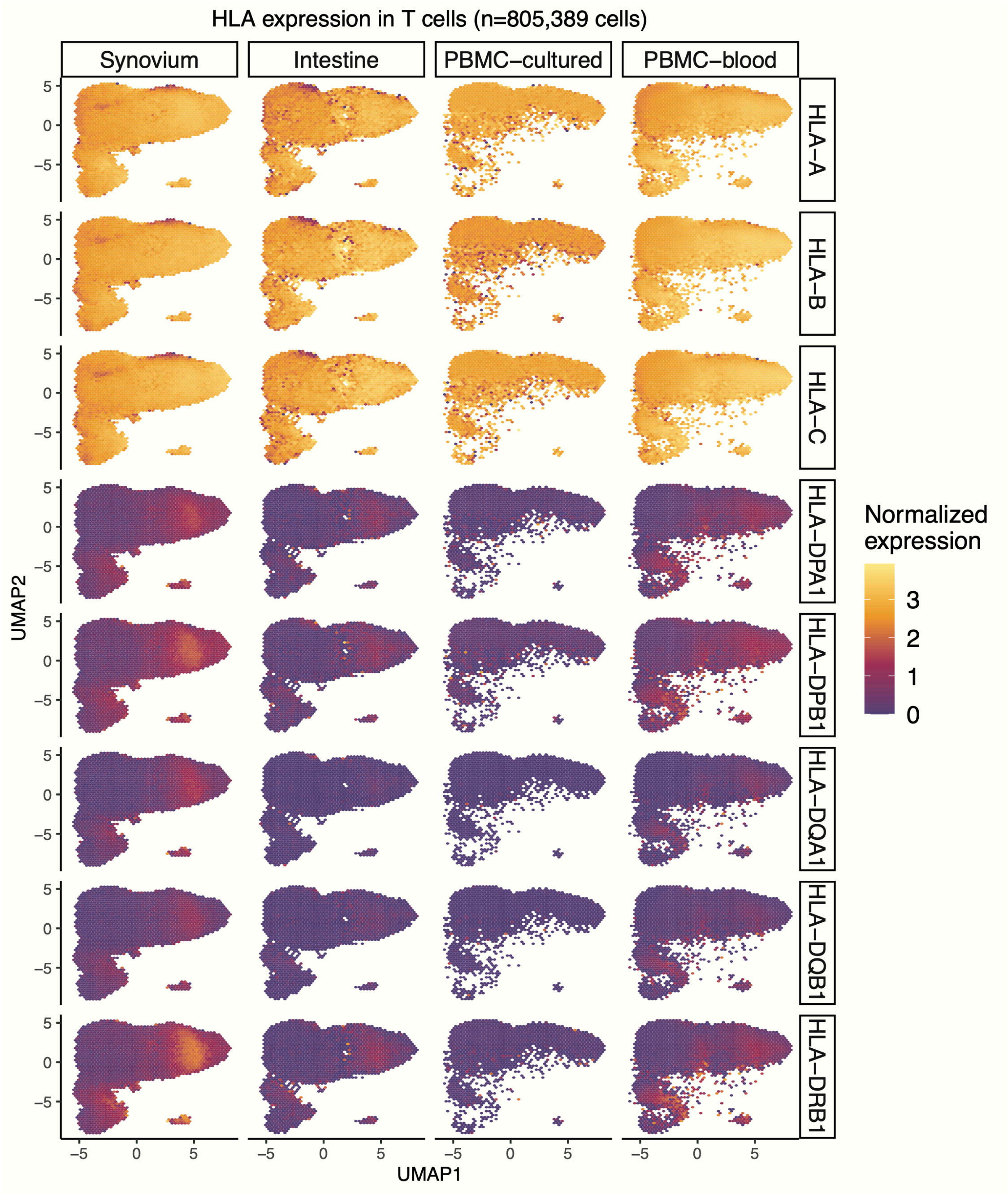

**Fig. S17. Atlas of *HLA* gene expression in T cells across four datasets.** Same as **Fig. S15** but for T cells in Synovium (n=82,423 cells), Intestine (n=47,868 cells), PBMC-cultured (n=136,519 cells), and PBMC-blood (n=538,579 cells).

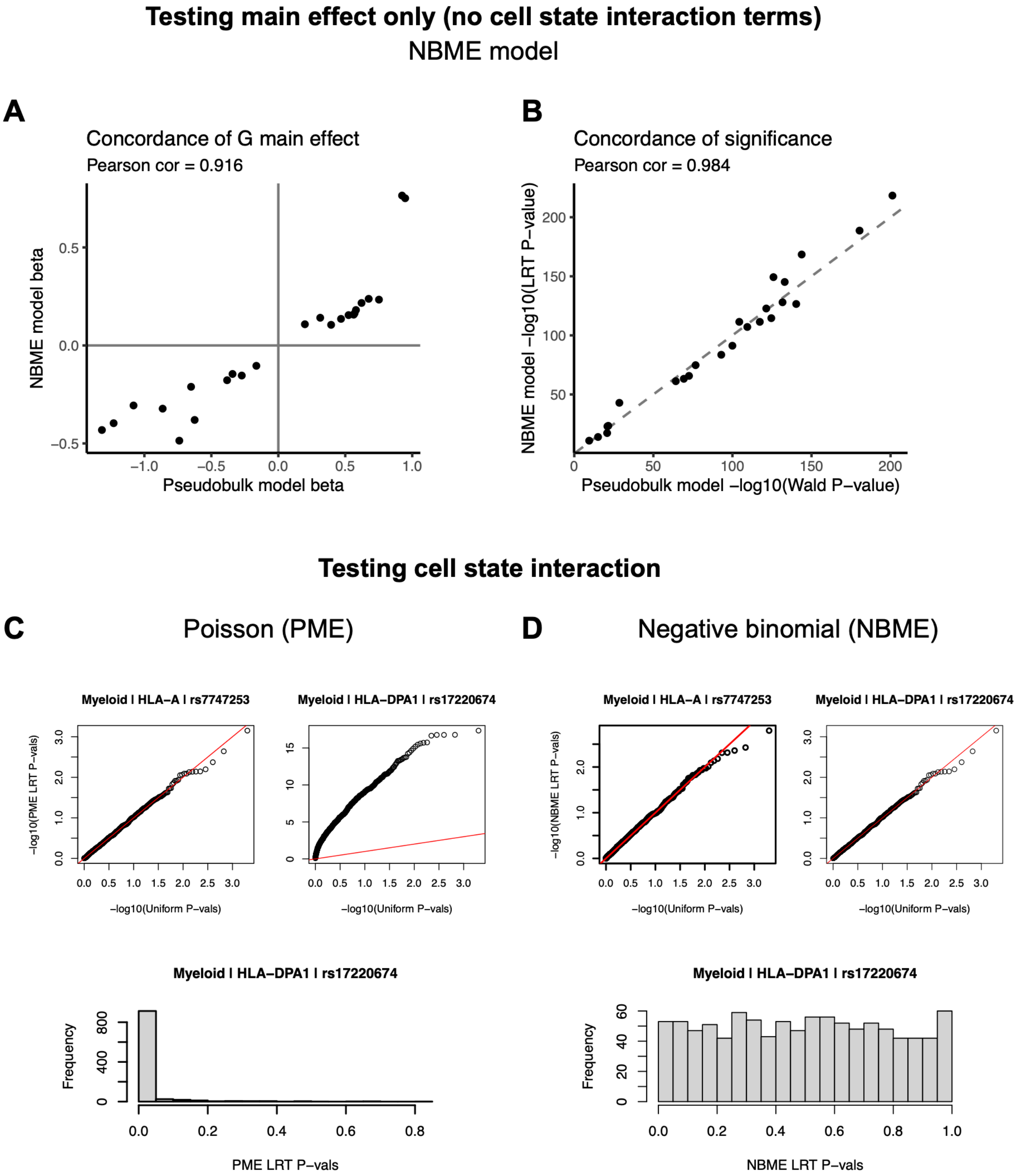

**Fig. S18. Testing single-cell NBME model for concordance with pseudobulk models and for calibration for genotype-cell-state interactions.** The models in (**A-B**) test genotype main effects, whereas (**C-D**) test genotype-cell-state interaction. (**A-B**) Concordance of genotype main effect estimates (**A**) and significance of genotype main effect (**B**) between the NBME model (*y*-axis) and the pseudobulk model for the PBMC-blood dataset (*x*-axis) across all cell types and classical *HLA* genes. (**C-D**) We permuted cell state (10 hPCs as a block) for 1,000 tests and obtained interaction *P*-values from a likelihood ratio test (LRT) comparing to the null model without GxhPC interaction terms. Q-Q plots showing statistical calibration (compared to uniform *P*-values) for (**C**) PME model versus (**D**) NBME model when testing for cell state interactions for representative class I (*HLA-A*) and class II (*HLA-DPA1*) genes in myeloid cells in PBMC-blood. The red line is the identity line. The histograms below show distributions of LRT *P*-values for *HLA-DPA1*.

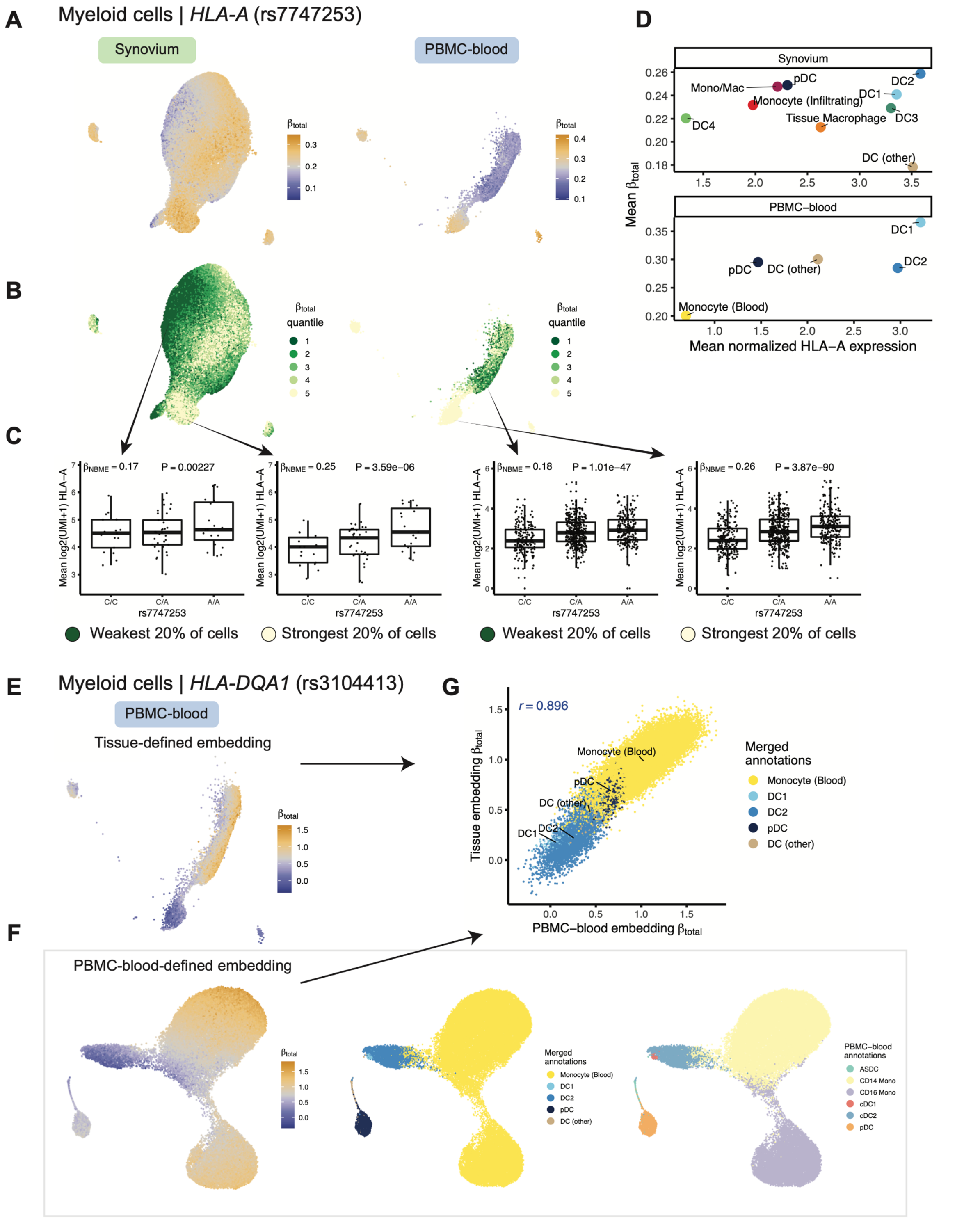

**Fig. S19. Dynamic eQTLs in myeloid cells.** Lead *HLA-A* eQTL (rs7747253) in myeloid cells (n=66,789 cells in Synovium, 40,568 in PBMC-blood). (**A**) UMAP colored by per-cell estimated eQTL effect ($\beta_{total}$), from blue (weakest) to orange (strongest). (**B**) Cells colored by quintiles of $\beta_{total}$. (**C**) Boxplot showing the eQTL effect across individuals in the top and bottom quintiles of cells. Labeled $\beta_{NBME}$ and *P*-value are derived from fitting the NBME model without cell state interaction terms on cells from the discrete quintile and comparing to a null model without genotype using an LRT. Mean log2(UMI+1) across cells per individual (*y*-axis) by each genotype. Boxplot center line represents the median; lower and upper box limits represent the 25% and 75% quantiles, respectively; whiskers extend to box limit ±1.5 × IQR; outlying points are plotted individually. (**D**) Scatter plot showing the mean estimated $\beta_{total}$ (*y*-axis) compared to the mean log(CP10k+1)-normalized expression of *HLA-A* (*x*-axis) across annotated cell states (color). (**E-G**) Comparing myeloid *HLA-DQA1* eQTL (rs3104413) effects in two different cell embeddings. UMAP of PBMC-blood myeloid cells (n=40,568 cells) in (**E**) tissue-defined hPCs versus (**F**) hPCs defined using PBMC-blood alone, colored by $\beta_{total}$ (left), merged cell annotations (middle), and dataset annotations (right). (**G**) Concordance between per-cell $\beta_{total}$ values in tissue-defined (*y*-axis) versus PBMC-blood embedding (*x*-axis); Pearson *r* is labeled.

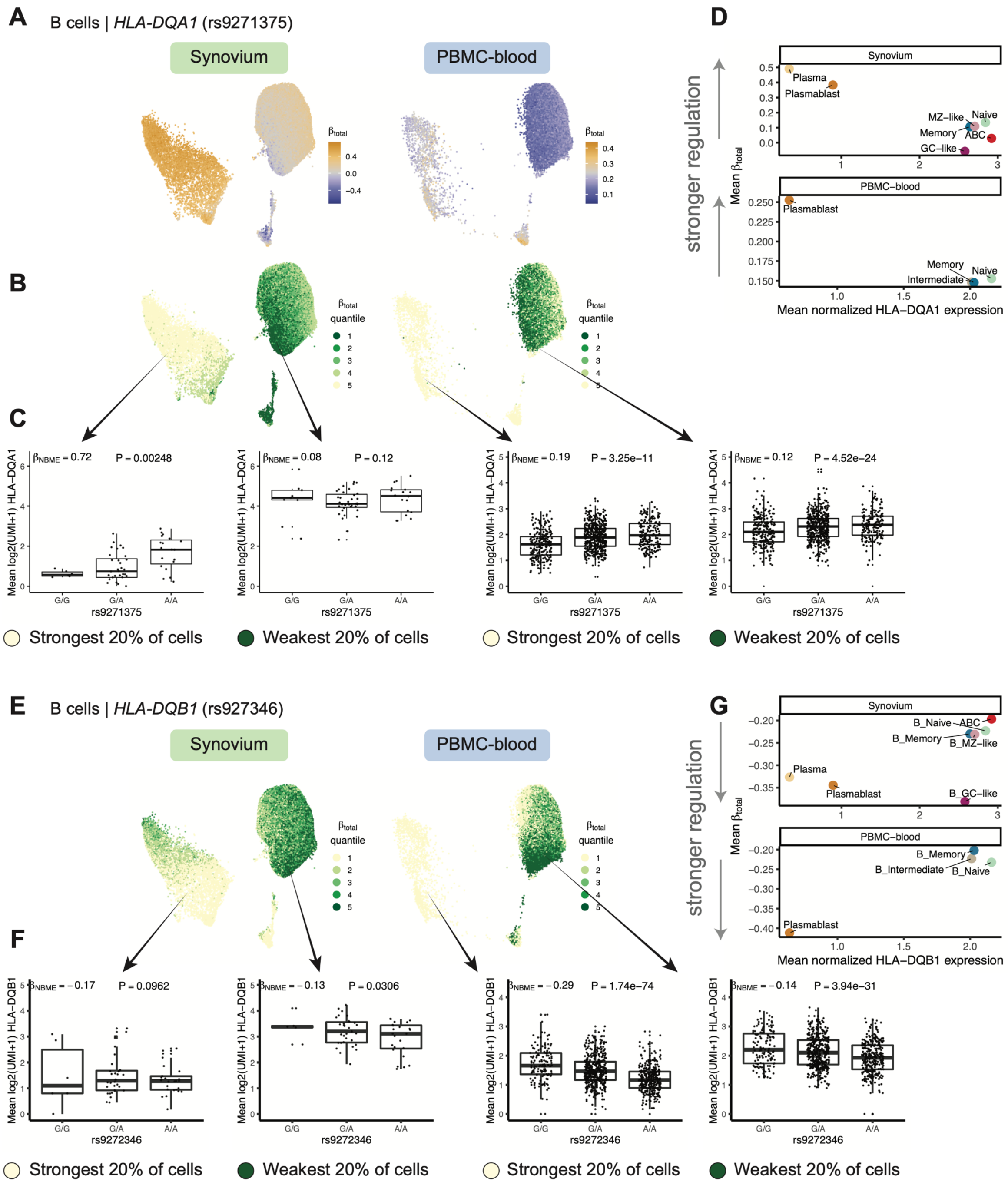

**Fig. S20. Dynamic *HLA-DQ* eQTLs in B cells.** (**A-D**) Dynamic *HLA-DQA1* eQTL in B cells (n=25,917 cells in Synovium, n=80,784 in PBMC-blood). (**A**) UMAP of cells for tissue-defined embedding, colored by $\beta_{total}$, from blue (weakest) to orange (strongest). (**B**) Cells colored by quintiles of $\beta_{total}$. (**C**) Boxplot showing the eQTL effect across individuals in the bottom and top quintiles. Labeled $\beta_{NBME}$ and *P*-value are from fitting the NBME model without cell state interaction terms on the cells from the discrete quintile and comparing to a null model without genotype using an LRT. Mean log2(UMI+1) across cells per individual (*y*-axis) by each genotype. Boxplot center line represents the median; lower and upper box limits represent the 25% and 75% quantiles, respectively; whiskers extend to box limit ±1.5 × IQR; outlying points are plotted individually. (**D**) Scatter plot showing the mean estimated $\beta_{total}$ (*y*-axis) compared to the mean log(CP10k+1)-normalized expression of *HLA-DQA1* (*x*-axis) across annotated cell states (color). Panels (**E**-**G**) are analogous to panels (**B-D**), respectively, for lead *HLA-DQB1* eQTL in B cells. Note: in this *HLA-DQB1* example, the effect of the eQTL is negative as defined by the ALT allele, so more negative $\beta_{NBME}$ (quintile 1) corresponds to stronger eQTL effect.

| Dataset abbrev. | Synovium | Intestine | PBMC-cultured | PBMC-blood |
|---|---|---|---|---|
| | **Cohort characteristics** | | | |
| **Source of data** | AMP Phase 2 Consortium Zhang et al. (in revision; *bioRxiv*, 2022) | Smillie et al. (*Cell,* 2019) | Randolph et al. (*Science,* 2021) | OneK1K Cohort Yazar et al. (*Science*, 2022) |
| **Tissue** | Synovial joint | Intestine | PBMCs | PBMCs |
| **Conditions** | RA (60) and OA (9) | UC (13) and HC (9) | IAV (73) and control (73) | N/A (population cohort) |
| **# indivs.** | 69 | 22 | 73 | 909 |
| **# cells** | 275,323 | 137,321 | 188,507 | 765,079 |
| **Ancestry** | Multiple | Multiple | European and African | European |
| | **Dataset technical details** | | | |
| **Genotype assay (# MHC variants)** | Illumina MEGA (10,159) | Illumina GSA (7,544) or custom array (772) | Low-pass WGS (10,814) | Illumina GSA (7,046) |
| **Single-cell assay** | 10x CITE-seq (3’ v3) | 10x scRNA-seq (3’ v1, v2) | 10x scRNA-seq (3’ v2) | 10x scRNA-seq (3’ v2) |
| **Read length(s)** | 94 bp | 55 bp | 84 and 289 bp | 98 bp |
| **Input format** | FASTQ | FASTQ | BAM | BAM |
| **Barcode length** | 16 | 14 and 16 | 16 | 16 |
| **UMI length** | 12 and 10 | 10 | 10 | 10 |
| **Whitelist (10x Genomics)** | V3 (3M-febrary-2018.txt.gz ) | V1 (737K-april-2014_rc.txt) and V2 (737K-august-2016.txt) | V2 (737K-august-2016.txt) | V2 (737K-august-2016.txt) |
| **Notes** | Some samples have 10bp UMI but all use v3 whitelist | Barcode length = 14 for V1 | Demultiplexed batch-level BAMs with sinto | Demultiplexed batch-level BAMs with sinto |

**Table S1. Datasets included in the study.** Cohort characteristics include reference publication, sampled tissue, biological conditions (if any), number of individuals, number of single cells, and genetic ancestry. The numbers of individuals and cells are shown after removing individuals with uncertain *HLA* allele calls and low-quality cells. All PBMC-cultured individuals had samples from both conditions (treated with influenza A virus and mock conditions). Dataset technical details include type of genotype data (and # of variants in the MHC as input to HLA imputation), single-cell assay, read length(s), input format, barcode and UMI length, and whitelist used for STARsolo. Abbreviations: PBMCs, peripheral blood mononuclear cells; RA, rheumatoid arthritis; OA, osteoarthritis; UC, ulcerative colitis; HC, healthy control; IAV, influenza A virus; MHC, major histocompatibility complex; WGS, whole-genome sequencing; GSA, Global Screening Array; MEGA, Multi-ethnic Genotyping Array.

| | Dataset | Synovium | Intestine | PBMC-cultured | PBMC-blood | Total |
|---|---|---|---|---|---|---|
| **Individual-level counts** | **# individuals with genotype and scRNA** | 78 | 25 | 89 | 973 | 1165 |
| | **# individuals passing genotype QC** | 78 | 25 | 88 | 969 | 1160 |
| | **# individuals with complete HLA imputation** | 69 | 22 | 73 | 909 | 1073 |
| **Cell-level counts** | **# cells before cell QC** | 278238 | 265629 | 189488 | 772252 | 1505607 |
| | **# Myeloid** | 66789 | 14492 | 23241 | 40568 | 145090 |
| | **# B** | 25917 | 56572 | 17662 | 80784 | 180935 |
| | **# T** | 82423 | 47868 | 136519 | 538579 | 805389 |
| | **# NK** | 7749 | 1883 | 11085 | 105148 | 125865 |
| | **# Fibroblast** | 69246 | 13405 | 0 | 0 | 82651 |
| | **# Endothelial** | 23199 | 3101 | 0 | 0 | 26300 |
| | **Sum (all cell types)** | 275323 | 137321 | 188507 | 765079 | 1366230 |
| | **Sum (myeloid, B, T)** | 175129 | 118932 | 177422 | 659931 | 1131414 |

**Table S2. Sample and cell numbers before and after QC.** Top section includes number of individuals per dataset before QC and during each step. Bottom section includes number of cells before QC and cell counts for each major cell type and total count after QC per dataset. Bottom row shows sum of cells used in eQTL analysis (myeloid, B, and T cells).

| **Mean DR2 for two-field alleles with AF > 5%** | | | | | | |
|---|---|---|---|---|---|---|
| **Gene** | **Synovium** | **Intestine (GSA)** | **Intestine (custom)** | **PBMC-cultured** | **PBMC-blood** | **Mean** |
| ***HLA-A*** | 0.960 | 0.966 | 0.966 | 0.914 | 0.966 | 0.954 |
| ***HLA-B*** | 0.943 | 0.950 | 0.958 | 0.940 | 0.961 | 0.950 |
| ***HLA-C*** | 0.965 | 0.977 | 0.973 | 0.951 | 0.976 | 0.968 |
| ***HLA-DPA1*** | 0.960 | 0.960 | 0.960 | 0.963 | 0.970 | 0.963 |
| ***HLA-DPB1*** | 0.968 | 0.972 | 0.948 | 0.978 | 0.972 | 0.968 |
| ***HLA-DQA1*** | 0.971 | 0.977 | 0.977 | 0.977 | 0.977 | 0.976 |
| ***HLA-DQB1*** | 0.982 | 0.972 | 0.967 | 0.950 | 0.971 | 0.968 |
| ***HLA-DRB1*** | 0.947 | 0.920 | 0.919 | 0.877 | 0.958 | 0.924 |
| **Mean DR2 for two-field alleles with AF > 1%** | | | | | | |
| **Gene** | **Synovium** | **Intestine (GSA)** | **Intestine (custom)** | **PBMC-cultured** | **PBMC-blood** | **Mean** |
| ***HLA-A*** | 0.913 | 0.912 | 0.881 | 0.861 | 0.919 | 0.897 |
| ***HLA-B*** | 0.880 | 0.890 | 0.908 | 0.897 | 0.908 | 0.897 |
| ***HLA-C*** | 0.922 | 0.944 | 0.929 | 0.901 | 0.948 | 0.929 |
| ***HLA-DPA1*** | 0.864 | 0.943 | 0.937 | 0.886 | 0.950 | 0.916 |
| ***HLA-DPB1*** | 0.927 | 0.905 | 0.884 | 0.909 | 0.922 | 0.909 |
| ***HLA-DQA1*** | 0.971 | 0.967 | 0.969 | 0.954 | 0.969 | 0.966 |
| ***HLA-DQB1*** | 0.942 | 0.945 | 0.938 | 0.921 | 0.950 | 0.939 |
| ***HLA-DRB1*** | 0.881 | 0.880 | 0.882 | 0.834 | 0.868 | 0.869 |

**Table S3. SNP2HLA imputation quality for *HLA* alleles.** Mean imputation dosage $R^2$ (DR2) for two-field *HLA* alleles with AF > 5% (top) and >1% (bottom) for each *HLA* gene (row) across each array dataset (columns), as well as mean across datasets (rightmost column).

| | | Percent change in expression after personalization | | | |
|---|---|---|---|---|---|
| **Gene** | **Dataset** | **mean** | **median** | **q25** | **q75** |
| *HLA-A* | Synovium | 0.268 | 0.117 | -0.122 | 0.447 |
| | Intestine | -1.644 | 0.015 | -2.027 | 0.614 |
| | PBMC-cultured | 0.323 | 0.278 | 0.145 | 0.455 |
| | PBMC-blood | -0.015 | -0.012 | -0.094 | 0.073 |
| *HLA-B* | Synovium | -11.802 | -12.91 | -21.893 | 0.384 |
| | Intestine | -9.897 | -9.36 | -19.358 | 0.438 |
| | PBMC-cultured | -8.247 | -7.217 | -13.426 | 1.424 |
| | PBMC-blood | -12.071 | -13.907 | -21.57 | 0.758 |
| *HLA-C* | Synovium | 26.278 | 29.411 | 4.625 | 43.818 |
| | Intestine | 28.794 | 24.934 | 6.184 | 49.257 |
| | PBMC-cultured | 38.129 | 40.353 | 4.264 | 53.649 |
| | PBMC-blood | 5.339 | 2.224 | -0.062 | 12.058 |
| *HLA-DPA1* | Synovium | 3.022 | 0.098 | -0.088 | 4.348 |
| | Intestine | 0.857 | 0.044 | -0.018 | 1.552 |
| | PBMC-cultured | 5.995 | 4.103 | 0 | 10.563 |
| | PBMC-blood | 0.554 | 0.085 | 0 | 0.998 |
| *HLA-DPB1* | Synovium | 3.618 | 1.761 | -0.475 | 6.335 |
| | Intestine | 2.416 | 0.627 | -0.384 | 4.257 |
| | PBMC-cultured | 34.893 | 26.977 | 0.192 | 48.413 |
| | PBMC-blood | 1.055 | 0.237 | -0.273 | 1.962 |
| *HLA-DQA1* | Synovium | 29.007 | 20.261 | 2.834 | 44.221 |
| | Intestine | 22.159 | 15.612 | 2.788 | 32.822 |
| | PBMC-cultured | 43.461 | 10.767 | 2.875 | 60.908 |
| | PBMC-blood | 24.203 | 13.885 | 4.347 | 41.031 |
| *HLA-DQB1* | Synovium | 6.757 | 5.061 | 2.876 | 10.191 |
| | Intestine | 7.536 | 7.047 | 2.716 | 10.17 |
| | PBMC-cultured | 20.486 | 7.309 | 1.517 | 27.283 |
| | PBMC-blood | 3.58 | 2.894 | 1.203 | 5.156 |
| *HLA-DRB1* | Synovium | 29.021 | 24.545 | 10.216 | 37.902 |
| | Intestine | 17.531 | 16.65 | 6.834 | 22.706 |
| | PBMC-cultured | 53.603 | 35.305 | 10.341 | 53.387 |
| | PBMC-blood | 17.565 | 16.283 | 6.217 | 26.721 |

**Table S4. Percent change in estimated *HLA* expression after scHLApers.** For each classical *HLA* gene in each dataset (rows), the mean, median, $25^{th}$ and $75^{th}$ quantile of percent change in total UMI counts (sum across all cells per individual) using scHLApers relative to a standard pipeline without personalization.

**Table S5. Merging cell annotations across datasets to shared labels.** (See Kang_etal_SuppTables.xlsx, tab "Table S5")

Mapping between the cell annotations provided by the original dataset, major cell types in this study (B, myeloid, T, NK, fibroblast, or endothelial), and merged finer-grained annotations (for B, myeloid, and T cells in PBMC-blood and Synovium datasets).

**Table S6. Characteristics of MHC variants used for eQTL testing.** (See Kang_etal_SuppTables.xlsx, tab "Table S6")
Information regarding the 12,050 variants across the MHC used for eQTL testing, including chromosome 6 genomic position in GRCh38 (POS), REF and ALT alleles (for one- and two-field *HLA* alleles, A denotes absent, and T denotes present), imputation quality in Synovium (DR2), MAF in each cohort, and hg19 position (hg19_POS; as output by SNP2HLA based on MHC reference). For variant names, "rs" prefix indicates variant in the MHC region (dbSNP name), and "HLA" prefix indicates classical *HLA* allele.

**Table S7. Multi-cohort pseudobulk eQTL full results for myeloid, B, and T cells.** (See Kang_etal_TableS7.csv)
Results from testing each of 12,050 for association with classical *HLA* gene expression in each cell type (total 8 genes x 3 cell types x 12,050 variants = 289,200 tests) in the multi-cohort pseudobulk linear model. Columns list the variants in multi-cohort analysis, cell type, gene, effect size of variant on covariate-corrected standardized gene expression (beta), standard error of beta estimate, nominal *P*-value, and REF and ALT alleles. See **Table S6** for metadata about each tested variant.

**Table S8. Multi-cohort pseudobulk lead eQTL results for myeloid, B, and T cells.** (See Kang_etal_SuppTables.xlsx, tab "Table S8")
The lead eQTLs for each *HLA* gene and cell type in the multi-cohort pseudobulk linear model. Columns list the effect size (beta), standard error of beta estimate, nominal *P*-value, and REF and ALT alleles.

**Table S9. Comparison of effect sizes from multi-cohort vs. single-cohort pseudobulk eQTL models.** (See Kang_etal_SuppTables.xlsx, tab "Table S9")
Data for **Fig. 3D**. Columns list the lead variants from multi-cohort analysis, cell type, gene, dataset (either one of four single-dataset cohorts or the combined multi-dataset cohort), effect size of variant on covariate-corrected standardized gene expression (beta), standard error of beta estimate, nominal *P*-value, and REF and ALT alleles. See **Table S6** for metadata about each tested variant.

**Table S10. Grouping of classical *HLA* alleles by lead eQTLs.** (See Kang_etal_SuppTables.xlsx, tab "Table S10")
For each lead eQTL variant that was not itself an *HLA* allele, we determined the co-occurrence pattern between the eQTL variant REF and ALT alleles versus two-field classical *HLA* alleles for the eQTL-associated gene. Each row corresponds to one [eQTL]-[*HLA*-allele] pair, listing the number of haplotypes in the multi-ethnic HLA reference panel with the two-field *HLA* allele and the REF eQTL allele (nHaplos_wREF), number of haplotypes with the two-field allele and ALT eQTL allele (nHaplos_wALT), and proportion of total reference haplotypes with the two-field allele (nHaplos_withAllele) with the ALT version (prop_ALT).

**Table S11. Multi-cohort cell-type-interaction analysis results.** (See Kang_etal_SuppTables.xlsx, tab "Table S11")
Results from testing lead *HLA* eQTLs from multi-cohort pseudobulk analysis for cell-type interaction. For the lead eQTL in each gene/cell type pair, table lists the effect size (beta), standard error, and Wald *P*-value for the cell type it was the lead eQTL for, the LRT *P*-value from the mixed-effects model testing for cell type interaction, and the betas and Wald *P*-values

in each cell type (myeloid, B, and T, from the original multi-cohort pseudobulk model without cell type interaction) for comparison.

**Table S12. Multi-cohort pseudobulk conditional analysis results.** (See Kang_etal_TableS12.csv)
Results from conditional analysis identifying eQTLs, conditioning on the lead variant(s) from previous round(s). Columns list the variant, cell type, gene, round of conditional analysis (conditional_iter, ranging from 1 to 4 for primary to quaternary effects), effect size of eQTL (beta), standard error of beta estimate, and nominal *P*-value. Includes only variants with nominal P < 0.05 to reduce file size. See **Table S6** for metadata about each tested variant.

**Table S13. Proportion of gene expression variance explained by cell state.** (See Kang_etal_SuppTables.xlsx, tab "Table S13")
The estimated proportion of variance in each classical *HLA* gene explained by cell state (first 10 tissue-defined hPCs) in Synovium and PBMC-blood in myeloid, B, and T cells. Columns indicate estimated $R^2$ for the full NBME model (full_rsq), $R^2$ for a model without cell state (nostate_rsq), and the difference (full_rsq - nostate_rsq) representing variance explained by cell state.

**Table S14. Testing eQTLs for cell-state interaction with single-cell NBME model.** (See Kang_etal_SuppTables.xlsx, tab "Table S14")
Results from testing lead *HLA* eQTLs for cell-state dependence using the single-cell NBME model with cell state defined using the top 10 tissue-defined hPCs per cell type. For each gene, lead eQTL variant, dataset, and cell type (row), column E lists the significance (LRT *P*-value) of the genotype main effect as determined using a NBME model with genotype but without cell state terms (used to define 58 variant-gene pairs with robust main effects). Columns F-M show the results from the NBME model testing for cell-state interactions: the hPC with the most significant interaction with genotype ($\beta_{G\times hPC}$, max_int_term), the interaction effect size and Wald *P*-value, the genotype main effect ($\beta_G$, G_main_Estimate) and its Wald *P*-value, the size of the maximum interaction effect size in proportion to the genotype main effect (int_prop_main), and the significance of cell-state-dependency (LRT *P*-value and Chi-Square statistic comparing the full model for all hPCs to a null model without cell state interaction terms).

| **Gene** | **Mean $\chi^2$ statistic** | **Number of tests** |
|---|---|---|
| *HLA-A* | 133.1 | 9 |
| *HLA-B* | 61.7 | 7 |
| *HLA-C* | 177.8 | 9 |
| *HLA-DPA1* | 39.4 | 6 |
| *HLA-DPB1* | 43.1 | 6 |
| *HLA-DQA1* | 596.9 | 8 |
| *HLA-DQB1* | 220.4 | 8 |
| *HLA-DRB1* | 58.1 | 5 |
| Total | | 58 |

**Table S15. Degree of cell-state-dependency by gene.** The mean LRT $\chi^2$ statistic value from testing for cell-state-dependence across all variant-dataset-cell-type tests (and number of tests performed) for each gene.

### Supplementary Text 2: More Stringent Quality Control of OneK1K Cohort Single-cell RNA-seq Dataset

Laurie Rumker
Joyce B. Kang
Soumya Raychaudhuri

March 13, 2023

#### 1 Motivation

In collaboration with the authors of the OneK1K dataset index publication [1], we applied more stringent quality control to these PBMC scRNA-seq profiles before employing the dataset in our own analyses. Our additional dataset processing, summarized in this document, was prompted by our observations of isolated populations with mixed type assignments that expressed unexpected marker genes. We initially observed these putative doublet populations when performing a standard PCA-based analysis on each major cell type separately (e.g. PCA on cells labeled B cells). We observed fragmented cell populations with mixed type assignments (e.g. mixed B naive and B memory labels in a population separate from the major populations for these B cell subtypes) that also contained expression of unexpected marker genes that did not match the assigned labels (e.g. CD3 among these "B cells"). We we found that these populations corresponded to droplets identified as doublets by Demuxlet [2] or Scrublet [3] but not previously removed from the dataset.

#### 2 Approach

We received scRNA-seq profiling (cells-by-counts matrix), as well as Demuxlet and Scrublet method output, directly from the study authors. Cell type assignments provided by the study authors were based on Azimuth mapping to a PBMC reference dataset [4]. After affirming basic per-profile QC thresholds were met (>200 genes, <8% mitochondrial gene reads), and removing 7680 genes that appeared in fewer than three profiles, we subdivided the profiles by major cell type using the following mapping from the 31 available labels to 7 major types:

- CD4+ T = [CD4 TCM, CD4 Naive, CD4 TEM, Treg, CD4 CTL, CD4 Proliferating]
- Other T = [CD8 TEM, CD8 Naive, CD8 TCM, MAIT, CD8 Proliferating, gdT, dnT]
- NK = [NK, NK_CD56bright, NK Proliferating, ILC]
- Monocyte = [CD14 Mono, CD16 Mono]
- DC = [cDC1, cDC2, pDC, ASDC]
- B = [B naive, B memory, B intermediate, Plasmablast]
- Other = [HSPC, Platelet, Eryth]

For each major cell type, we followed standard processing using scanpy (with parameters as described in the "Preprocessing and clustering 3k PBMCs" tutorial unless otherwise specified [5]) to total-count normalize to 10,000 reads per profile, logarithmize the data, retain only highly-variable genes and compute principal components (PCs). For each major cell type, we corrected these PCs for batch with harmony (batch = "pool", nclust = 50, sigma = 0.2, max_iter_harmony = 50) to generate hPCs. Resuming the scanpy pipeline, we used these hPCs to construct a nearest-neighbor graph and UMAP embedding per major cell type.

The index publication authors had previously removed any droplet identified as a doublet by both Scrublet and Demuxlet, but retained all droplets identified as doublets by only one of these two methods. Of the 1,249,037 profiles provided by the OneK1K dataset authors, 22,662 were identified as doublets by Scrublet (predicted_doublet_mask==True) and 382,464 were called as doublets by Demuxlet ('BEST' assignment to 'DBL-'). We chose to remove these profiles. None of the cells included in the published dataset provided by the original authors had been classified by Demuxlet as ambiguous. Given that many profiles identified as doublets by Scrublet or Demuxlet were observed to cluster together transcriptionally in the dataset (Figures 1, 2, 3, 4, 5, 6, 7), we performed fine-grained clustering within each major cell type and removed any clusters for which >2/3 profiles were identified as doublets (by either Demuxlet or Scrublet).

We used Wilcoxon rank-sum tests to identify differentially-expressed genes per fine-grained cluster (scanpy's rank_gene_groups function with method = 'wilcoxon'). For major cell type groups besides "Other"—which contains the profiles assigned by Azimuth to the Platelet type—we also removed fine-grained clusters for which differential expression analysis identified PPBP, PF4, GP1BB and NRGN among the top 6 cluster-characteristic markers, suggestive of platelet doublets.

Finally, 1803 profiles lacked results from Demuxlet and Scrublet. Of these profiles, 131 were labeled "Doublet" by the publication authors and the remainder corresponded to an individual who also failed genotype data quality control in our analyses (not described here). We removed these 1083 profiles.

In summary, we removed each profile if:

- The profile was identified as a doublet by either Demuxlet or Scrublet OR
- The profile was assigned to a doublet-dominated fine-grained cluster OR
- The profile was labeled as a non-platelet type but assigned to a fine-grained cluster characterized by platelet-related genes OR
- The profile lacked doublet-calling results

Finally, we reassigned cell type labels to our retained cells, applying the same approach used by the publication authors for the initially-provided cell type labels: Azimuth reference mapping to the Azimuth PBMC reference. To accommodate Azimuth data volume limitations, we split the total dataset into 15 subsets by batch pool group and applied Azimuth separately to each subset. The major cell type classifications for the retained cells (i.e. among T, B, NK, and Myeloid groups) were unchanged for the vast majority of cells when compared to each cell's original major type assignment.

### 3 Results

Of the 1,249,037 profiles provided by the study authors from the published dataset, we chose to remove 416,556 (33%), the vast majority of which (405,126 profiles, 97%) were identified as doublets by either Scrublet or Demuxlet, and the remainder selected using our other two criteria (Tables 1, 2).

We found that the droplets identified as doublets by Scrublet or Demuxlet largely explained the isolated cell populations with mixed assigned types that we had observed, and platelet-contaminated populations explained some remaining fragmented populations (Figures 1, 2, 3, 4, 5, 6, 7, 8).

| Major Type | Profiles | Resolution | Demuxlet | Scrublet | Fraction Removed |
|---|---|---|---|---|---|
| DC | 6648 | 1.0 | 0.2 | 0.04 | 0.28 |
| Mono | 51876 | 2.0 | 0.22 | 0.02 | 0.25 |
| B | 129588 | 3.0 | 0.29 | 0.02 | 0.32 |
| NK | 172397 | 4.0 | 0.29 | 0.02 | 0.33 |
| CD4+ T | 624592 | 6.0 | 0.32 | 0.01 | 0.34 |
| Other T | 259893 | 6.0 | 0.31 | 0.03 | 0.34 |
| Other | 3912 | 0.2 | 0.44 | 0.04 | 0.61 |

Table 1: Profiles selected for removal, by major type. For each major type group, the total number of profiles assigned to that group ("Profiles") is shown, along with the resolution used for fine-grained clustering ("Resolution"), the fraction of all profiles identified as doublets by Demuxlet or Scrublet ("Demuxlet" and "Scrublet", respectively), and the fraction selected for removal based on all criteria ("Fraction Removed")

| Removed | Scrublet Dblt. | Demuxlet Dblt. | Platelet Clust. | Dblt. Clust. | Count |
|---|---|---|---|---|---|
| F | F | F | F | F | 832481 |
| T | F | F | F | T | 7734 |
| T | F | F | T | F | 1893 |
| T | F | T | F | F | 358161 |
| T | F | T | F | T | 23198 |
| T | F | T | T | F | 1105 |
| T | T | F | F | F | 18602 |
| T | T | F | F | T | 4033 |
| T | T | F | T | F | 27 |
| T | NA | NA | NA | NA | 1803 |

Table 2: Profiles selected for removal, by criterion. "Removed": T if the profile was selected for removal. "Scrublet Dblt.": T if Scrublet identified the profile as a doublet. "Demuxlet Dblt.": T if Demuxlet identified the profile as a doublet. "Platelet Clust.": T if the profile was assigned to a fine-grained cluster characterized by platelet-related genes. "Dblt. Clust.": T if the profile was assigned to a fine-grained cluster with <2/3 doublets. "Count": The number of profiles matching the combination of features captured by the corresponding row. The authors had previously removed profiles called as doublets by both Scrublet and Demuxlet. The final row captures profiles for which doublet-calling results were not available.

We make available a table containing the results of this data processing. This table contains one row per cell, indexed by barcode. In addition to cell-level metadata provided in the published dataset, we have added the following columns:

- demuxlet_DBL: True iff the cell was assigned as a doublet by Demuxlet
- demuxlet_AMB: True iff the cell was assigned as ambiguous by Demuxlet
- scrublet_DBL: True iff the cell was assigned as a doublet by Scrublet
- scrublet_score: Score assigned by Scrublet
- preQC_Azimuth_type: Azimuth-based cell types shared by the publication authors
- DBL_cluster: True iff the cell belonged to a cluster with >2/3 cells assigned as doublets by Scrublet or Demuxlet
- Platelet_cluster: True iff the cell belonged to a cluster characterized by platelet-associated marker genes
- remove_cellQC: True iff the cell met one of the four critera for removal described here
- remove_sampleQC: True iff the cell was associated with a sample we removed for our analyses (samples with low-quality or missing genotyping data, or labeled as ethnic outliers)
- fail_QC: True iff remove_cellQC or remove_sampleQC is True
- celltype: Azimuth cell type assignments for retained cells
- majortype: Major cell type assignments, aggregated from celltype

### 4 Discussion

Identification and removal of doublet droplets is a crucial quality control step in single-cell data analysis. Scrublet and Demuxlet are two of many available methods to accomplish this task. Scrublet simulates doublet transcriptional profiles as combinations of observed profiles and compares the observed profiles to these simulates. Demuxlet identifies droplets whose transcripts reflect a combination of genetic variants unlikely to arise from a single individual in the dataset. Because these methods have contrasting failure modes, applying both to the same dataset can enable the detection of droplets

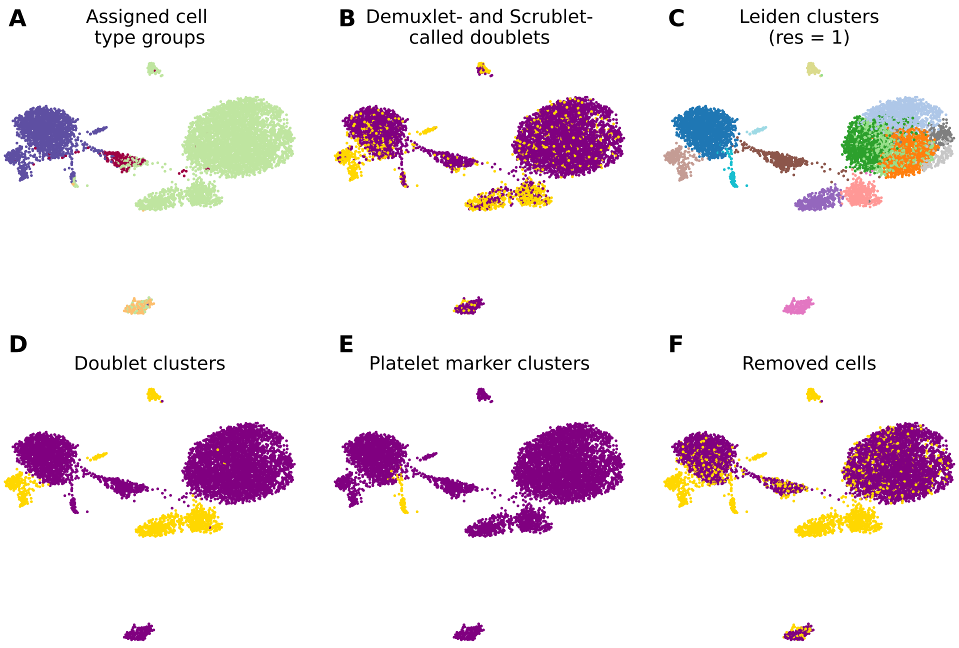

Figure 1: **Dendritic cells.** (**A**) Profiles colored by type label, as provided by the publication authors. (**B**) Profiles identified by either Scrublet or Demuxlet as doublets, in gold. (**C**) Profile assignments to fine-grained clusters. (**D**) Clusters containing >2/3 profiles called as doublets by either Scrublet or Demuxlet, in gold. (**E**) Clusters characterized by platelet-related genes, in gold. (**F**) All profiles selected for removal, in gold; the union of gold profiles in B, D, and E.

by one method that were missed by the other. In the original publication of the OneK1K dataset, only cells called as doublets on the basis of both Scrublet and Demuxlet were removed, a small fraction of all identified doublets. Of the retained profiles identified as doublets, the vast majority were flagged by Demuxlet (i.e. on the basis of contrasting genotypes detected in the same droplet). We found that the retained doublets were transcriptionally perturbed in the dataset relative to cells identified as singlets and have chosen a more stringent quality control approach to remove these cells. In collaboration with the OneK1K dataset authors, we make available a table indicating which cells were selected for removal in our more stringent quality control.

[4] Yuhan Hao, Stephanie Hao, Erica Andersen-Nissen, William M. Mauck, Shiwei Zheng, Andrew Butler, Maddie J. Lee, Aaron J. Wilk, Charlotte Darby, Michael Zager, Paul Hoffman, Marlon Stoeckius, Efthymia Papalexi, Eleni P. Mimitou, Jaison Jain, Avi Srivastava, Tim Stuart, Lamar M.

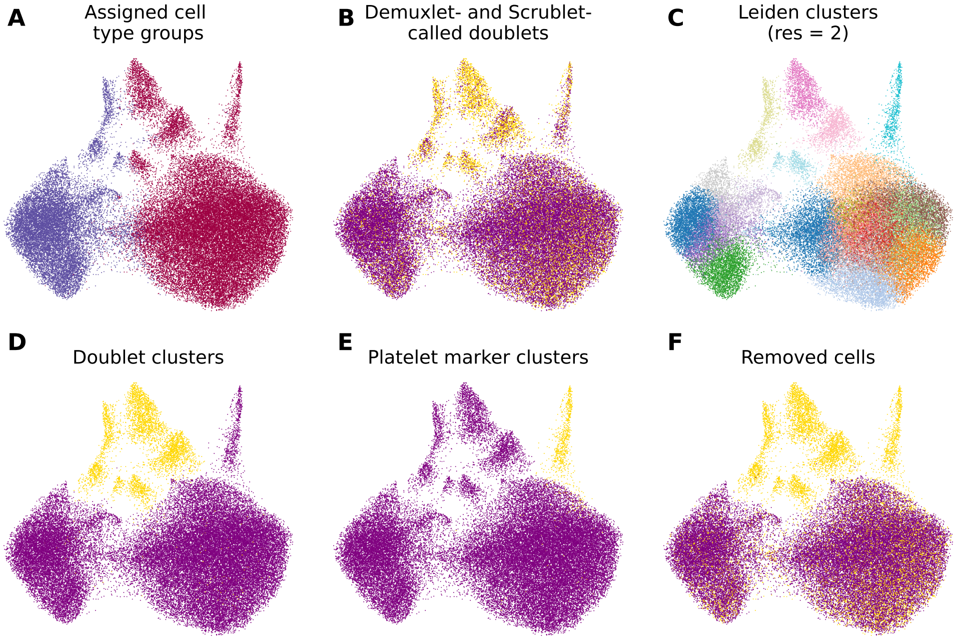

Figure 2: **Monocytes.** (**A**) Profiles colored by type label, as provided by the publication authors. (**B**) Profiles identified by either Scrublet or Demuxlet as doublets, in gold. (**C**) Profile assignments to fine-grained clusters. (**D**) Clusters containing >2/3 profiles called as doublets by either Scrublet or Demuxlet, in gold. (**E**) Clusters characterized by platelet-related genes, in gold. (**F**) All profiles selected for removal, in gold; the union of gold profiles in B, D, and E.

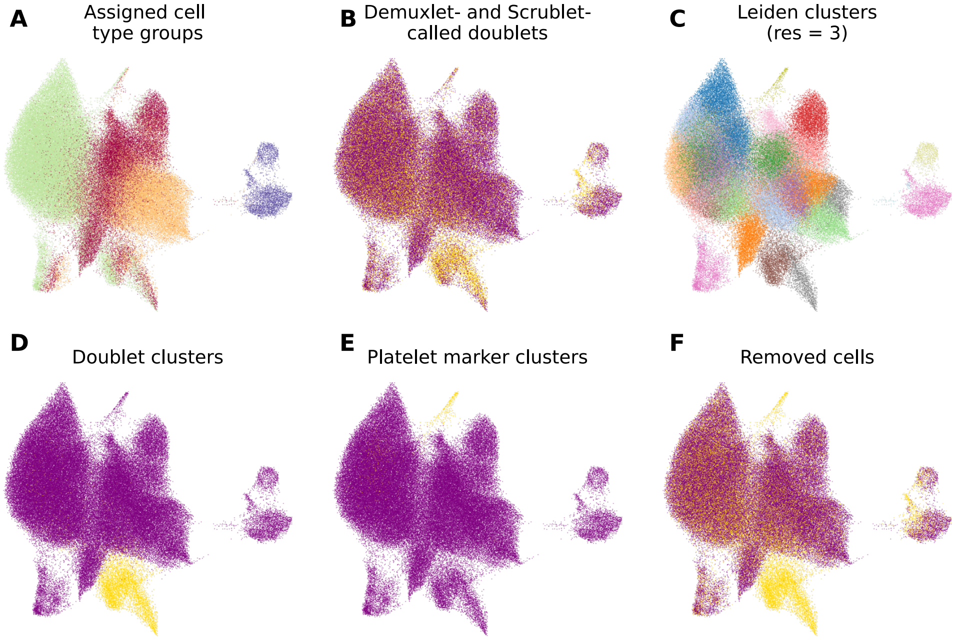

Figure 3: **B cells.** (**A**) Profiles colored by type label, as provided by the publication authors. (**B**) Profiles identified by either Scrublet or Demuxlet as doublets, in gold. (**C**) Profile assignments to fine-grained clusters. (**D**) Clusters containing >2/3 profiles called as doublets by either Scrublet or Demuxlet, in gold. (**E**) Clusters characterized by platelet-related genes, in gold. (**F**) All profiles selected for removal, in gold; the union of gold profiles in B, D, and E.

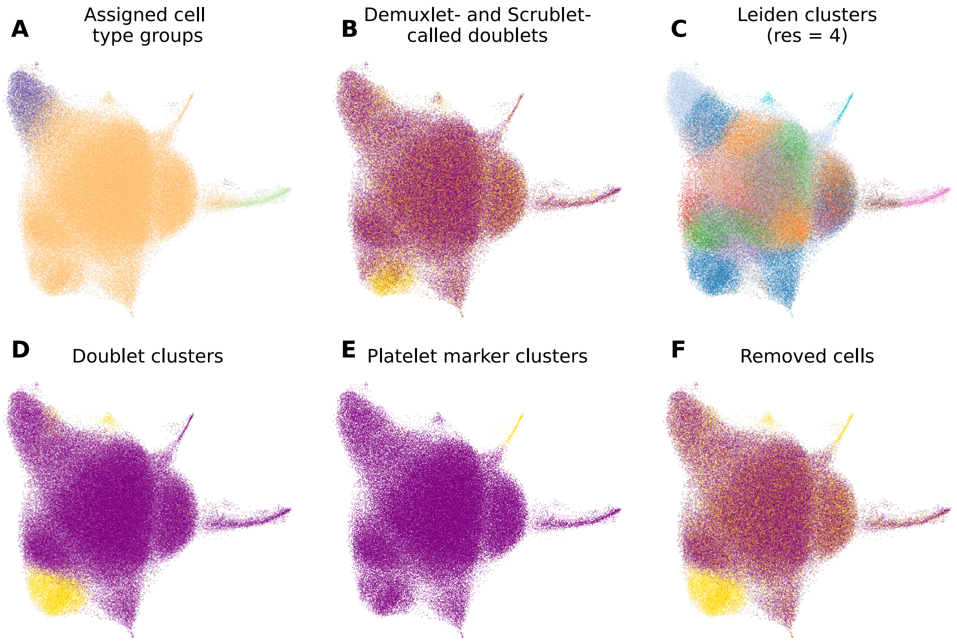

Figure 4: **NK cells.** (**A**) Profiles colored by type label, as provided by the publication authors. (**B**) Profiles identified by either Scrublet or Demuxlet as doublets, in gold. (**C**) Profile assignments to fine-grained clusters. (**D**) Clusters containing >2/3 profiles called as doublets by either Scrublet or Demuxlet, in gold. (**E**) Clusters characterized by platelet-related genes, in gold. (**F**) All profiles selected for removal, in gold; the union of gold profiles in B, D, and E.

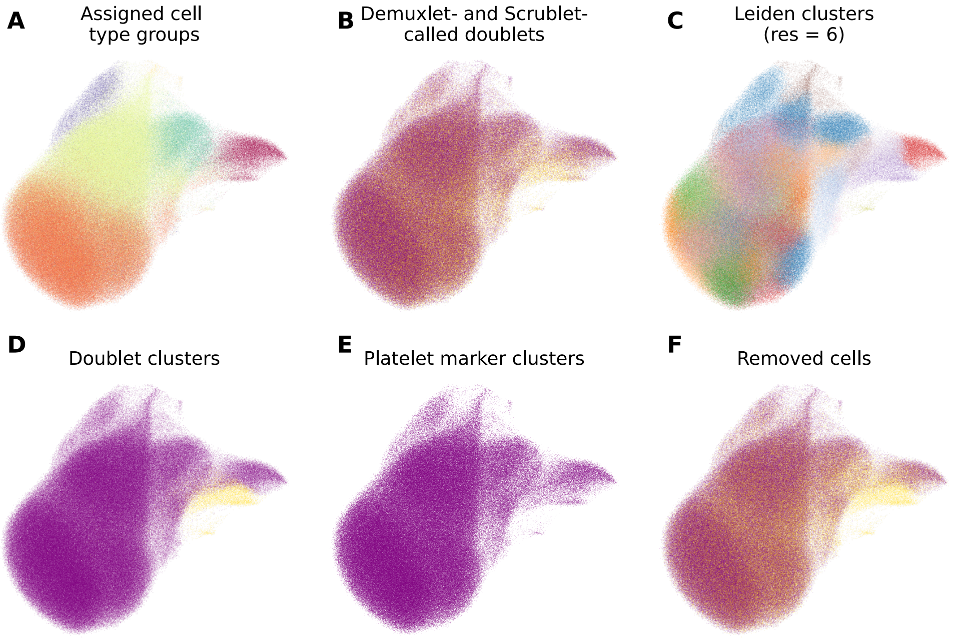

Figure 5: **CD4+ T cells.** (**A**) Profiles colored by type label, as provided by the publication authors. (**B**) Profiles identified by either Scrublet or Demuxlet as doublets, in gold. (**C**) Profile assignments to fine-grained clusters. (**D**) Clusters containing >2/3 profiles called as doublets by either Scrublet or Demuxlet, in gold. (**E**) Clusters characterized by platelet-related genes, in gold. (**F**) All profiles selected for removal, in gold; the union of gold profiles in B, D, and E.

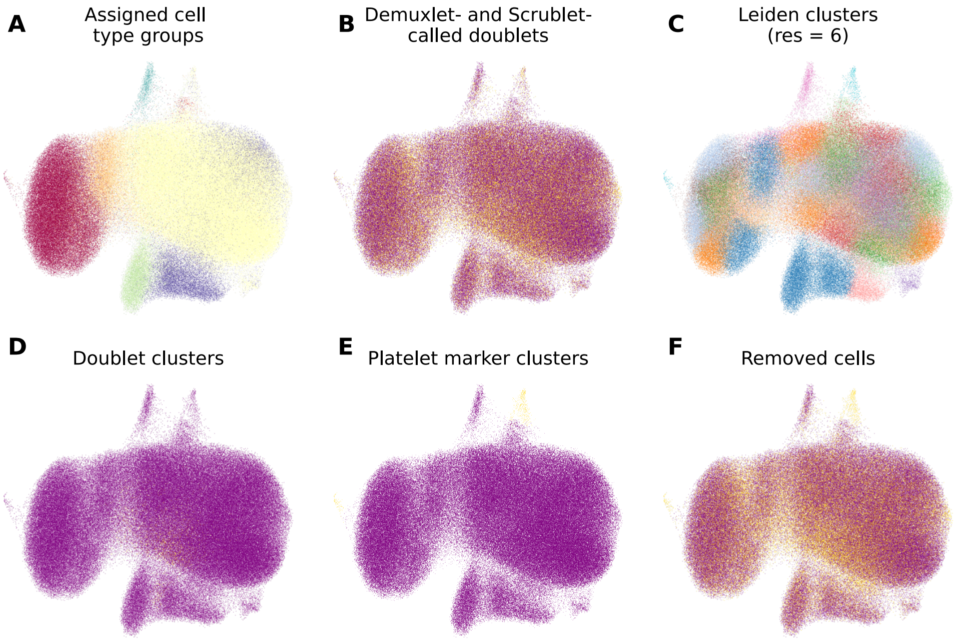

Figure 6: **Other T cells.** (**A**) Profiles colored by type label, as provided by the publication authors. (**B**) Profiles identified by either Scrublet or Demuxlet as doublets, in gold. (**C**) Profile assignments to fine-grained clusters. (**D**) Clusters containing >2/3 profiles called as doublets by either Scrublet or Demuxlet, in gold. (**E**) Clusters characterized by platelet-related genes, in gold. (**F**) All profiles selected for removal, in gold; the union of gold profiles in B, D, and E.

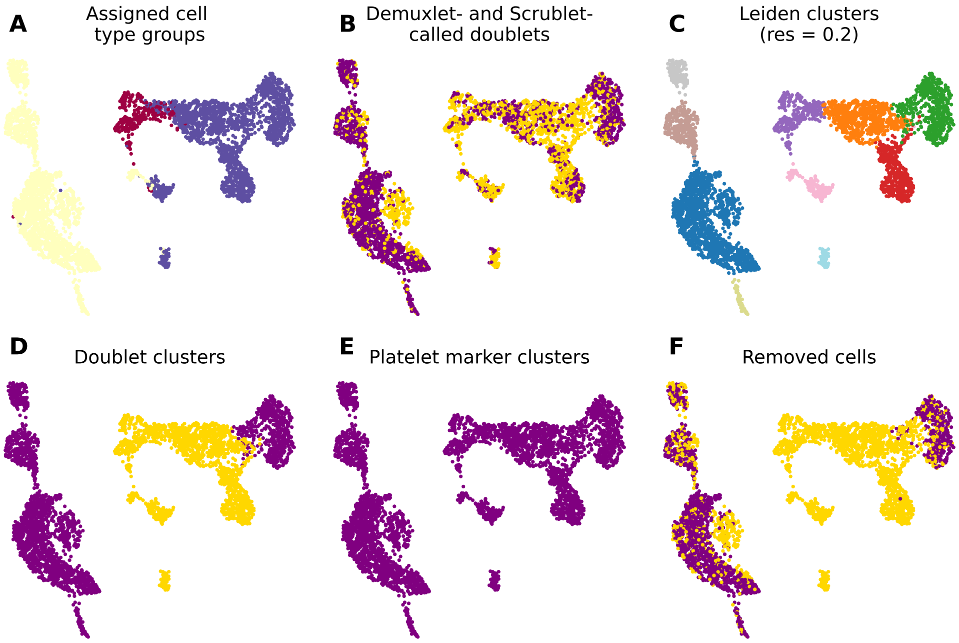

Figure 7: **All other cells.** (**A**) Profiles colored by type label, as provided by the publication authors. (**B**) Profiles identified by either Scrublet or Demuxlet as doublets, in gold. (**C**) Profile assignments to fine-grained clusters. (**D**) Clusters containing >2/3 profiles called as doublets by either Scrublet or Demuxlet, in gold. (**E**) Clusters characterized by platelet-related genes, in gold. (**F**) All profiles selected for removal, in gold; the union of gold profiles in B, D, and E.

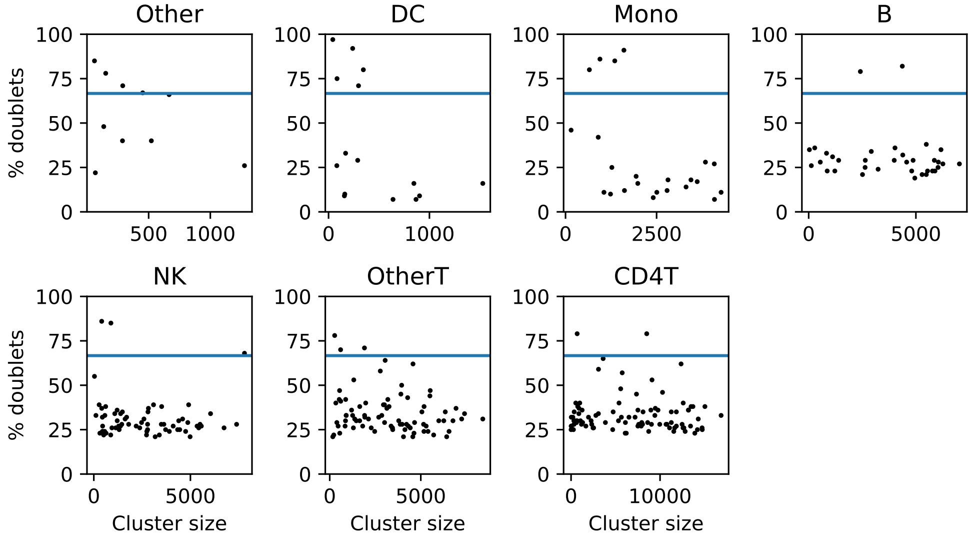

Figure 8: **Doublet cluster identification.** The profiles within each major type were clustered at a fine-grain resolution to identify and remove doublet-predominant clusters, in addition to isolated doublet profiles. The fraction of profiles called as doublets (by Scrublet or Demuxlet) for each fine-grained cluster is shown along the y axis, while the size of each cluster (number of profiles) is shown along the x axis, with plots separated by major type. Clusters with >2/3 doublets, above the blue line, were selected for removal.

Fleming, Bertrand Yeung, Angela J. Rogers, Juliana M. McElrath, Catherine A. Blish, Raphael Gottardo, Peter Smibert, and Rahul Satija. Integrated analysis of multimodal single-cell data. *Cell*, 184(13):3573–3587.e29, June 2021. Publisher: Elsevier.

[5] Rahul Satija, Jeffrey A Farrell, David Gennert, Alexander F Schier, and Aviv Regev. Spatial reconstruction of single-cell gene expression data. *Nature Biotechnology*, 33(5):495–502, May 2015.
